# Short- and Long-term Outcomes of children hospitalized with COVID-19 or Influenza: results of the AUTCOV study

**DOI:** 10.1101/2024.08.28.24312702

**Authors:** Christine Wagenlechner, Ralph Wendt, Berthold Reichardt, Michael Mildner, Julia Mascherbauer, Clemens Aigner, Johann Auer, Hendrik Jan Ankersmit, Alexandra Christine Graf

## Abstract

**Background:** Recent literature gives different results on morbidity and mortality after COVID-19 as compared to Influenza hospitalized children and results of large, population based studies are scant. In this population-based study in Austria, we evaluated and compared the short- and long-term outcomes after COVID-19 or Influenza hospitalization and associations with their baseline drug profile.

**Methods:** Individual data were provided on children ≤ 18 years hospitalized with COVID-19 in the years 2020 and 2021 or Influenza in 2016 – 2021 as well as on age-, sex- and region-matched controls from the Austrian Health Insurance Funds. The primary outcome was time to hospital discharge. Secondary outcomes were in-hospital death, all-cause mortality and readmission to hospital due to any reason. The median follow-up time was 430 days (IQR: 245-552) in the COVID-19 and 1221 days (IQR: 881-1599) in the Influenza group.

**Results:** 1063 children were hospitalized due to COVID-19 and 2781 children due to Influenza in the study period. Children hospitalized due to COVID-19 or Influenza were more likely to have a larger disease burden as compared to the general population. Influenza hospitalized patients were observed to be generally younger and a larger percentage of polypharmacy than those with COVID-19. No significant difference in the time to hospital discharge was found between cohorts (HR: 1.22 [95%-CI: 0.97-1.55], p=0.093). The risk for readmission was significantly higher for Influenza (HR: 1.23 [95%-CI: 1.03-1.47], p=0.021). In-hospital mortality (0.94% vs. 0.22%, p=0.004) and long-term mortality (p=0.009) was significantly larger in COVID-19 patients. One-year mortality after hospitalization was estimated with 1.13% (CI: 0.49-1.77) in the COVID and 0.32% (CI: 0.11-0.53) in the Influenza group.

**Conclusion:** A general picture of COVID-19 being a milder disease compared to Influenza may not be drawn. No significant difference for time to hospital discharge was observed between cohorts but the risk of readmission was significantly larger in the Influenza group. Death rates of COVID-19 hospitalized children seem to be higher, however, the low number of severe events may limit the findings.

## Introduction

The outbreak of Coronavirus Disease 2019 (COVID-19) has put great strain on hospitals and health systems. In Austria and other European countries, the pandemic led to several “lockdowns” to flatten the curve of cases and to avoid an overload of the health systems. In Austria, children and adolescents were faced with kindergarten and school closures.

Age and gender are well-known risk factors for severe COVID-19 related mortality and morbidity.^(1-4)^ Recent literature discusses that the presence of comorbidities may increase the risk of COVID-19-related death in adult patients.^(1,5-8)^ Furthermore, an increased risk of readmission and mortality after hospital discharge has been observed after COVID-19 hospitalization.^(8-12)^ However, patients who require hospitalization for COVID-19 may have a greater comorbidity burden, more severe initial diseases, higher rates of polypharmacy and are expected to have worse short-term outcomes.^(8,12-16)^

The case-fatality rate in COVID-19 is assumed to be higher than in seasonal Influenza, even though both diseases mainly affect older adults with frailty and the health burden of patients with COVID-19 and seasonal influenza requiring hospitalization may differ. ^(17-19)^

Children appear less susceptible to infection by COVID-19 as compared to other viruses such as Influenza and RSV^(20,21)^ and may be at lower risk of hospitalization and life-threatening conditions.^(22)^ Children, less frequently, have risk factors such as co-morbidities and obesity. But adolescents and children with pre-existing illnesses may be high risk groups for severe outcomes.^(20)^

However, population based studies evaluating short and long-term outcomes as well as their baseline medication profiles of children aged ≤18 after COVID-19 and Influenza hospitalization are scant and published study results are conflicting.^(23)^

We present data on children ≤18 years of age, hospitalized due to COVID-19- or Influenza and corresponding age-, sex- and region- matched control groups, representing the general population, provided by the Austrian Health Insurance Funds with a median follow-up of 430 days (IQR: 245-552) for COVID-19 and 1221 days (IQR: 881-1599) days for Influenza hospitalized patients. The aim of this study was to analyze short-term (time to hospital discharge, in-hospital death) as well as long-term outcomes (readmission to hospital, all-cause death) of COVID-19 and Influenza hospitalized children. Furthermore, we wanted to evaluate the association between the baseline medication profiles of hospitalized children and outcomes and compared these characteristics with age-, sex-, and region-matched control groups. This investigation, presenting a real-world picture, may help policy makers, health economists, and physicians to identify children at risk for adverse outcomes.

## Methods

### Study Design and Cohorts

This retrospective, national population-based study adheres to the Declaration of Helsinki and was approved by the ethics committee of Lower Austria (GS1-EK-4/747-2021). The data were available from the Austrian Health Insurance Funds. Approximately 98% of the Austrian population is registered in the public health insurance system. Health care in Austria is a national system with good access to care, few malpractice lawsuits, and little tendency toward overuse of medical resources. Access to individual services is regulated by social insurance law.^(24)^

Patients ≤18 years of age, hospitalized in Austria due to the main diagnosis COVID-19 (ICD-10 Codes U071, U072, U049) from 1 January 2020 to 31 December 2021 (i.e., including patients from the first four pandemic waves) and patients ≤18 hospitalized in Austria due to the main diagnosis Influenza (ICD-10 codes: J09, J100, J101, J108, J110, J111, J118, J10) from 30 December 2015 to 31 December 2021 were included in this study. Patient characteristics (age, sex, region, and medication profiles) were obtained from 1 year before index-hospitalization until study cut-off.

For both groups, age-, sex-, and region-matched control groups (approximately 10 controls for each patient) consisting of individuals not hospitalized due to COVID-19 (in the years 2020 and 21) as well as Influenza (in the years 2016 – 21) were randomly chosen from the population registered in the Austrian Health Insurance Fund. Data on the control groups were available from 1 year before the first patient was hospitalized until study cut-off. Death dates were available at study cut-off.

### Study Outcomes

The primary outcome of the study was time from hospital admission to hospital discharge. Secondary outcomes were readmission after hospital discharge due to any reason (conditional on hospital survival), in-hospital death and all-cause death. Readmission was defined based on billing information (MEL codes) available from the Austrian Health Insurance Funds. For patients, we used the first readmission after the index hospital stay. Readmissions within 2 weeks after the index admission were assumed to belong to the index admission and were not counted as readmission. For controls, in-hospital death and time to death was evaluated from the index hospital admission date of the matched patient (Table S1).

### Statistical Analysis

For each patient, age, sex, region (Vienna, Lower Austria, Upper Austria, Burgenland, Carinthia, Salzburg, Tyrol, Styria, or Vorarlberg), and ATC codes describing prescribed medications were available from the Austrian Health Insurance Funds from 1 year before the index hospitalization. ATC codes for medications before hospitalization were summarized in medication groups (Table S2).

A binary variable was defined for each medication group, which was set to 1 if a drug of the corresponding ATC-codes was prescribed at least once 1 year before the index COVID-19 hospitalization and 0 if not. Data was evaluated for 30 medication groups (MG 1: Anticoagulants, MG 2: Antibiotics, Antivirals, Antiprotozoals or Anthelmintics, MG 3: Insulins and other Antidiabetics, MG 4: heart drugs, MG 5: Antihypertensives incl. Diuretics and Renin-angiotensin-aldosterone system inhibitors, MG 6: Beta Blockers, MG 7: Statins, Fibrates incl. Proprotein convertase subtilisin/kexin type 9 inhibitors and Inclisiran, MG 8: Immunosuppressants and Immunomodulators, MG 9: Systemic Steroids, MG 10: Chemotherapy, MG 11: Iron supplements, Erythropoietic stimulating agents, Vitamin B12, folic acid, MG 12: Antacids incl. Antihistamines, MG 13: Vitamin D and other Vitamin supplements, MG 14: Caplacizumab, MG 15: Systemic Hemostatics, MG 16: Hereditary angioedema Therapeutics, MG 17: Peripheral Vasodilators, MG 18: Hormonal contraceptives and similar hormone preparations, MG 19: Immunoglobulins, MG 20: Interferons and CSF, MG 21: NSAR and other anti-inflammatory drugs, MG 22: Gout medications, MG 23: Antiepileptics, MG 24: Antipsychotics, MG 25: Rhinological and throat antiseptics, MG 26: inhaled anti-obstructive drugs, MG 27: inhaled steroids, MG 28: other COPD drugs, MG 29: Cold and Cough preparations, MG 30: Systemic Antihistamines). Due to underrepresentation, medication groups prescribed for less than 5 patients in the COVID-19 and Influenza group were not evaluated for statistical modelling. For controls, a similar medication profile was generated using the drugs prescribed 1 year before the index hospital stay of the matched patient.

Note that data of the AUTCOV-study regarding adults (patients and controls >18 years of age) were published on medRxiv^(8)^ using similar definitions for medication groups and outcomes as discussed for the presented analyses.

All medication groups, except Rhinological and throat antiseptics (MG 25) and hormonal contraceptives and similar hormone preparations (MG18) were summed up for the number of medication groups, categorized in 0, 1, ≥2. For the category ≥2 we use the term “polypharmacy”.

Numbers and percentages were used to summarize categorical variables, medians or interquartile ranges for continuous variables. Medication groups were compared between groups using Fisher-Exact tests.

To evaluate the association between medication groups, polypharmacy and time to hospital discharge, first, for each medication group simple Cox regression models were calculated accounting for sex, age (categorized in 0-5, 6-10, 11-15 and 16-18), polypharmacy, wave and the respective medication group with clustering variable region. Due to the very small number of deaths, no competing risk model was performed. Patients dying during the in-hospital follow-up were censored at the time of death. A multivariable Cox-Regression model was performed including sex, age, polypharmacy, wave and all medication groups with a p-value smaller than 0.1 in the simple models. These analyses were performed separately for the COVID-19 and the Influenza Group.

To compare time to discharge between COVID-19 and Influenza hospitalized patients, we first used propensity score matching to account for potential bias. Propensity score matching was based on age, sex, region and all medication groups with a p<0.05 in the comparison between cohorts using Fishers exact test. To evaluate the association between group (COVID-19, Influenza) and time to hospital discharge, first, for each medication group simple Cox regression models were calculated accounting for group, sex, age, polypharmacy, wave and the respective medication group with clustering variable region. A multivariable Cox-Regression model was performed including sex, age, polypharmacy, wave and all medication groups with a p-value smaller than 0.1 in the simple models. For time to discharge, a hazard ratio smaller than one is indicating a larger probability for hospital discharge in the reference group, i.e. patients in the reference group are more likely to have a shorter hospital stay.

Time to readmission was evaluated using similar analyses within groups and to compare both groups. For time to readmission, a hazard ratio smaller than one is indicating a larger probability for readmission in the reference group. For a detailed definition of medication groups and confounder, see supplemental Tables S1 and S2.

Due to the small number of deaths, no statistical modelling or PSM could be performed. In-hospital mortality was exploratory compared with Fisher-Exact test and all-cause mortality using log-rank test.

Schönfeld residuals were used to evaluate the proportional hazard assumption and variance inflation factors to evaluate multicollinearity. A p-value smaller than 0.05 was considered significant. All analyses were performed using R, release 4.2.2.^(25)^

## Results

The study included 1063 COVID-19 hospitalized and 2781 Influenza-hospitalized children as well as 10626 age-, sex- and region matched COVID-19 and 27634 Influenza-controls with a follow-up in the timeline as the matched patients (Table 1, Figure 1A, C). The median follow-up time was 430 days (IQR: 245-552) in the COVID and 1221 days (IQR: 881-1599) in the Influenza group. Note that during 2020 and 2021 hospitalization due to Influenza was rare.

**Table 1.** Demographic data for COVID-19- and Influenza-hospitalized patients in Austria.

| Covid Patients |  |  |  |  |  |  |  |  |  |  |  |
| --- | --- | --- | --- | --- | --- | --- | --- | --- | --- | --- | --- |
| Age |  | 0-5 |  | 6-10 |  | 11-15 |  | 16-18 |  | Overall |  |
|  |  | n = 609 |  | n = 98 |  | n = 167 |  | n = 189 |  | n = 1063 |  |
|  |  | n | % | n | % | n | % | n | % | n | % |
| Sex | M | 339 | 55.67 | 42 | 42.86 | 81 | 48.5 | 70 | 37.04 | 532 | 50.05 |
|  | W | 270 | 44.33 | 56 | 57.14 | 86 | 51.5 | 119 | 62.96 | 531 | 49.95 |
| Medication groups | 0 | 485 | 79.64 | 43 | 43.88 | 84 | 50.3 | 93 | 49.21 | 705 | 66.32 |
|  | 1 | 78 | 12.81 | 29 | 29.59 | 40 | 23.95 | 47 | 24.87 | 194 | 18.25 |
|  | ≥2 | 46 | 7.55 | 26 | 26.53 | 43 | 25.75 | 49 | 25.93 | 164 | 15.43 |
| Follow-up in days | Median (IQR) | 366 (232.75-528.25) |  | 441 (260-576) |  | 453 (278-567) |  | 450 (298-588) |  | 430 (245-551.5) |  |
| Time to death | Median(IQR) | 6 (2-21) |  | 201 (201-201) |  | 8 (4-12) |  | 32.5 (22.25-64.25) |  | 18.5 (4.25-35.5) |  |
| Length of hospital stay | Median(IQR) | 3 (2-5) |  | 3 (2-6) |  | 3 (2-6) |  | 3 (2-6) |  | 3 (2-5) |  |
| Influenza patients |  |  |  |  |  |  |  |  |  |  |  |
| Age |  | 0-5 |  | 6-10 |  | 11-15 |  | 16-18 |  | Overall |  |
|  |  | n = 1961 |  | n = 480 |  | n = 207 |  | n = 133 |  | n = 2781 |  |
|  |  | n | % | n | % | n | % | n | % | n | % |
| Sex | M | 1063 | 54.21 | 270 | 56.25 | 116 | 56.04 | 63 | 47.37 | 1512 | 54.37 |
|  | W | 898 | 45.79 | 210 | 43.75 | 91 | 43.96 | 70 | 52.63 | 1269 | 45.63 |
| Medication groups | 0 | 1023 | 52.17 | 211 | 43.96 | 88 | 42.51 | 38 | 28.57 | 1360 | 48.9 |
|  | 1 | 570 | 29.07 | 161 | 33.54 | 62 | 29.95 | 48 | 36.09 | 841 | 30.24 |
|  | ≥2 | 368 | 18.77 | 108 | 22.5 | 57 | 27.54 | 47 | 35.34 | 580 | 20.86 |
| Follow-up in days | Median (IQR) | 1223 (883-1593) |  | 1208 (879-1609) |  | 915 (878-1592) |  | 1223.5 (884-1621) |  | 1221 (881-1599) |  |
| Time to death | Median(IQR) | 185 (29-359) |  | 196.5 (8.75-726.5) |  | 948.5 (661.75-1235.25) |  | 709 (709-709) |  | 336 (10.5-596.25) |  |
| Length of hospital stay | Median(IQR) | 3 (3-5) |  | 3 (2-4) |  | 3 (2-4) |  | 3 (2-4) |  | 3 (2-4) |  |

**Figure 1.**
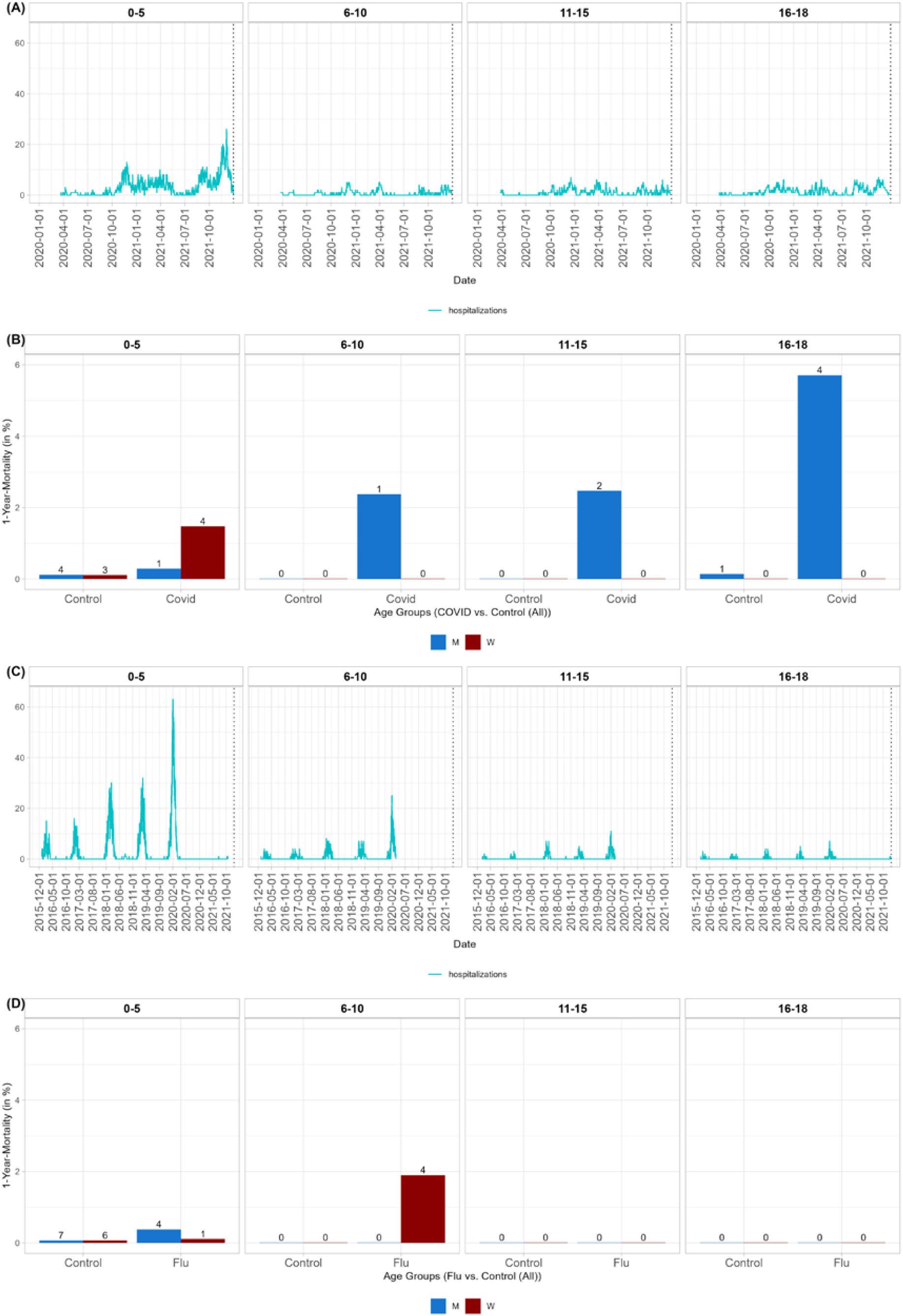
Time-line and 1-year mortality by age group. Timeline of hospitalization due to (A) COVID-19 in 2020 and 2021 (C) Influenza from 2015 to 2021 (hospitalizations per day in blue). 1-year mortality in age groups and by sex for (B) COVID-19-hospitalized (D) Influenza-hospitalized children and corresponding age-, sex- and region-matched controls.

We observed a greater disease burden based on observed medication groups for children hospitalized due to COVID-19 as compared to the general population. For almost all medication groups that were not underrepresented in the children cohort, a significant difference to the general population was observed (Table S3). Overall, polypharmacy was observed in 15.43 % of COVID-hospitalized children. Whereas for the youngest age group (0-5 years) only 7.55 % with polypharmacy were found, for all other age groups this value increased to more than 20% (Table 1). Similar results were found for the Influenza group (Table S4). However, polypharmacy was present in 20.86% of Influenza hospitalized children. Generally, over the investigated age groups, a larger percentage of polypharmacy was observed in Influenza as compared to COVID hospitalized children (Table 1). 70.5% of Influenza patients were aged 0 -5 and 4.8 % aged 16-18 as compared to 57.3% and 17.8% in the COVID-group respectively (Table 1).

### Time to hospital discharge

Overall, the median length of hospital stay was 3 days (IQR: 2-5) in the COVID-19 group. This did not vary over the age groups (Table 1). Age group and sex generally did not show a significant association with the probability for hospital discharge in the COVID-cohort, except in children aged 6-10, which showed a significant longer time to hospital discharge as compared to children aged 0-5 (p<0.001, Table S5, Figure S1 (A)). In the simple models, children with polypharmacy showed a significant longer time to hospital discharge (p=0.003, see Table S5, Figure S2 (A)). However, this effect did not remain significant in the multivariable model including all medication groups (Table S5). The multivariable model evaluated a significant longer time to hospital discharge for COVID-19 hospitalized children with prescribed Iron supplements, erythropoietic stimulating ages, Vitamin B12 and folic acid (p=0.002), Vitamin D and other Vitamin supplements (p<0.001), interferons and CSF (p<0.001) as well as antiepileptics (p<0.001) (Figure 2 (A) and Table S5).

**Figure 2.**
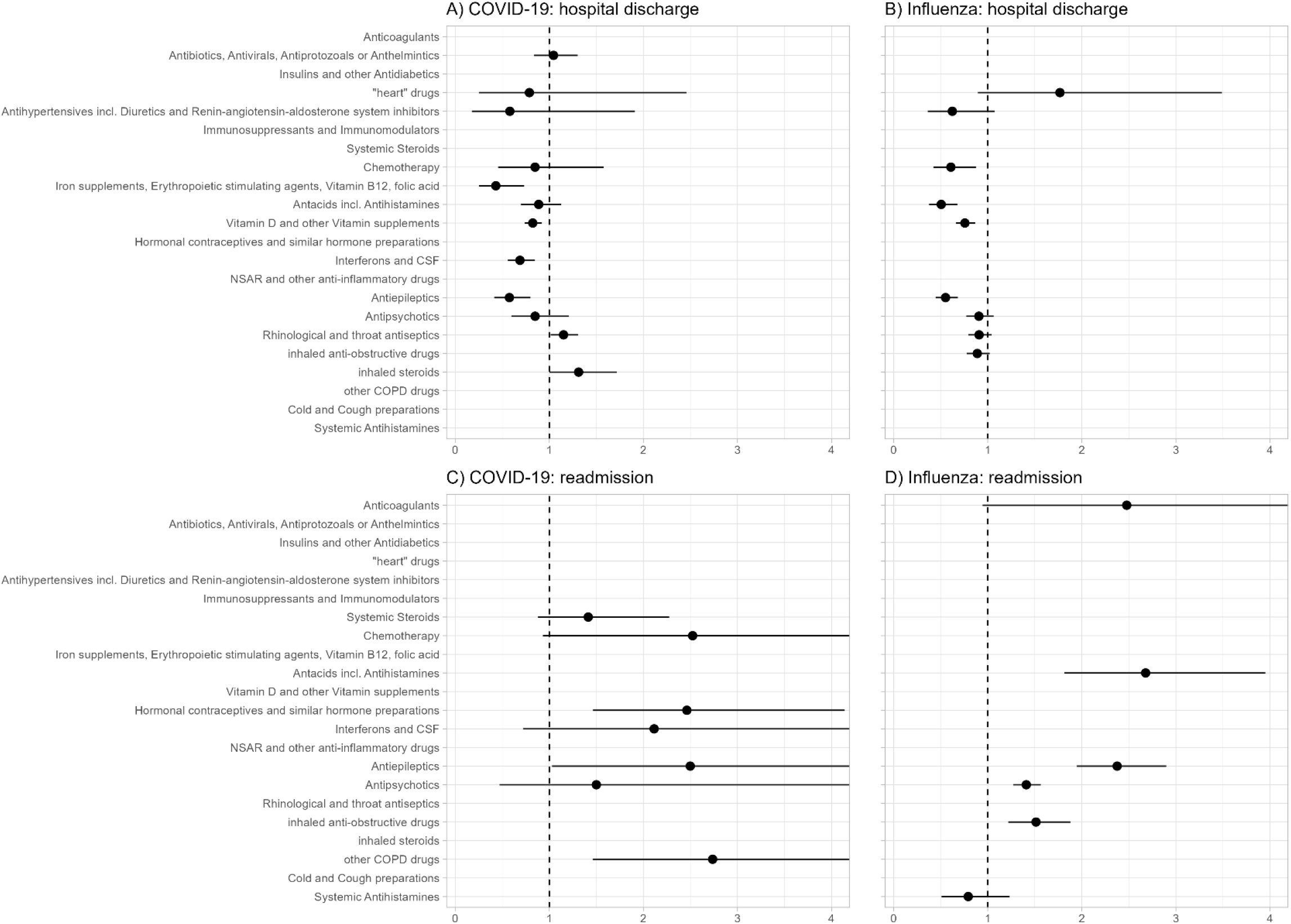
Hazard ratios and 95% confidence intervals for medication groups of the outcome hospital discharge (first row) and readmission (second row) after index COVID (first column) and Influenza (second column) hospitalisation for the patient groups. A hazard ratio >1 indicates larger probability of discharge (risk of readmission respectively) of patients receiving a drug in the corresponding medication group.

Within the Influenza group, the median length of hospital stay was 3 days (IQR: 2-4). Age groups 6-10 (p<0.001), 11-15 (p=0.012) and 16-18 (p<0.001) showed a significant lower probability for hospital discharge as compared to children aged 0-5 (Table S6, Figure S1 (B)). Polypharmacy showed a significant association with time to hospital discharge (p=0.003, Table S6, Figure S1 (B)). Children with polypharmacy showed a longer time to hospital discharge (p=0.003, Table S6, Figure S2 (B)). The multivariable model evaluated a significant longer time to hospital discharge for Influenza hospitalized children receiving Chemotherapy (p=0.008), Antacids incl. Antihistamines (p<0.001), Vitamin D and other Vitamin supplements (p<0.001) as well as antiepileptics (p<0.001) in the year before hospitalization (Figure 2 (B) and Table S6).

To compare the hospitalized Influenza and COVID-19 cohort we performed propensity score matching to account for potential confounders (Table 2). No significant result for the time to hospital discharge was found between children, hospitalized due to Influenza or COVID-19 (HR: 1.22 [95%-CI: 0.97-1.55], p=0.093, Figure 3 (A), Table S7).

**Table 2.**
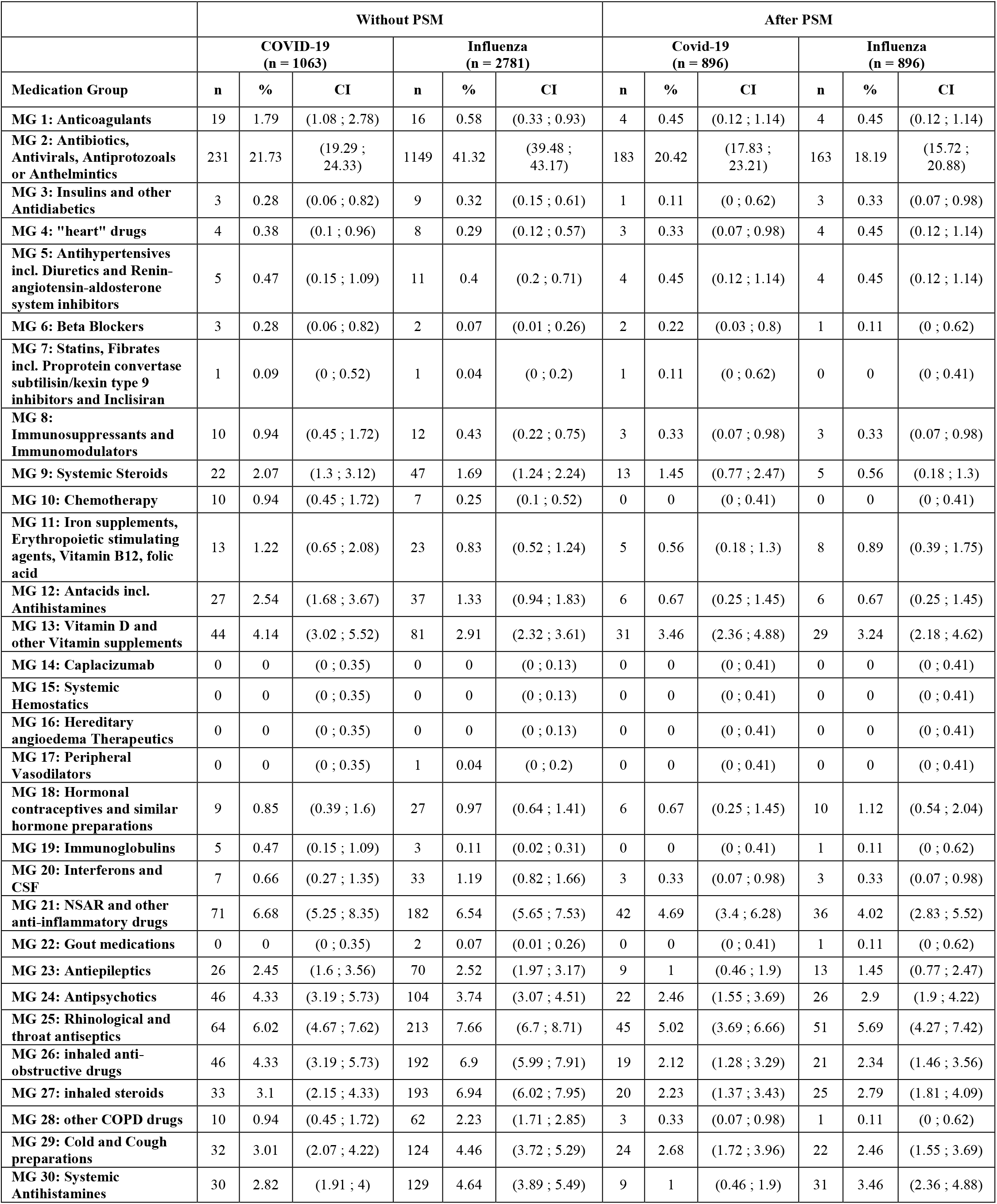
Medication Groups presented as number Yes, percentage and the corresponding 95%-CI observed in COVID-19 or Influenza hospitalized children in Austria before and after propensity score matching.

|  | Without PSM |  |  |  |  |  | After PSM |  |  |  |  |  |
| --- | --- | --- | --- | --- | --- | --- | --- | --- | --- | --- | --- | --- |
|  | COVID-19<br>(n = 1063) |  |  | Influenza<br>(n = 2781) |  |  | Covid-19<br>(n = 896) |  |  | Influenza<br>(n = 896) |  |  |
| Medication Group | n | % | CI | n | % | CI | n | % | CI | n | % | CI |
| MG 1: Anticoagulants | 19 | 1.79 | (1.08 ; 2.78) | 16 | 0.58 | (0.33 ; 0.93) | 4 | 0.45 | (0.12 ; 1.14) | 4 | 0.45 | (0.12 ; 1.14) |
| MG 2: Antibiotics, Antivirals, Antiprotozoals or Anthelmintics | 231 | 21.73 | (19.29 ; 24.33) | 1149 | 41.32 | (39.48 ; 43.17) | 183 | 20.42 | (17.83 ; 23.21) | 163 | 18.19 | (15.72 ; 20.88) |
| MG 3: Insulins and other Antidiabetics | 3 | 0.28 | (0.06 ; 0.82) | 9 | 0.32 | (0.15 ; 0.61) | 1 | 0.11 | (0 ; 0.62) | 3 | 0.33 | (0.07 ; 0.98) |
| MG 4: "heart" drugs | 4 | 0.38 | (0.1 ; 0.96) | 8 | 0.29 | (0.12 ; 0.57) | 3 | 0.33 | (0.07 ; 0.98) | 4 | 0.45 | (0.12 ; 1.14) |
| MG 5: Antihypertensives incl. Diuretics and Renin-angiotensin-aldosterone system inhibitors | 5 | 0.47 | (0.15 ; 1.09) | 11 | 0.4 | (0.2 ; 0.71) | 4 | 0.45 | (0.12 ; 1.14) | 4 | 0.45 | (0.12 ; 1.14) |
| MG 6: Beta Blockers | 3 | 0.28 | (0.06 ; 0.82) | 2 | 0.07 | (0.01 ; 0.26) | 2 | 0.22 | (0.03 ; 0.8) | 1 | 0.11 | (0 ; 0.62) |
| MG 7: Statins, Fibrates incl. Proprotein convertase subtilisin/kexin type 9 inhibitors and Inclisiran | 1 | 0.09 | (0 ; 0.52) | 1 | 0.04 | (0 ; 0.2) | 1 | 0.11 | (0 ; 0.62) | 0 | 0 | (0 ; 0.41) |
| MG 8: Immunosuppressants and Immunomodulators | 10 | 0.94 | (0.45 ; 1.72) | 12 | 0.43 | (0.22 ; 0.75) | 3 | 0.33 | (0.07 ; 0.98) | 3 | 0.33 | (0.07 ; 0.98) |
| MG 9: Systemic Steroids | 22 | 2.07 | (1.3 ; 3.12) | 47 | 1.69 | (1.24 ; 2.24) | 13 | 1.45 | (0.77 ; 2.47) | 5 | 0.56 | (0.18 ; 1.3) |
| MG 10: Chemotherapy | 10 | 0.94 | (0.45 ; 1.72) | 7 | 0.25 | (0.1 ; 0.52) | 0 | 0 | (0 ; 0.41) | 0 | 0 | (0 ; 0.41) |
| MG 11: Iron supplements, Erythropoietic stimulating agents, Vitamin B12, folic acid | 13 | 1.22 | (0.65 ; 2.08) | 23 | 0.83 | (0.52 ; 1.24) | 5 | 0.56 | (0.18 ; 1.3) | 8 | 0.89 | (0.39 ; 1.75) |
| MG 12: Antacids incl. Antihistamines | 27 | 2.54 | (1.68 ; 3.67) | 37 | 1.33 | (0.94 ; 1.83) | 6 | 0.67 | (0.25 ; 1.45) | 6 | 0.67 | (0.25 ; 1.45) |
| MG 13: Vitamin D and other Vitamin supplements | 44 | 4.14 | (3.02 ; 5.52) | 81 | 2.91 | (2.32 ; 3.61) | 31 | 3.46 | (2.36 ; 4.88) | 29 | 3.24 | (2.18 ; 4.62) |
| MG 14: Caplacizumab | 0 | 0 | (0 ; 0.35) | 0 | 0 | (0 ; 0.13) | 0 | 0 | (0 ; 0.41) | 0 | 0 | (0 ; 0.41) |
| MG 15: Systemic Hemostatics | 0 | 0 | (0 ; 0.35) | 0 | 0 | (0 ; 0.13) | 0 | 0 | (0 ; 0.41) | 0 | 0 | (0 ; 0.41) |
| MG 16: Hereditary angioedema Therapeutics | 0 | 0 | (0 ; 0.35) | 0 | 0 | (0 ; 0.13) | 0 | 0 | (0 ; 0.41) | 0 | 0 | (0 ; 0.41) |
| MG 17: Peripheral Vasodilators | 0 | 0 | (0 ; 0.35) | 1 | 0.04 | (0 ; 0.2) | 0 | 0 | (0 ; 0.41) | 0 | 0 | (0 ; 0.41) |
| MG 18: Hormonal contraceptives and similar hormone preparations | 9 | 0.85 | (0.39 ; 1.6) | 27 | 0.97 | (0.64 ; 1.41) | 6 | 0.67 | (0.25 ; 1.45) | 10 | 1.12 | (0.54 ; 2.04) |
| MG 19: Immunoglobulins | 5 | 0.47 | (0.15 ; 1.09) | 3 | 0.11 | (0.02 ; 0.31) | 0 | 0 | (0 ; 0.41) | 1 | 0.11 | (0 ; 0.62) |
| MG 20: Interferons and CSF | 7 | 0.66 | (0.27 ; 1.35) | 33 | 1.19 | (0.82 ; 1.66) | 3 | 0.33 | (0.07 ; 0.98) | 3 | 0.33 | (0.07 ; 0.98) |
| MG 21: NSAR and other anti-inflammatory drugs | 71 | 6.68 | (5.25 ; 8.35) | 182 | 6.54 | (5.65 ; 7.53) | 42 | 4.69 | (3.4 ; 6.28) | 36 | 4.02 | (2.83 ; 5.52) |
| MG 22: Gout medications | 0 | 0 | (0 ; 0.35) | 2 | 0.07 | (0.01 ; 0.26) | 0 | 0 | (0 ; 0.41) | 1 | 0.11 | (0 ; 0.62) |
| MG 23: Antiepileptics | 26 | 2.45 | (1.6 ; 3.56) | 70 | 2.52 | (1.97 ; 3.17) | 9 | 1 | (0.46 ; 1.9) | 13 | 1.45 | (0.77 ; 2.47) |
| MG 24: Antipsychotics | 46 | 4.33 | (3.19 ; 5.73) | 104 | 3.74 | (3.07 ; 4.51) | 22 | 2.46 | (1.55 ; 3.69) | 26 | 2.9 | (1.9 ; 4.22) |
| MG 25: Rhinological and throat antiseptics | 64 | 6.02 | (4.67 ; 7.62) | 213 | 7.66 | (6.7 ; 8.71) | 45 | 5.02 | (3.69 ; 6.66) | 51 | 5.69 | (4.27 ; 7.42) |
| MG 26: inhaled anti-obstructive drugs | 46 | 4.33 | (3.19 ; 5.73) | 192 | 6.9 | (5.99 ; 7.91) | 19 | 2.12 | (1.28 ; 3.29) | 21 | 2.34 | (1.46 ; 3.56) |
| MG 27: inhaled steroids | 33 | 3.1 | (2.15 ; 4.33) | 193 | 6.94 | (6.02 ; 7.95) | 20 | 2.23 | (1.37 ; 3.43) | 25 | 2.79 | (1.81 ; 4.09) |
| MG 28: other COPD drugs | 10 | 0.94 | (0.45 ; 1.72) | 62 | 2.23 | (1.71 ; 2.85) | 3 | 0.33 | (0.07 ; 0.98) | 1 | 0.11 | (0 ; 0.62) |
| MG 29: Cold and Cough preparations | 32 | 3.01 | (2.07 ; 4.22) | 124 | 4.46 | (3.72 ; 5.29) | 24 | 2.68 | (1.72 ; 3.96) | 22 | 2.46 | (1.55 ; 3.69) |
| MG 30: Systemic Antihistamines | 30 | 2.82 | (1.91 ; 4) | 129 | 4.64 | (3.89 ; 5.49) | 9 | 1 | (0.46 ; 1.9) | 31 | 3.46 | (2.36 ; 4.88) |

**Figure 3.**
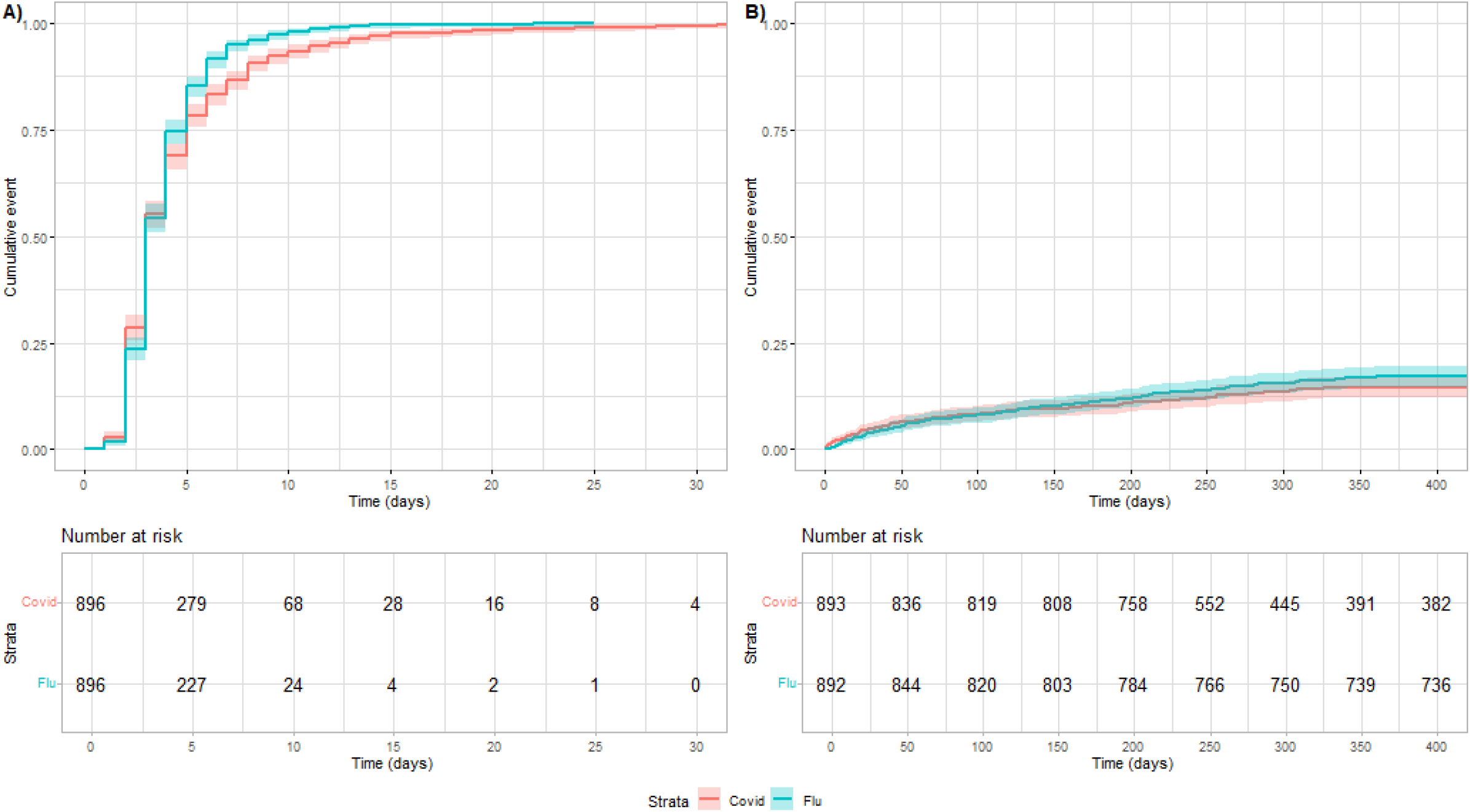
Estimated event probability and corresponding 95% confidence intervals for comparing propensity score matched COVID- and Influenza-patients for discharge (A) and readmission due to any reason (B).

### Readmission due to any reason

The probability of readmission within one year after hospitalizations was estimated with to be 14.5% (CI: 16.94-11.99) in the COVID-19 group and 17.06% (CI 19.49-14.55) in the Influenza group (Tables S8 and S9). Readmission rates were generally low, statistical models should therefore interpreted with care.

Within the COVID-19 and Influenza groups, children with polypharmacy showed a significant higher probability for readmission (both p<0.001, Table S10 and S11). Concerning medication groups, the multivariable model evaluated a significant higher risk for readmission for children with prescribed Hormonal contraceptives and similar hormone preparations (p<0.001) and other COPD-drugs (p=0.002) within the COVID-19 cohort (Table S10) as well as for children with prescribed chemotherapy, antacids incl. antihistamines, antiepileptics, antipsychotics and inhaled anti-obstructive drugs (all p<0.001) within the Influenza-cohort (Table S11).

A significant larger probability for readmission was found for children, hospitalized due to Influenza as compared to COVID-19 (HR: 1.23 CI: 1.03 – 1.47, p=0.0214, Figure 3 B, Table S12). However, although we performed propensity score matching and corrected for several medication groups in the statistical model we cannot rule out that Influenza hospitalized patients generally had a larger disease burden.

### Death

Death rates were very small in COVID-19 as well as in Influenza hospitalized children. In 2020 and 2021 a number of 10 children (0.94%) died during COVID-hospital stay. In contrast, in the years 2015 to 2021 a number of 6 children (0.22%) died during Influenza-hospital stay (p=0.004; Tables S13, S14). 7 out of the 10 children dying during COVID hospital stay and 5 out of the 6 children dying during Influenza hospital stay had prescribed drugs of at least 2 medication groups in the year before hospital stay (S15, S16). A trend towards a larger probability of all-cause death was observed for COVID-19 patients (p=0.009), however, due to the small number of events, no propensity score matching or correction for confounders could be performed. One-year mortality after hospitalizations was estimated with 1.13% (CI: 0.49-1.77) in the COVID and 0.32% (CI: 0.11-0.53) in the Influenza group. Note that the numbers in the respective age-, sex- and region-matched control groups were 0.08% (CI: 0.02-0.13) for COVID and 0.05% (CI: 0.02-0.07) for Influenza (see Figure 1 (B), (D), Tables S13-S16).

## Discussion

In this population based study we present data on children ≤18years of age, hospitalized due to COVID-19- or Influenza provided by the Austrian Health Insurance Funds to evaluate short- and long-term outcomes as well as their disease burden as compared to the general population.

A few studies have directly compared the respective burden of COVID-19 and influenza epidemics in children and the results are conflicting. Severe outcomes seem to be rare in both cohorts. Piroth et al ^(19)^ found that in children in-hospital mortality was higher in COVID patients but generally low death numbers were observed. They discussed that children with COVID-19 were more frequently obese or overweight, and more frequently had diabetes, hypertension, dyslipidaemia than patients with Influenza, whereas those with Influenza more frequently head heart failure, chronic respiratory disease, cirrhosis and deficiency anaemia. In a nationwide UK study, PICU admissions with severe COVID-19 in children were infrequent. In comparison with Influenza, severe infected children were older, more often Black or Asian, had a higher weight z-score, and higher deprivation index. Comorbidities, frequency of organ support, and length of stay were similar. ^(26)^

In a study in US children admitted to the ICU with Influenza or COVID-19 they observed, that children with Influenza were younger than those with COVID-19, less likely to be non-Hispanic Black or Hispanic, and less likely to have ≥1 underlying condition or be obese, and a shorter hospital stay. Despite differences in demographics and clinical characteristics of children with Influenza or COVID-19, the prevalence of life-threatening illness was similar between the 2 cohorts after adjusting for significant differences in demographics and clinical characteristics. Furthermore, they observed that adolescents (aged 13–17 years) and preschoolers (aged 24–59 months) had longer hospital and ICU stays with COVID-19 than with influenza.^(27).^ Laris-Gonzalez A. et al ^(28)^ concluded that comorbidities were frequent in both groups, but they were more common in patients with influenza. In-hospital mortality and the need for mechanical ventilation among symptomatic patients were similar between groups in the multivariate analysis.

A meta-analysis of 16 studies found adverse clinical outcomes (e.g. mortality and need for mechanical ventilation) to be similar between both COVID-19 and Influenza groups, but significantly higher inflammatory markers in COVID-19 patients.^(23)^ This systematic review also included several studies concluding that Influenza is more systematic or have a more severe clinical and laboratory course than COVID-19.^(23)^ Brigadoi et al ^(29)^ discussed that COVID-19 in children is clinically similar to other viral RTIs but is associated with a less severe infection course with a lower risk of being admitted, receiving respiratory support, needing antibiotic therapy and developing complications. In a Canadian study, among children aged 5-17 years, COVID-19 hospitalization rates were lower than in historical Influenza cohorts. The COVID-19 hospitalization rate among 0-4 years old, during Omicron, was higher than Influenza 2015/2016 and 2016/2017 and lower than 2009/2010 pandemic.^(30)^ In a cohort study of US children with COVID-19 or seasonal Influenza, there was no difference in hospitalization rates, intensive care unit admission rates, and mechanical ventilator use between the 2 groups. ^(31)^

Other investigations revealed a different picture. Tso et al ^(32)^ observed that for hospitalized children Omicron BA.2 appeared to be more neuropathogenic than Influenza and parainfluenza viruses. The relative risk of PICU admission was significantly higher for COVID-19 than Influenza. Similarly, another study showed that pediatric COVID-19 patients were older than seasonal Influenza patients and the intensive care unit admission rate was higher among COVID-19 patients. However, no intergroup differences in hospitalization or mortality rates, oxygen requirements, or hospital length of stay between children with COVID-19 and Influenza were observed.^(32)^ In a large meta-analysis of 455 studies the duration of hospitalization in COVID-19 patients (14 days) was longer than influenza type A (6.5 days) and influenza type B (6.7 days). Case fatality rate of hospitalized patients was higher in COVID-19 (6.5%) than in influenza type A and B (6% and 3%).^(34)^

In another analysis from the US, the annual COVID-19-associated hospitalization rate during 2020-2021 was higher among adolescents and similar or lower among children <12 years compared with influenza during the 3 seasons before the COVID-19 pandemic. A higher proportion with COVID-19 required ICU admission compared with Influenza. However, there was no difference in in-hospital mortality or length of hospital stay. The authors concluded that COVID-19 adds substantially to the existing burden of pediatric hospitalizations and severe outcomes.^(35)^

In our large population based cohort from Austria, children tended to be younger (but not statistically significant) in the Influenza group. Although both cohorts showed a higher disease burden based on their medication profiles as compared to the general population, for Influenza patients we observed a significant larger proportion of prescribed Antibiotics, Antivirals, Antiprotozoals or Antihelmintics, inhaled anti-obstructive drugs, inhaled steroids, other COPD drugs and systemic antihistamines whereas in the COVID cohort, a significant larger proportion of Anticoagulants, chemotherapy and Immunoglobulins were found. Influenza patients were more likely to have polypharmacy. After correction for several confounding variables, no significant difference between COVID and Influenza hospitalized children for time to hospital discharge was observed.

In-hospital and all-cause mortality was significantly larger in COVID-19 as compared to the Influenza hospitalized children, but the finding is limited to a small number of events. The risk of readmission was significantly higher in the Influenza group.

The main strength of this study is the use of a large, representative, real-world national database from the Austrian Insurance Fund providing detailed information on demographics and medication with a long-term follow-up. The retrospective design, however, may lead to bias due to unknown confounding factors. As in any observational research we cannot rule out unmeasured confounding, i.e. that Influenza hospitalized patients generally had a larger health burden. We tried to account for potential bias by including several confounding factors in the statistical models and performing propensity score matching to support meaningful comparisons. During COVID-19 pandemic, Influenza hospitalization were rare so that the comparison of Influenza and COVID groups in the same time period was not possible which may lead to bias. Furthermore, no information was available from the Austrian Insurance Fund on COVID-19 or Influenza vaccination or intensive care in hospitals. Vaccination for COVID-19 was first authorized for children at the end of 2021; therefore, COVID-19 vaccination it may not be an important factor for children hospitalized in 2020 or 2021. Last, the study includes COVID hospitalizations in 2020 and 2021 and may therefore not be generalized to Omicron-Variants.

In conclusion, in hospitalized COVID-19 and Influenza children, both groups represent a pediatric population with higher disease burden and a higher mortality risk as compared to the general population. Children hospitalized with Influenza are generally younger than children with COVID-19, are more likely to have polypharmacy and have a significantly higher risk of readmission than children with COVID-19. Time to hospital discharge did not significantly differ between cohorts. However, a general picture of COVID-19 being a milder disease compared to Influenza may not be supported since, although only few deaths were observed, death rates of COVID-19 hospitalized children seem to be higher. However, a low number of severe events may limit the findings.

## Data Availability

Data supporting the results presented in the study are available upon reasonable request to the corresponding author.

## Author Contributions

H.J.A., B.R., R.W., and A.C.G. were responsible for conceptualization. B.R., A.C.G., C.W., J.M., and H.J.A. conceived the study and curated the data. C.W. and A.C.G cleaned, analyzed, and verified the underlying data. C.W., A.C.G., H.J.A. and R.W. wrote the paper and visualized the data. H.J.A. provided funding. A.C.G, B.R., C.W., M.M., J.M., C.A., J.A., R.W., and H.J.A. commented on the paper, oversaw the analysis, and edited the final manuscript.

## Acknowledgements

The work was funded by the working group ARGE Ankersmit, Laboratory for Cardiac and Thoracic Diagnosis and Regeneration at the Department of Thoracic Surgery at the Medical University of Vienna. We thank the Pharmaco-economics Advisory Council of the Austrian Sickness Funds for providing the data, especially Ms. Karin Allmer for quality assurance of the database query and Mr. Ludwig Weissengruber for organizational support for data generation.

## References

1) Williamson EJ., Walker AJ., Bhaskaran K., Bacon S., Bate X., Morton CE., Curtis HJ., Mehrkar A., Evans D., Inglesby P., Cockburn J., McDonald H., MacKenna B., Tomlinson L., Douglas IJ., Rentsch CT., Mathur R., Wong AYS., Grieve R., Harrisonh D., Forbes H., Schultze A., Croker R., Parry J., Hester F., Harper S., Perera R., Evans SJW., Liam Smeeth, Goldacre B. (2020). Factors associated with COVID-19-related death using OpenSAFELY, Nature: 584: 430–436

2) Posch M., Bauer P., Posch A., Koenig F. (2020). Analysis of Austrian COVID-19 deaths by age and sex. Wiener Klinische Wochenschrift: 132: 685–689.

3) Bauer P., Brugger J., Koenig F., Posch M. (2021). An international comparison of age and sex dependency of COVID-19 deaths in 2020: a descriptive analysis. Scientific reports, 11: 19143.

4) Zajic P., Hiesmayr M., Bauer P., Baron DM., Gruber A., Joannidis M., Posch M., Metnitz PHG. (2023) Nationwide analysis of hospital admissions and outcomes of patients with SARS-CoV-2 infection in Austria in 2020 and 2021. Scientific Reports: 13: 8548

5) Zheng Z., Peng F., Xu B., Zhao J., Liu H., Peng J., Li, Q., Jiang C., Zhou Y., Liu S., Ye C., Zhang P., Xing Y., Guo H., Tang W. (2020) Risk factors of critical & mortal COVID-19 cases: A systematic literature review and meta-analysis. Journal of Infection: 81: e16–e25

6) Ssentongo P., Ssentongo AE., Heilbrunn ES., Ba DM., Chinchilli VM. (2020). Association of cardiovascular disease and 10 other pre-existing comorbidities with COVID-19 mortality: A systematic review and meta-analysis. Plos One, doi: 10.1371/journal.pone.0238215

7) Liu S., Cao Y., Du T., Zhi Y. (2020). Prevalence of Comorbid Asthma and Related Outcomes in COVID-19: A Systematic Review and Meta-Analysis. J Allerg Clin Immunol Pract: doi: 10.1016/j.jaip.2020.11.054

8) Graf AC., Reichardt B., Wagenlechner C., Krotka P. Traxler-Weidenauer D., Mildner M., Mascherbauer J., Aigner C., Auer J., Wendt R., Ankersmit HJ. (2024) Baseline drug treatments and long-term outcomes in COVID-19-hospitalized patients: results of the 2020 AUTCOV study. Pre-print medRxiv doi: 10.1101/2024.08.22.24312424

9) Ayoubkhani D., Khunti K., Nafilyan V., Maddox T, Humberstone B., Diamond I, Banerjee A (2021). Post-covid syndrome in individuals admitted to hospital with covid-19: a retrospective cohort study. British Journal of Medicine, 372: n693

10) Günster C, Busse R, Spoden M, et al. (2021) 6-month mortality and readmissions of hospitalized COVID-19 patients: A nationwide cohort study of 8,679 patients in Germany. PLoS ONE, 16 (8): e0255427.

11) Oseran AS., Song Y., Xu J., Dahareh IJ., Wadhera RK., Lemos JA., Das SR., Sun T., Yen RW., Kazi DS. (2023) Long term risk of death and readmission after hospital admission with covid-19 among older adults: retrospective cohort study. BMJ: 382: e076222

12) Renda G, Ricci F, Spinoni EG, et al. (2022) Predictors of Mortality and Cardiovascular Outcome at 6 Months after Hospitalization for COVID-19. J Clin Med., 11 (3): 729.

13) Orlando V., Coscioni E., Guarino I., Mucherino S., Perrella A., Trama U., Limongelli G., Menditto E (2021) Drug-utilisation profiles and COVID-19, Scientific Reports, 11: 8913

14) Ghasemi H, Darvishi N, Salari N, Hosseinian-Far A, Akbari H, Mohammadi M. (2022). Global prevalence of polypharmacy among the COVID-19 patients: a comprehensive systematic review and meta-analysis of observational studies. Trop Med Heal., 50 (1): 60.

15) Iloanusi S, Mgbere O, Essien EJ. (2021) Polypharmacy among COVID-19 patients: A systematic review. J Am Pharm Assoc., 61 (5): e14–e25.

16) Visser AGR, Winkens B, Schols JMGA, Janknegt R, Spaetgens B. (2023) The impact of polypharmacy on 30-day COVID-related mortality in nursing home residents: a multicenter retrospective cohort study. Eur Geriatr Med.,14 (1): 51–57.

17) Peterson E., Koopmans M., Go U., Hamer DH., Petrosillo N., Castelli F., Storgaard M., Kahalili S., Simonsen L. (2020) Comparing SARS-CoV-2 with SARS-CoV and influenza pandemics. Lancet Infect Dis, 20: e238–244

18) Zayet S., Kadiane-Oussou NJ., Lepiller Q., Zahra H., Royer PY., Toko L., Gendrin V., Klopfenstein T. (2020). Clinical features of COVID-19 and influenza: a comparative study on Nord Franche-Comte cluster. Microbes and Infection: 22: 481–488.

19) Piroth L., Cottenet J., Mariet AS., Bonniaud P., Blot M., Tubert-Bitter P., Puantin C. (2021) Comparison of the characteristics, morbidity and mortality of COVID-19 and seasonal influenza: a nationwide population-based retrospective cohort study. Lancet Respir Med, 9: 251 – 259

20) Dhoschak N., Singhal T., Kabra SK, Lodha R. (2020). Pathophysiology of COVID-19: Why Children Fare Better than Adults? Indian Journal of Pediatrics, 87 (7): 537–546.

21) Szepfalusi Z., Schmidthaler K., Sieber J., Kopanja S., Götzinger F., Schoof A., Hoz J., Willinger B., Makristathis A., Weseslindther L., Stiasny K., Bohle B., Krotka P., Graf A., Frischer T. (2021). Lessons from low seroprevalence of SARS-CoV-2 antibodies in schoolschildren: A cross-sectional study, Pediatr Allergy Immunol, 32(4): 762–770.

22) Nikolopoulu GB, and Maltezou HC (2022). COVID-19 in Children: Were do we Stand? Archives of Medical Research, 53, 1–8

23) Yang Y., Zheng Q., Yang L., Wu L. (2024). Comparison of inflammatory markers, coagulation indicators and outcomes between influenza and COVID-19 infection amongst children: A systematic review and meta-analyses. Heliyon, 10: e30391

24) Bundesministerium für Arbeit, Soziales, Gesundheit und Konsumentenschutz (2019). The Austrian Health Care System: https://broschuerenservice.sozialministerium.at/Home/Download?publicationId=328&attachmentNme=The_Austrian_Health_Care_System_EN_pdfUA.pdf

25) R Core Team (2021). R: A language and environment for statistical computing. R Foundation for Statistical Computing, Vienna, Austria. https://www.R-project.org

26) Kanthimathinathan HK., Buckley H., Lamming C., Davis P., Ramnarayan P., Feltbower R., Draper E.S. for the PICANet COVID-19 Study group (2021). Characteristics of Severe Acute Respiratory Syndrome Coronavirus-2 Infection and Comparison with Influenza in Children admitted to U.K. PICUs. Critical Care Explor, 3 (3) e0362

27) Halasa NB., Spieker AJ, Young CC, Olson SM; Newhams MM., Amarin JZ, Moffitt KL, Nakamura MM, Levy, ER, Soma VL, Talj R, Weiss SL, Fitzgerald JC, Mack EH., Maddux AB, Schuster JE, Coates BM, Hall MW, Schartz SP, Scharz AJ, Kong M., Spinella PC, Lofits LL, McLaughlin GW, Hobbs CV, Rowan CM, Bembea MM, Nofziger RA, Babbitt CJ, Bowens C, Flori HR, Gertz SJ, Zinter MS, Guliano JS, Hume JR, Cvijanovich NZ, Singh AR, Crandall HA, Thomas NJ, Cullimore ML, Patel MM, Randolphand AG for the Pediatric Intensive Care Influenza and Overcoming COVID-19 Investigators (2023). Life-Threatening Complications of Influenza vs. Coronavirus Disease 2019 in US Children, Clin Infect Dis., 76 (3): e280–e290.

28) Laris-Gonzales A., Aviles-Robles M., Dominguez-Barrera C., Parra-Ortega I., Sanches-Huerta JL., Ojeda-Diezbarroso K., Bonilla-Pellegrini S., Olivar-Lopez V., Chavez-Lopez A., and Jimenez-Juarez R. (2021). Influenza vs. COVID-19: Comaprison of Clinical Characteristics and Outcomes in Pediatric Patients in Mexico City. Frontiers in Pediatrics, 9: 676611

29) Brigadoi G., Demarin GC., Boracchini R., Pierantoni L., Rossin S., Barbieri E., Tirelli F., Cantarutti A., Tempo G., Giaquinto C, Lanari M., Da Dalt L., Dona D. (2024). Comparison between the Viral Illness Caused by SARS-CoV-2, Influenza Virus, Respiratory Syncytial Virus and Other Respiratory Viruses in Pediatrics, Viruses, 16(2), 199

30) Setayeshgar S., Wilton J., Sbihi H., Zandy M., Janjua N., Choi A., Smolina K. (2023). comparison of influenza and COVID-19 hospitalizations in British Columbia Canada: a population-based study. BMJ Open Respiratory Research, 10: e001567

31) Song X., Delaney M., Shah RK.,, Campos JM., Wessel DV., DeBlasi RL. (2020). comparison of Clinical Features of COVID-19 vs. Seasonal Influenza A and B in US Children. JAMA Network Open, 3 (9): e2020495

32) Tso WWY., Kwan MYW., Wang YL., Leung LK, Leung D., Chua GT., Ip P. Fong DYT., Wong WHS, Chang SHS., Chan JFW, Peiris M., Lau YL., Duque JSR. (2022). Severity of SARSCoV-2 Omicron BA.2 infection in unvaccinated hospitalized children: comparison to influenza and parainfluenza infections, Emerging Microbes & Infections, 11:1, 1742–1750

33) Siddiqui M, Gültekingil A., Bakirci O, Uslu N., Baskin E., (2021). Comparison of clinical features and laboratory findings of coronavirus disease 2019 and influenza A and B infections in children: a single-center study. Clin Exp Pediatr. 64 (7), 364–369

34) Pormohammad A., Ghorbani S., Khatami A., Razizadeh M.H. Alborzi E., Zarei M. Idrovo J-P., Turner R.J. (2021). Comparison of influenza type A and B with COVID-19: A global systematic review and meta-analysis on clinical laboratory radiographic findings. Rev Med. Virol; 32: e2179

35) Delahoy MJ., Ujamaa D., Taylor CA., Cummings C., Anglin O., Holstein R., Milucky J., O’Halloran A., Patel K., Pham H., Whitaker M., Reingold A., Chai S.J., Aldern N.B., Kawasaki B., Meek J., Yousey-Hindes K., Anderson EJ., Opendo KP., Weigel A., Teno K., Reeg L., Leegwater L., Lynfield R., McMahon M., Ropp S., Rudin D., Muse A., Spina N., Bennet N., Popham K., Billing LM., Shlitz E., Sutton M., Thomas A., Schaffner W., Talbot HK., Crossland MT., McCaffrey K., Hall AJ., Burns E., McMorrow M., Reed C., Havers F.P., Garg S. (2022). Comparison of influenzan and COVID-19 associated hospitalizations among children <18 years old in the United States – FluSurv-NET (October-April 2017-2021) and COVID-NET (October 2020 – September 2021). Clin Infect Dis. ciac38.

