## Supplemental Figures and Tables for "Short- and Long-term Outcomes of children hospitalized with COVID-19 or Influenza: results of the AUTCOV study"

Note that data regarding adults (patients and controls >18 years of age) of the AUTCOV study are published on medRxiv, see Graf AC., Reichardt B., Wagenlechner C., Krotka P., Traxler-Weidenauer D., Mildner M., Mascherbauer J., Aigner C., Auer J., Wendt R., Ankersmit HJ. "Baseline durg treatments and long-term outcomes in COVID-19 hospitalized patients: results of the 2020 AUTCOV study" (2024),
doi: https://doi.org/10.1101/2024.08.22.24312424
Definitions of Medications Groups and Outcomes presented in the corresponding supplement also available on medRxiv are therefore similar to them presented below. Presented definitions on medication groups are used for the entire AUTCOV study and its publications.

### Definitions of primary and secondary outcome

This study includes all patients aged 0 to 18 years who were hospitalized in Austria due to the main diagnosis COVID-19 (ICD-10 Codes U071, U072, U049) from 1 January 2020 to 31 December 2021 and Influenza (ICD-10 Codes J09, J100, J101, J108, J110, J111, J118, J10) from 30 December 2015 to 31 December 2021. The patient data were available from the Austrian Health Insurance Funds. The data set includes 1063 COVID-19 patients and 2781 Influenza patients.

For both groups (COVID and INFLUENZA), age-, sex-, and region-matched control groups (approximately 10 controls for each patient) consisted of individuals not hospitalized due to COVID-19 (in the years 2020 and 21) as well as Influenza (in the years 2015 – 2021) were randomly chosen from the population registered in the Austrian Health Insurance Fund, representing the Austrian population. Data on the control group were available from 1 year before the first matched patient was hospitalized until study cut-off. Death dates were available at study cut-off.

**Table S1. Definitions of primary and secondary outcomes**

| Outcome | Definition |
|---|---|
| **Hospital Discharge (primary outcome)** | Hospital discharge after first COVDI-19 or Influenza hospital admission. Time from COVID-19/Influenza hospital admission to hospital discharge. |
| **In-hospital death (secondary outcome)** | for patients: death during COVID-19/Influenza index hospital stay: YES/NO<br>for controls: death within the time of COVID-19/Influenza hospital stay of the age-, sex and region-matched COVID-19 patient: YES/NO |
| **All-cause mortality (secondary outcome)** | for patients: time from COVID-19/Influenza hospital admission to death or last follow-up<br>for controls: time from COVID-19/Influenza hospital admission of the age-, sex and region-matched COVID-19 patient to death or last follow-up |
| **Readmission due to any reason (secondary outcome)** | time from (alive) COVID-19/Influenza hospital discharge to the first hospital admission (due to any reason) after index hospital stay |

### Definitions of confounder

For the statistical models separately for COVID-19 and for Influenza patients, the confounder wave was defined as follows:
For COVID-19 patients, the confounder 'wave' indicates the period during which patients were hospitalised for COVID between January to December 2020, January to June 2021 or July to December 2021.
For influenza patients, the confounder 'wave' was specified for rehospitalisation in the categories January 2016 to June 2017, July 2017 to December 2019 and January 2019 to December 2021, and for discharge in the categories January to June 2016, July 2016 to June 2017, July 2017 to June 2018, July 2018 to June 2019, June 2019 to July 2020, July 2020 to June 2021 and July to December 2021.

ATC codes describing prescribed medication were available from the Austrian Health Insurance Funds 1 year before the hospitalisation due to COVID-19 or Influenza. ATC codes for medications before hospitalisation were summarized into medication groups (**Table S2**) using binary variables, which were set to 1 if the patient received at least one medicament from the corresponding medication group at least once in the year before COVID-19 or Influenza hospitalisation. Not all medication groups were considered in the statistical models due to underrepresentation.

**Table S2: Definitions of medication groups**

| Medication group | ATC codes |
|---|---|
| **Anticoagulants (MG1)** | B01AA01, B01AA02, B01AA03, B01AA04, B01AA07, B01AA08, B01AA09, B01AA10, B01AA11, B01AA12, B01AB01, B01AB02, B01AB04, B01AB05, B01AB06, B01AB07, B01AB08, B01AB09, B01AB10, B01AB11, B01AB12, B01AB51, B01AC01, B01AC02, B01AC03, B01AC04, B01AC05, B01AC06, B01AC07, B01AC08, B01AC09, B01AC10, B01AC11, B01AC13, B01AC14, B01AC15, B01AC16, B01AC17, B01AC18, B01AC19, B01AC21, B01AC22, B01AC23, B01AC24, B01AC25, B01AC27, B01AC30, B01AC56, B01AD01, B01AD02, B01AD03, B01AD04, B01AD05, B01AD06, B01AD07, B01AD08, B01AD09, B01AD10, B01AD11, B01AD12, B01AE01, B01AE02, B01AE03, B01AE04, B01AE05, B01AE06, B01AE07, B01AF01, B01AF02, B01AF03, B01AF04, B01AX01, B01AX04, B01AX05, B01AX06 |
| **Antibiotics, antivirals, antiprotozoals, or anthelmintics (MG2)** | A07AA01, A07AA02, A07AA03, A07AA04, A07AA05, A07AA06, A07AA07, A07AA08, A07AA09, A07AA10, A07AA11, A07AA12, A07AA13, A07AA51, A07AA54, A07AB02, A07AB03, A07AB04, A07AC01, J01AA01, J01AA02, J01AA03, J01AA04, J01AA05, J01AA06, J01AA07, J01AA08, J01AA09, J01AA10, J01AA11, J01AA12, J01AA13, J01AA14, J01AA15, J01AA20, J01AA56, J01BA01, J01BA02, J01BA52, J01CA01, J01CA02, J01CA03, J01CA04, J01CA05, J01CA06, J01CA07, J01CA08, J01CA09, J01CA10, J01CA11, J01CA12, J01CA13, J01CA14, J01CA15, J01CA16, J01CA17, J01CA18, J01CA19, J01CA20, J01CA51, J01CE01, J01CE02, J01CE03, J01CE04, J01CE05, J01CE06, J01CE07, J01CE08, J01CE09, J01CE10, J01CE30, J01CF01, J01CF02, J01CF03, J01CF04, J01CF05, J01CG01, J01CG02, J01CR01, J01CR02, J01CR03, J01CR04, J01CR05, J01CR50, J01DA01, J01DA02, J01DA03, J01DA04, J01DA05, J01DA06, J01DA07, J01DA08, J01DA09, J01DA10, J01DA11, J01DA12, J01DA13, J01DA14, J01DA15, J01DA16, J01DA17, J01DA18, J01DA19, J01DA21, J01DA22, J01DA23, J01DA24, J01DA25, J01DA26, J01DA27, J01DA30, J01DA31, J01DA32, J01DA33, J01DA34, J01DA35, J01DA36, J01DA37, J01DA38, J01DA39, J01DA40, J01DA41, J01DA42, J01DA63, J01DB01, J01DB02, J01DB03, J01DB04, J01DB05, J01DB06, J01DB07, J01DB08, J01DB09, J01DB10, J01DB11, J01DB12, J01DC01, J01DC02, J01DC03, J01DC04, J01DC05, J01DC06, J01DC07, J01DC08, J01DC09, J01DC10, J01DC11, J01DC12, J01DC13, J01DC14, J01DD01, J01DD02, J01DD03, J01DD04, J01DD05, J01DD06, J01DD07, J01DD08, J01DD09, J01DD10, J01DD11, J01DD12, J01DD13, J01DD14, J01DD15, J01DD16, J01DD18, J01DD52, J01DD54, J01DD62, J01DD63, J01DD64, J01DE01, J01DE02, J01DF01, J01DF02, J01DH02, J01DH03, J01DH04, J01DH06, J01DH51, J01DH52, J01DH55, J01DH56, J01DI01, J01DI02, J01DI04, J01DI54, J01EA01, J01EA02, J01EB01, J01EB02, J01EB03, J01EB04, J01EB05, J01EB06, J01EB07, J01EB08, J01EB20, J01EC01, J01EC02, J01EC03, J01EC20, J01ED01, J01ED02, J01ED03, J01ED04, J01ED05, J01ED06, J01ED07, J01ED08, J01ED09, J01ED20, J01EE01, J01EE02, J01EE03, J01EE04, J01EE05, J01EE06, J01EE07, J01FA01, J01FA02, J01FA03, J01FA05, J01FA06, J01FA07, J01FA08, J01FA09, J01FA10, J01FA11, J01FA12, J01FA13, J01FA14, J01FA15, J01FF01, J01FF02, J01FG01, J01FG02, J01GA01, J01GA02, J01GB01, J01GB03, J01GB04, J01GB05, J01GB06, J01GB07, J01GB08, J01GB09, J01GB10, J01GB11, J01GB12, J01GB13, J01GB14, J01MA01, J01MA02, J01MA03, J01MA04, J01MA05, J01MA06, J01MA07, J01MA08, J01MA09, J01MA10, J01MA11, J01MA12, J01MA13, J01MA14, J01MA15, J01MA16, J01MA17, J01MA18, J01MA19, J01MA21, J01MA22, J01MA23, J01MA24, J01MB01, J01MB02, J01MB03, J01MB04, J01MB05, J01MB06, J01MB07, J01RA01, J01RA02, J01RA03, J01RA04, J01XA01, J01XA02, J01XA03, J01XA04, J01XA05, J01XB01, J01XB02, J01XC01, J01XD01, |

| | |
|---|---|
| | J01XD02, J01XD03, J01XE01, J01XE02, J01XX01, J01XX02, J01XX03, J01XX04, J01XX05, J01XX06, J01XX07, J01XX08, J01XX09, J01XX11, J01XX12, J02AA01, J02AA02, J02AB01, J02AB02, J02AC01, J02AC02, J02AC03, J02AC04, J02AC05, J02AX01, J02AX04, J02AX05, J02AX06, J04AA01, J04AA02, J04AA03, J04AB01, J04AB02, J04AB03, J04AB04, J04AB05, J04AB06, J04AB30, J04AC01, J04AC51, J04AD01, J04AD02, J04AD03, J04AK01, J04AK02, J04AK03, J04AK04, J04AK05, J04AK06, J04AK07, J04AM01, J04AM02, J04AM03, J04AM04, J04AM05, J04AM06, J04AM07, J04AM08, J04BA01, J04BA02, J04BA03, P01AA01, P01AA02, P01AA04, P01AA05, P01AA52, P01AB01, P01AB02, P01AB03, P01AB04, P01AB05, P01AB06, P01AB07, P01AC01, P01AC02, P01AC03, P01AC04, P01AR01, P01AR02, P01AR03, P01AR53, P01AX01, P01AX02, P01AX04, P01AX05, P01AX06, P01AX07, P01AX08, P01AX09, P01AX10, P01AX11, P01AX52, P01BA01, P01BA02, P01BA03, P01BA06, P01BA07, P01BB01, P01BB02, P01BB51, P01BC01, P01BC02, P01BD01, P01BD51, P01BE01, P01BE02, P01BE03, P01BE04, P01BE05, P01BE52, P01BF01, P01BF02, P01BF03, P01BF04, P01BF05, P01BF06, P01BX01, P01CA02, P01CA03, P01CB01, P01CB02, P01CC01, P01CC02, P01CD01, P01CD02, P01CX01, P01CX02, P01CX03, P01CX04, P02BA01, P02BA02, P02BB01, P02BX01, P02BX02, P02BX03, P02BX04, P02CA01, P02CA02, P02CA03, P02CA04, P02CA05, P02CA06, P02CA51, P02CB01, P02CB02, P02CC01, P02CC02, P02CE01, P02CF01, P02CX01, P02CX02, P02CX03, P02DA01, P02DX01, P02DX02, P03AA01, P03AA02, P03AA03, P03AA04, P03AA05, P03AA54, P03AB01, P03AB02, P03AB51, P03AC01, P03AC02, P03AC03, P03AC04, P03AC51, P03AC52, P03AC53, P03AC54, P03AX01, P03AX02, P03AX03, P03AX04, P03AX05, P03BA01, P03BA02, P03BA03, P03BA04, P03BX01, P03BX02, P03BX03, P03BX04, P03BX05, P03BX06 |
| **Insulin and other antidiabetics (MG3)** | A10AB01, A10AB02, A10AB03, A10AB04, A10AB05, A10AB06, A10AB30, A10AC01, A10AC02, A10AC03, A10AC04, A10AC30, A10AD01, A10AD02, A10AD03, A10AD04, A10AD05, A10AD06, A10AD30, A10AE01, A10AE02, A10AE03, A10AE04, A10AE05, A10AE06, A10AE30, A10AE54, A10AE56, A10AF01, A10BA01, A10BA02, A10BA03, A10BB01, A10BB02, A10BB03, A10BB04, A10BB05, A10BB06, A10BB07, A10BB08, A10BB09, A10BB10, A10BB11, A10BB12, A10BB31, A10BC01, A10BD01, A10BD02, A10BD03, A10BD04, A10BD05, A10BD06, A10BD07, A10BD08, A10BD09, A10BD10, A10BD11, A10BD12, A10BD13, A10BD14, A10BD15, A10BD16, A10BD17, A10BD18,A10BD19, A10BD20, A10BD21, A10BD22, A10BD23, A10BD24, A10BD25, A10BD26, A10BF01, A10BF02, A10BF03, A10BG01, A10BG02, A10BG03, A10BG04, A10BH01, A10BH02, A10BH03, A10BH04, A10BH05, A10BH06, A10BH07, A10BH08, A10BH52, A10BJ01, A10BJ02, A10BJ03, A10BJ04, A10BJ05, A10BJ06, A10BK01, A10BK02, A10BK03, A10BK04, A10BK05, A10BK06, A10BK07, A10BX01, A10BX02, A10BX03, A10BX04, A10BX05, A10BX06, A10BX07, A10BX08, A10BX09, A10BX10, A10BX11, A10BX12, A10BX13, A10BX14, A10XA01 |
| **Heart drugs (MG4)** | C01AA01, C01AA02, C01AA03, C01AA04, C01AA05, C01AA06, C01AA07, C01AA08,C01AA09, C01AA52, C01AB01, C01AB51, C01AC01, C01AC03, C01AX02, C01BA01, C01BA02, C01BA03, C01BA04, C01BA05, C01BA08, C01BA12, C01BA51, C01BA71, C01BB01, C01BB02, C01BB03, C01BB04, C01BC03, C01BC04, C01BC07, C01BC08, C01BD01, C01BD02, C01BD03, C01BD04, C01BD05, C01BD06, C01BD07, C01BG01, C01BG07, C01BG11, C01CA01, C01CA02, C01CA03, C01CA04, C01CA05, C01CA06, C01CA07, C01CA08, C01CA09, C01CA10, C01CA11, C01CA12, C01CA13, C01CA14, C01CA15, C01CA16, C01CA17, C01CA18, C01CA19, C01CA21, C01CA22, C01CA23, C01CA24, C01CA25, C01CA26, C01CA30, C01CA51, C01CE01, C01CE02, C01CE03, C01CE04, C01CX06, C01CX07, C01CX08, C01CX09, C01DA02, C01DA04, C01DA05, C01DA07, C01DA08, C01DA09, C01DA13, C01DA14, C01DA20, C01DA38, C01DA52, C01DA54, C01DA55, C01DA57, C01DA58, C01DA59, C01DA63, C01DA70, C01DB01, C01DX01, C01DX02, C01DX03, C01DX04, C01DX05, C01DX06, C01DX07, C01DX08, C01DX09, C01DX10, C01DX11, C01DX12, C01DX13, C01DX14, C01DX15, C01DX16, C01DX18, C01DX19, C01DX51, C01DX52, C01DX53, C01DX54, C01EA01, C01EB02, C01EB03, C01EB04, C01EB05, C01EB06, C01EB07, C01EB09, C01EB10, C01EB11, C01EB12, C01EB13, C01EB15, C01EB16, C01EB17, C01EB18, C01EB19, C01EB21 |
| **Antihypertensives, including diuretics and renin-angiotensin-aldosterone system inhibitors (MG5)** | C02AA01, C02AA02, C02AA03, C02AA04, C02AA05, C02AA06, C02AA07, C02AA52, C02AA53, C02AA57, C02AB01, C02AB02, C02AC01, C02AC02, C02AC04, C02AC05, C02AC06, C02BA01, C02BB01, C02CA01, C02CA02, C02CA03, C02CA04, C02CA06, C02CA08, C02CC01, C02CC02, C02CC03, C02CC04, C02CC05, C02CC06, C02CC07, C02DA01, C02DB01, C02DB02, C02DB03, C02DB04, C02DC01, C02DD01, C02DG01, C02KA01, C02KB01, C02KC01, C02KD01, C02KX01, C02KX02, C02KX03, C02KX04, C02KX05, C02KX52, C02LA01, C02LA02, C02LA03, C02LA04, C02LA07, C02LA08, C02LA09, C02LA50, C02LA51, C02LA52, C02LA71, C02LB01, C02LC01, C02LC05, C02LC51, C02LE01, C02LF01, C02LG01, C02LG02, C02LG03, C02LG51, C02LG73, C02LK01, C02LL01, C02LX01, C03AA01, C03AA02, C03AA03, C03AA04, C03AA05, C03AA06, C03AA07, C03AA08, C03AA09, C03AA13, C03AB01, C03AB02, C03AB03, C03AB04, C03AB05, C03AB06, C03AB07, C03AB08, C03AB09, C03AH01, C03AH02, C03AX01, C03BA02, C03BA03, C03BA04, C03BA05, C03BA07, C03BA08, C03BA09, C03BA10, C03BA11, C03BA12, C03BA13, C03BA82, C03BB02, C03BB03, C03BB04, |

| | |
|---|---|
| | C03BB05, C03BB07, C03BC01, C03BD01, C03BX03, C03CA01, C03CA02, C03CA03, C03CA04, C03CB01, C03CB02, C03CC01, C03CC02, C03CD01, C03CX01, C03DA01, C03DA02, C03DA03, C03DA04, C03DB01, C03DB02, C03EA01, C03EA02, C03EA03, C03EA04, C03EA05, C03EA06, C03EA07, C03EA12, C03EA13, C03EA14, C03EB01, C03EB02, C08CA01, C08CA02, C08CA03, C08CA04, C08CA05, C08CA06, C08CA07, C08CA08, C08CA09, C08CA10, C08CA11, C08CA12, C08CA13, C08CA14, C08CA15, C08CA16, C08CA51, C08CA55, C08CX01, C08DA01, C08DA02, C08DA51, C08DB01, C08EA01, C08EA02, C08EX01, C08EX02, C08GA01, C08GA02, C09AA01, C09AA02, C09AA03, C09AA04, C09AA05, C09AA06, C09AA07, C09AA08, C09AA09, C09AA10, C09AA11, C09AA12, C09AA13, C09AA14, C09AA15, C09AA16, C09BA01, C09BA02, C09BA03, C09BA04, C09BA05, C09BA06, C09BA07, C09BA08, C09BA09, C09BA12, C09BA13, C09BA15, C09BB02, C09BB03, C09BB04, C09BB05, C09BB06, C09BB07, C09BB10, C09BB12, C09BX01, C09BX03, C09BX04, C09BX05, C09CA01, C09CA02, C09CA03, C09CA04, C09CA05, C09CA06, C09CA07, C09CA08, C09CA09, C09DA01, C09DA02, C09DA03, C09DA04, C09DA06, C09DA07, C09DA08, C09DA10, C09DB01, C09DB02, C09DB04, C09DB05, C09DB06, C09DB07, C09DB09, C09DX01, C09DX02, C09DX03, C09DX04, C09DX05, C09DX06, C09DX07, C09XA01, C09XA02, C09XA52, C09XA53, C09XA54 |
| **Beta-blockers (MG6)** | C07AA01, C07AA02, C07AA03, C07AA05, C07AA06, C07AA07, C07AA12, C07AA14, C07AA15, C07AA16, C07AA17, C07AA19, C07AA23, C07AA27, C07AA57, C07AB01, C07AB02, C07AB03, C07AB04, C07AB05, C07AB06, C07AB07, C07AB08, C07AB09, C07AB10, C07AB11, C07AB12, C07AB13, C07AB14, C07AB52, C07AG01, C07AG02, C07BA02, C07BA05, C07BA06, C07BA07, C07BA12, C07BA68, C07BB02, C07BB03, C07BB04, C07BB06, C07BB07, C07BB12, C07BB52, C07BG01, C07CA02, C07CA03, C07CA17, C07CA23, C07CB02, C07CB03, C07CB53, C07CG01, C07DA06, C07DB01, C07FA05, C07FB02, C07FB03, C07FB07, C07FB13, C07FX01, C07FX02, C07FX03, C07FX04, C07FX05, C07FX06 |
| **Statins, fibrates, including proprotein convertase subtilisin/kexin type 9 inhibitors and inclisiran (MG7)** | C10AA01, C10AA02, C10AA03, C10AA04, C10AA05, C10AA06, C10AA07, C10AA08, C10AA51, C10AA52, C10AA53, C10AA55, C10AB01, C10AB02, C10AB03, C10AB04, C10AB05, C10AB06, C10AB07, C10AB08, C10AB09, C10AB10, C10AB11, C10AX06, C10AX07, C10AX08, C10AX09, C10AX10, C10AX11, C10AX12, C10AX13, C10AX14, C10AX15, C10AX16, C10BA01, C10BA02, C10BA03, C10BA04, C10BA05, C10BA06, C10BA07, C10BA08, C10BA09, C10BA10, C10BX01, C10BX02, C10BX03, C10BX04, C10BX05, C10BX06, C10BX07, C10BX08, C10BX09, C10BX10, C10BX11, C10BX12, C10BX13, C10BX14, C10BX15, C10BX16, C10BX17, C10BX18 |
| **Immunosuppressants and immunomodulators (MG8)** | D11AH05, L04AA01, L04AA02, L04AA03, L04AA04, L04AA05, L04AA06, L04AA08, L04AA09, L04AA10, L04AA11, L04AA12, L04AA13, L04AA14, L04AA15, L04AA16, L04AA17, L04AA18, L04AA19, L04AA21, L04AA22, L04AA23, L04AA24, L04AA25, L04AA26, L04AA27, L04AA28, L04AA29, L04AA31, L04AA32, L04AA33, L04AA34, L04AA36, L04AA37, L04AA38, L04AA39, L04AA40, L04AA41, L04AA42, L04AA43, L04AA44, L04AA45, L04AA46, L04AB01, L04AB02, L04AB03, L04AB04, L04AB05, L04AB06, L04AB07, L04AC01, L04AC02, L04AC03, L04AC04, L04AC05, L04AC06, L04AC07, L04AC08, L04AC09, L04AC10, L04AC11, L04AC12, L04AC13, L04AC14, L04AC15, L04AC16, L04AC17, L04AC18, L04AC19, L04AD01, L04AD02, L04AD03, L04AX01, L04AX02, L04AX03, L04AX04, L04AX05, L04AX06, L04AX07, L04AX08 |
| **Systemic steroids (MG9)** | H02AA01, H02AA02, H02AA03, H02AB01, H02AB02, H02AB03, H02AB04, H02AB05, H02AB06, H02AB07, H02AB08, H02AB09, H02AB10, H02AB11, H02AB12, H02AB13, H02AB14, H02AB15, H02AB17, H02BX01, H02CA01, H02CA02, H02CA03 |
| **Chemotherapy (MG10)** | L01AA01, L01AA02, L01AA03, L01AA05, L01AA06, L01AA07, L01AA08, L01AA09, L01AB01, L01AB02, L01AB03, L01AC01, L01AC02, L01AC03, L01AD01, L01AD02, L01AD03, L01AD04, L01AD05, L01AD06, L01AD07, L01AD08, L01AG01, L01AX01, L01AX02, L01AX03, L01AX04, L01BA01, L01BA03, L01BA04, L01BB02, L01BB03, L01BB04, L01BB05, L01BB06, L01BB07, L01BC01, L01BC02, L01BC03, L01BC04, L01BC05, L01BC06, L01BC07, L01BC08, L01BC09, L01BC52, L01BC53, L01BC59, L01CA01, L01CA02, L01CA03, L01CA04, L01CA05, L01CB01, L01CB02, L01CC01, L01CD01, L01CD02, L01CD03, L01CD04, L01CE01, L01CE02, L01CE03, L01CE04, L01CX01, L01DA01, L01DB01, L01DB02, L01DB03, L01DB04, L01DB05, L01DB06, L01DB07, L01DB08, L01DB09, L01DB10, L01DB11, L01DC01, L01DC02, L01DC03, L01DC04, L01EA01, L01EA02, L01EA03, L01EA04, L01EA05, L01EB01, L01EB02, L01EB03, L01EB04, L01EB05, L01EB07, L01EB08, L01EC01, L01EC02, L01EC03, L01ED01, L01ED02, L01ED03, L01ED04, L01ED05, L01EE01, L01EE02, L01EE03, L01EF01, L01EF02, L01EF03, L01EG01, L01EG02, L01EH01, L01EH02, L01EJ01, L01EJ02, L01EK01, L01EK03, L01EL01, L01EL02, L01EM01, L01EM02, L01EM03, L01EX01, L01EX02, L01EX03, L01EX04, L01EX05, L01EX07, L01EX08, L01EX09, L01EX10, L01EX11, L01EX12, L01EX13, L01EX14, L01XA01, L01XA02, L01XA03, L01XA04, L01XA05, L01XB01, L01XC01, L01XC02, L01XC03, L01XC04, L01XC05, L01XC06, L01XC07, L01XC08, L01XC09, L01XC10, L01XC11, L01XC12, L01XC13, L01XC14, L01XC15, L01XC16, L01XC17, L01XC18, L01XC19, L01XC21, L01XC22, L01XC23, L01XC24, L01XC25, L01XC26, L01XC27, L01XC28, L01XC29, L01XC31, L01XC32, L01XC33, L01XC34, L01XC35, L01XC36, L01XC37, L01XC38, L01XC39, L01XC40, L01XC41, L01XD01, L01XD02, L01XD03, L01XD04, L01XD05, L01XD06, |

| | |
|---|---|
| | L01XD07, L01XE01, L01XE02, L01XE03, L01XE04, L01XE05, L01XE06, L01XE07, L01XE08, L01XE09, L01XE10, L01XE11, L01XE12, L01XE13, L01XE14, L01XE15, L01XE16, L01XE17, L01XE18, L01XE21, L01XE23, L01XE24, L01XE25, L01XE26, L01XE27, L01XE28, L01XE29, L01XE31, L01XE33, L01XE34, L01XE35, L01XE36, L01XE37, L01XE38, L01XE39, L01XE41, L01XE42, L01XE43, L01XE44, L01XE45, L01XE46, L01XE47, L01XE48, L01XE50, L01XE51, L01XE52, L01XE53, L01XE54, L01XE56, L01XE57, L01XF01, L01XF02, L01XF03, L01XG01, L01XG02, L01XG03, L01XH01, L01XH02, L01XH03, L01XH05, L01XJ01, L01XJ02, L01XJ03, L01XK01, L01XK02, L01XK03, L01XK04, L01XX01, L01XX02, L01XX03, L01XX05, L01XX07, L01XX08, L01XX09, L01XX10, L01XX11, L01XX14, L01XX16, L01XX17, L01XX18, L01XX19, L01XX22, L01XX23, L01XX24, L01XX25, L01XX27, L01XX28, L01XX29, L01XX31, L01XX32, L01XX33, L01XX34, L01XX35, L01XX36, L01XX37, L01XX38, L01XX39, L01XX40, L01XX41, L01XX42, L01XX43, L01XX44, L01XX45, L01XX46, L01XX47, L01XX48, L01XX50, L01XX51, L01XX52, L01XX53, L01XX54, L01XX55, L01XX56, L01XX57, L01XX58, L01XX59, L01XX60, L01XX61, L01XX62, L01XX63, L01XX64, L01XX65, L01XX66, L01XX67, L01XX68, L01XX70, L01XX71, L01XY01, L01XY02, L02AA01, L02AA02, L02AA03, L02AA04, L02AB01, L02AB02, L02AB03, L02AE01, L02AE02, L02AE03, L02AE04, L02AE05, L02AE51, L02BA01, L02BA02, L02BA03, L02BB01, L02BB02, L02BB03, L02BB04, L02BB05, L02BB06, L02BG01, L02BG02, L02BG03, L02BG04, L02BG05, L02BG06, L02BX01, L02BX02, L02BX03 |
| **Iron supplements, erythropoietic stimulating agents, vitamin B12, folic acid (MG11)** | B03AA01, B03AA02, B03AA03, B03AA04, B03AA05, B03AA06, B03AA07, B03AA08, B03AA09, B03AA10, B03AA11, B03AA12, B03AB01, B03AB02, B03AB03, B03AB04, B03AB05, B03AB06, B03AB07, B03AB08, B03AB09, B03AB10, B03AC01, B03AC02, B03AC03, B03AC05, B03AC06, B03AC07, B03AD01, B03AD02, B03AD03, B03AD04, B03AD05, B03AE01, B03AE02, B03AE03, B03AE04, B03AE10, B03BA01, B03BA02, B03BA03, B03BA04, B03BA05, B03BA51, B03BA53, B03BB01, B03BB51, B03XA01, B03XA02, B03XA03, B03XA05, B03XA06 |
| **Antacids, including antihistamines (MG12)** | A02BB01, A02BB02, A02BC01, A02BC02, A02BC03, A02BC04, A02BC05, A02BC06, A02BC08 |
| **Vitamin D and other vitamin supplements (MG13)** | A11CA01, A11CA02, A11CC01, A11CC02, A11CC03, A11CC04, A11CC05, A11CC06, A11CC07, A11CC20, A11CC55, A11DA01, A11DA02, A11DA03, A11GA01, A11GB01, A11HA01, A11HA02, A11HA03, A11HA04, A11HA05, A11HA06, A11HA07, A11HA08, A11HA30, A11HA31, A11HA32 |
| **Caplacizumab (MG14)** | B01AX07 |
| **Systemic hemostatics (MG15)** | B02BX01, B02BX02, B02BX03, B02BX04, B02BX05, B02BX06, B02BX07, B02BX08, B02BX09 |
| **Hereditary angioedema therapeutics (MG16)** | B06AC01, B06AC02, B06AC03, B06AC04, B06AC05 |
| **Peripheral vasodilators (MG17)** | C04AA01, C04AA02, C04AA31, C04AB01, C04AB02, C04AC01, C04AC02, C04AC03, C04AC07, C04AD01, C04AD02, C04AD03, C04AD04, C04AE01, C04AE02, C04AE04, C04AE51, C04AE54, C04AF01, C04AX01, C04AX02, C04AX07, C04AX10, C04AX11, C04AX13, C04AX17, C04AX19, C04AX20, C04AX21, C04AX23, C04AX24, C04AX26, C04AX27, C04AX28, C04AX30, C04AX32 |
| **Hormonal contraceptives and similar hormone preparations (MG18)** | G03AA01, G03AA02, G03AA03, G03AA04, G03AA05, G03AA06, G03AA07, G03AA08, G03AA09, G03AA10, G03AA11, G03AA12, G03AA13, G03AA14, G03AA15, G03AA16, G03AA17, G03AB01, G03AB02, G03AB03, G03AB04, G03AB05, G03AB06, G03AB07, G03AB08, G03AB09, G03AC01, G03AC02, G03AC03, G03AC04, G03AC05, G03AC06, G03AC07, G03AC08, G03AC09, G03AC10, G03AD01, G03AD02, G03BA01, G03BA02, G03BA03, G03BB01, G03BB02, G03CA01, G03CA03, G03CA04, G03CA06, G03CA07, G03CA09, G03CA53, G03CA57, G03CB01, G03CB02, G03CB03, G03CB04, G03CC02, G03CC03, G03CC04, G03CC05, G03CC06, G03CC07, G03CX01, G03DA01, G03DA02, G03DA03, G03DA04, G03DB01, G03DB02, G03DB03, G03DB04, G03DB05, G03DB06, G03DB07, G03DB08, G03DC01, G03DC02, G03DC03, G03DC04, G03DC05, G03DC06, G03DC31, G03EA01, G03EA02, G03EA03, G03EK01, G03FA01, G03FA02, G03FA03, G03FA04, G03FA05, G03FA06, G03FA07, G03FA08, G03FA09, G03FA10, G03FA11, G03FA12, G03FA13, G03FA14, G03FA15, G03FA16, G03FA17, G03FB01, G03FB02, G03FB03, G03FB04, G03FB05, G03FB06, G03FB07, G03FB08, G03FB09, G03FB10, G03FB11, G03GA01, G03GA02, G03GA03, G03GA04, G03GA05, G03GA06, G03GA07, G03GA08, G03GA09, G03GA10, G03GA30, G03GB01, G03GB02, G03GB03, G03HA01, G03HB01, G03XA01, G03XA02, G03XB01, G03XB02, G03XC01, G03XC02, G03XC03, G03XX01 |
| **Immunoglobulins (MG19)** | J06BA01, J06BA02 |
| **Interferons and CSF (MG20)** | L03AA02, L03AA03, L03AA09, L03AA10, L03AA12, L03AA13, L03AA14, L03AA16, L03AA17, L03AB01, L03AB02, L03AB03, L03AB04, L03AB05, L03AB06, L03AB07, L03AB08, L03AB09, L03AB10, L03AB11, L03AB12, L03AB13, L03AB15, L03AB60, L03AB61, L03AC01, L03AC02, L03AX01, L03AX02, L03AX03, L03AX04, L03AX05, L03AX07, L03AX08, L03AX09, L03AX10, L03AX11, L03AX12, L03AX13, L03AX14, L03AX15, L03AX16, L03AX17, L03AX21 |

| | |
|---|---|
| **NSAID and other anti-inflammatory drugs (MG21)** | M01AA01, M01AA02, M01AA03, M01AA05, M01AA06, M01AB01, M01AB02, M01AB03, M01AB04, M01AB05, M01AB06, M01AB07, M01AB08, M01AB09, M01AB10, M01AB11, M01AB12, M01AB13, M01AB14, M01AB15, M01AB16, M01AB17, M01AB51, M01AB55, M01AC01, M01AC02, M01AC04, M01AC05, M01AC06, M01AC56, M01AE01, M01AE02, M01AE03, M01AE04, M01AE05, M01AE06, M01AE07, M01AE08, M01AE09, M01AE10, M01AE11, M01AE12, M01AE13, M01AE14, M01AE15, M01AE16, M01AE17, M01AE18, M01AE51, M01AE52, M01AE53, M01AE56, M01AG01, M01AG02, M01AG03, M01AG04, M01AH01, M01AH02, M01AH03, M01AH04, M01AH05, M01AH06, M01AH07, M01AX01, M01AX02, M01AX04, M01AX05, M01AX07, M01AX12, M01AX13, M01AX14, M01AX17, M01AX18, M01AX21, M01AX22, M01AX23, M01AX24, M01AX25, M01AX26, M01AX68, M01BA01, M01BA02, M01BA03, M01CA03, M01CB01, M01CB02, M01CB03, M01CB04, M01CB05, M01CC01, M01CC02 |
| **Gout medications (MG22)** | M04AA01, M04AA02, M04AA03, M04AA51, M04AB01, M04AB02, M04AB03, M04AB04, M04AB05, M04AC01, M04AC02, M04AX01 |
| **Antiepileptics (MG23)** | N03AA01, N03AA02, N03AA03, N03AA04, N03AA30, N03AB01, N03AB02, N03AB03, N03AB04, N03AB05, N03AB52, N03AB54, N03AC01, N03AC02, N03AC03, N03AD01, N03AD02, N03AD03, N03AD51, N03AE01, N03AF01, N03AF02, N03AF03, N03AF04, N03AG01, N03AG02, N03AG03, N03AG04, N03AG05, N03AG06, N03AX03, N03AX07, N03AX09, N03AX10, N03AX11, N03AX12, N03AX13, N03AX14, N03AX15, N03AX16, N03AX17, N03AX18, N03AX21, N03AX22, N03AX23, N03AX24, N03AX30 |
| **Antipsychotics (MG24)** | N05AA01, N05AA02, N05AA03, N05AA04, N05AA05, N05AA06, N05AA07, N05AB01, N05AB02, N05AB03, N05AB04, N05AB05, N05AB06, N05AB07, N05AB08, N05AB09, N05AB10, N05AC01, N05AC02, N05AC03, N05AC04, N05AD01, N05AD02, N05AD03, N05AD04, N05AD05, N05AD06, N05AD07, N05AD08, N05AD09, N05AE01, N05AE02, N05AE03, N05AE04, N05AF01, N05AF02, N05AF03, N05AF04, N05AF05, N05AG01, N05AG02, N05AG03, N05AH01, N05AH02, N05AH03, N05AH04, N05AH05, N05AH06, N05AK01, N05AL01, N05AL02, N05AL03, N05AL04, N05AL05, N05AL06, N05AL07, N05AN01, N05AX07, N05AX08, N05AX09, N05AX10, N05AX11, N05AX12, N05AX13, N05AX14, N05AX15, N05BA01, N05BA02, N05BA03, N05BA04, N05BA05, N05BA06, N05BA07, N05BA08, N05BA09, N05BA10, N05BA11, N05BA12, N05BA13, N05BA14, N05BA15, N05BA16, N05BA17, N05BA18, N05BA19, N05BA21, N05BA22, N05BA23, N05BA56, N05BB01, N05BB02, N05BB51, N05BC01, N05BC03, N05BC04, N05BC51, N05BD01, N05BE01, N05BX01, N05BX02, N05BX03, N05BX05, N05CA01, N05CA02, N05CA03, N05CA04, N05CA05, N05CA06, N05CA07, N05CA08, N05CA09, N05CA10, N05CA11, N05CA12, N05CA15, N05CA16, N05CA19, N05CA20, N05CA21, N05CA22, N05CB01, N05CB02, N05CC01, N05CC02, N05CC03, N05CC04, N05CC05, N05CD01, N05CD02, N05CD03, N05CD04, N05CD05, N05CD06, N05CD07, N05CD08, N05CD09, N05CD10, N05CD11, N05CD12, N05CD13, N05CD14, N05CD15, N05CE01, N05CE02, N05CE03, N05CF01, N05CF02, N05CF03, N05CF04, N05CH01, N05CH02, N05CM01, N05CM02, N05CM03, N05CM04, N05CM05, N05CM06, N05CM07, N05CM08, N05CM09, N05CM10, N05CM11, N05CM12, N05CM13, N05CM15, N05CM16, N05CM17, N05CM18, N05CM19, N05CX01, N05CX02, N05CX03, N05CX04, N05CX05, N05CX06, N06AA01, N06AA02, N06AA03, N06AA04, N06AA05, N06AA06, N06AA07, N06AA08, N06AA09, N06AA10, N06AA11, N06AA12, N06AA13, N06AA14, N06AA15, N06AA16, N06AA17, N06AA18, N06AA19, N06AA21, N06AA23, N06AB02, N06AB03, N06AB04, N06AB05, N06AB06, N06AB07, N06AB08, N06AB09, N06AB10, N06AF01, N06AF02, N06AF03, N06AF04, N06AF05, N06AF06, N06AG02, N06AG03, N06AX01, N06AX02, N06AX03, N06AX04, N06AX05, N06AX06, N06AX07, N06AX08, N06AX09, N06AX10, N06AX11, N06AX12, N06AX13, N06AX14, N06AX15, N06AX16, N06AX17, N06AX18, N06AX19, N06AX21, N06AX22, N06AX23, N06AX24, N06AX25, N06AX26, N06AX27, N06BA01, N06BA02, N06BA03, N06BA04, N06BA05, N06BA06, N06BA07, N06BA08, N06BA09, N06BA10, N06BA11, N06BA12, N06BA14, N06BC01, N06BC02, N06BX01, N06BX02, N06BX03, N06BX04, N06BX05, N06BX06, N06BX07, N06BX08, N06BX09, N06BX10, N06BX11, N06BX12, N06BX13, N06BX14, N06BX15, N06BX16, N06BX17, N06BX18, N06BX21, N06CA01, N06CA02, N06DA01, N06DA02, N06DA03, N06DA04, N06DA52, N06DX01, N06DX02 |
| **Rhinological and throat antiseptics (MG25)** | R01AA02, R01AA03, R01AA04, R01AA05, R01AA06, R01AA07, R01AA08, R01AA09, R01AA10, R01AA11, R01AA12, R01AA13, R01AA14, R01AB01, R01AB02, R01AB03, R01AB05, R01AB06, R01AB07, R01AB08, R01AC01, R01AC02, R01AC03, R01AC04, R01AC05, R01AC06, R01AC07, R01AC08, R01AC51, R01AD01, R01AD02, R01AD03, R01AD04, R01AD05, R01AD06, R01AD07, R01AD08, R01AD09, R01AD11, R01AD12, R01AD13, R01AD52, R01AD53, R01AD57, R01AD58, R01AD60, R01AX01, R01AX02, R01AX03, R01AX05, R01AX06, R01AX07, R01AX08, R01AX09, R01AX10, R01AX30, R01BA01, R01BA02, R01BA03, R01BA51, R01BA52, R01BA53, R02AA01, R02AA02, R02AA03, R02AA05, R02AA06, R02AA09, R02AA10, R02AA11, R02AA12, R02AA13, R02AA14, R02AA15, R02AA16, R02AA17, R02AA18, R02AA19, R02AA20, R02AB01, R02AB02, R02AB03, R02AB04, R02AB30, R02AD01, R02AD02, R02AD03, R02AD04, R02AX01, R02AX03 |
| **Inhaled anti-obstructive drugs (MG26)** | R03AA01, R03AB02, R03AB03, R03AC02, R03AC03, R03AC04, R03AC05, R03AC06, R03AC07, R03AC08, R03AC09, R03AC10, R03AC11, R03AC12, R03AC13, R03AC14, |

| | |
|---|---|
| | R03AC15, R03AC16, R03AC17, R03AC18, R03AC19, R03AK01, R03AK02, R03AK03, R03AK04, R03AK05, R03AK06, R03AK07, R03AK08, R03AK09, R03AK10, R03AK11, R03AK13, R03AK14, R03AL01, R03AL02, R03AL03, R03AL04, R03AL05, R03AL06, R03AL07, R03AL08, R03AL09, R03AL10, R03AL11, R03AL12, R03BB01, R03BB02, R03BB03, R03BB04, R03BB05, R03BB06, R03BB07, R03BB08, R03BC01, R03BC03, R03BX01, R03CA02, R03CB01, R03CB02, R03CB03, R03CB51, R03CB53, R03CC02, R03CC03, R03CC04, R03CC05, R03CC06, R03CC07, R03CC08, R03CC09, R03CC10, R03CC11, R03CC12, R03CC13, R03CC14, R03CC53, R03CC63, R03DA01, R03DA02, R03DA03, R03DA04, R03DA05, R03DA06, R03DA07, R03DA08, R03DA09, R03DA10, R03DA11, R03DA12, R03DA20, R03DA51, R03DA54, R03DA55, R03DA57, R03DA74, R03DB01, R03DB02, R03DB03, R03DB04, R03DB05, R03DB06 |
| **Inhaled steroids (MG27)** | R03BA01, R03BA02, R03BA03, R03BA04, R03BA05, R03BA06, R03BA07, R03BA08, R03BA09 |
| **Other COPD drugs (MG28)** | R03DC01, R03DC02, R03DC03, R03DC04, R03DX01, R03DX02, R03DX03, R03DX05, R03DX06, R03DX07, R03DX08, R03DX09, R03DX10 |
| **Cold and cough preparations (MG29)** | R05CA01, R05CA02, R05CA03, R05CA04, R05CA05, R05CA06, R05CA07, R05CA08, R05CA09, R05CA10, R05CA11, R05CA12, R05CA13, R05CB01, R05CB02, R05CB03, R05CB04, R05CB05, R05CB06, R05CB07, R05CB08, R05CB09, R05CB10, R05CB11, R05CB12, R05CB13, R05CB14, R05CB15, R05CB16, R05DA01, R05DA03, R05DA04, R05DA05, R05DA06, R05DA07, R05DA08, R05DA09, R05DA10, R05DA11, R05DA12, R05DA20, R05DB01, R05DB02, R05DB03, R05DB04, R05DB05, R05DB07, R05DB09, R05DB10, R05DB11, R05DB12, R05DB13, R05DB14, R05DB15, R05DB16, R05DB17, R05DB18, R05DB19, R05DB20, R05DB21, R05DB22, R05DB23, R05DB24, R05DB25, R05DB26, R05DB27, R05DB28, R05FA01, R05FA02, R05FB01, R05FB02 |
| **Systemic antihistamines (MG30)** | R06AA01, R06AA02, R06AA04, R06AA06, R06AA07, R06AA08, R06AA09, R06AA52, R06AA54, R06AA56, R06AA57, R06AA59, R06AB01, R06AB02, R06AB03, R06AB04, R06AB05, R06AB06, R06AB07, R06AB51, R06AB52, R06AB54, R06AB56, R06AC01, R06AC02, R06AC03, R06AC04, R06AC05, R06AC06, R06AC52, R06AC53, R06AD01, R06AD02, R06AD03, R06AD04, R06AD05, R06AD06, R06AD07, R06AD08, R06AD09, R06AD52, R06AD55, R06AE01, R06AE03, R06AE04, R06AE05, R06AE06, R06AE07, R06AE09, R06AE51, R06AE53, R06AE55, R06AX01, R06AX02, R06AX03, R06AX04, R06AX05, R06AX07, R06AX08, R06AX09, R06AX11, R06AX12, R06AX13, R06AX15, R06AX16, R06AX17, R06AX18, R06AX19, R06AX21, R06AX22, R06AX23, R06AX24, R06AX25, R06AX26, R06AX27, R06AX28, R06AX29, R06AX53, R06AX58 |

### Comparison of medication groups to control population

Tables **S3 and S4** show the numbers and percentages of patients receiving at least one medication of the corresponding medication group in the year before index hospitalization, separately for COVID-hospitalized patients (S3), Influenza-hospitalized patients (S4) with their respectively age-, sex- and region- matched controls.

**Table S3: Medication Groups for COVID-19 hospitalized children as compared to controls**

| | COVID (n = 1063) | | Control (n = 10626 ) | | |
|---|---|---|---|---|---|
| **Medication Groups** | **No: n (%)** | **Yes: n(%)** | **No: n (%)** | **Yes: n (%)** | **p-value** |
| **MG 1: Anticoagulants** | 1044 (98.21) | 19 (1.79) | 10559 (99.37) | 67 (0.63) | <0.001 |
| **MG 2: Antibiotics, Antivirals, Antiprotozoals or Anthelmintics** | 832 (78.27) | 231 (21.73) | 8941 (84.14) | 1685 (15.86) | <0.001 |
| **MG 3: Insulins and other Antidiabetics** | 1060 (99.72) | 3 (0.28) | 10619 (99.93) | 7 (0.07) | 0.0554 |
| **MG 4: "heart" drugs** | 1059 (99.62) | 4 (0.38) | 10585 (99.61) | 41 (0.39) | 1 |
| **MG 5: Antihypertensives incl. Diuretics and Renin-angiotensin-aldosterone system inhibitors** | 1058 (99.53) | 5 (0.47) | 10619 (99.93) | 7 (0.07) | 0.0028 |
| **MG 6: Beta Blockers** | 1060 (99.72) | 3 (0.28) | 10619 (99.93) | 7 (0.07) | 0.0554 |
| **MG 7: Statins, Fibrates incl. Proprotein convertase subtilisin/kexin type 9 inhibitors and Inclisiran** | 1062 (99.91) | 1 (0.09) | 10626 (100) | 0 (0.0) | 0.0909 |
| **MG 8: Immunosuppressants and Immunomodulators** | 1053 (99.06) | 10 (0.94) | 10615 (99.9) | 11 (0.1) | <0.001 |
| **MG 9: Systemic Steroids** | 1041 (97.93) | 22 (2.07) | 10573 (99.5) | 53 (0.5) | <0.001 |
| **MG 10: Chemotherapy** | 1053 (99.06) | 10 (0.94) | 10622 (99.96) | 4 (0.04) | <0.001 |
| **MG 11: Iron supplements, Erythropoietic stimulating agents, Vitamin B12, folic acid** | 1050 (98.78) | 13 (1.22) | 10591 (99.67) | 35 (0.33) | <0.001 |
| **MG 12: Antacids incl. Antihistamines** | 1036 (97.46) | 27 (2.54) | 10587 (99.63) | 39 (0.37) | <0.001 |
| **MG 13: Vitamin D and other Vitamin supplements** | 1019 (95.86) | 44 (4.14) | 10488 (98.7) | 138 (1.3) | <0.001 |
| **MG 14: Caplacizumab** | 1063 (100) | 0 (0.0) | 10626 (100) | 0 (0.0) | 1 |
| **MG 15: Systemic Hemostatics** | 1063 (100) | 0 (0.0) | 10626 (100) | 0 (0.0) | 1 |
| **MG 16: Hereditary angioedema Therapeutics** | 1063 (100) | 0 (0.0) | 10626 (100) | 0 (0.0) | 1 |
| **MG 17: Peripheral Vasodilators** | 1063 (100) | 0 (0.0) | 10626 (100) | 0 (0.0) | 1 |
| **MG 18: Hormonal contraceptives and similar hormone preparations** | 1054 (99.15) | 9 (0.85) | 10557 (99.35) | 69 (0.65) | 0.4278 |
| **MG 19: Immunoglobulins** | 1058 (99.53) | 5 (0.47) | 10626 (100) | 0 (0.0) | <0.001 |
| **MG 20: Interferons and CSF** | 1056 (99.34) | 7 (0.66) | 10588 (99.64) | 38 (0.36) | 0.185 |
| **MG 21: NSAR and other anti-inflammatory drugs** | 992 (93.32) | 71 (6.6) | 10358 (97.48) | 268 (2.52) | <0.001 |
| **MG 22: Gout medications** | 1063 (100) | 0 (0.0) | 10624 (99.98) | 2 (0.02) | 1 |
| **MG 23: Antiepileptics** | 1037 (97.55) | 26 (2.45) | 10591 (99.67) | 35 (0.33) | <0.001 |
| **MG 24: Antipsychotics** | 1017 (95.67) | 46 (4.33) | 10505 (98.86) | 121 (1.14) | <0.001 |
| **MG 25: Rhinological and throat antiseptics** | 999 (93.98) | 64 (6.02) | 10249 (96.45) | 377 (3.55) | <0.001 |
| **MG 26: inhaled anti-obstructive drugs** | 1017 (95.67) | 46 (4.33) | 10462 (98.46) | 164 (1.54) | <0.001 |
| **MG 27: inhaled steroids** | 1030 (96.9) | 33 (3.1) | 10456 (98.4) | 170 (1.6) | 0.0012 |
| **MG 28: other COPD drugs** | 1053 (99.06) | 10 (0.94) | 10588 (99.64) | 38 (0.36) | 0.0099 |
| **MG 29: Cold and Cough preparations** | 1031 (96.99) | 32 (3.01) | 10480 (98.63) | 146 (1.37) | <0.001 |
| **MG 30: Systemic Antihistamines** | 1033 (97.18) | 30 (2.82) | 10393 (97.81) | 233 (2.19) | 0.1922 |

**Table S4: Medication Groups for Influenza-hospitalized children as compared to controls**

| | Influenza (n = 2781) | | Control (n = 27634) | | |
|---|---|---|---|---|---|
| **Medication Groups** | **No: n (%)** | **Yes: n(%)** | **No: n (%)** | **Yes: n (%)** | **p-value** |
| **MG 1: Anticoagulants** | 2765 (99.42) | 16 (0.58) | 27578 (99.8) | 56 (0.2) | <0.001 |
| **MG 2: Antibiotics, Antivirals, Antiprotozoals or Anthelmintics** | 1632 (58.68) | 1149 (41.32) | 19902 (72.02) | 7732 (27.98) | <0.001 |
| **MG 3: Insulins and other Antidiabetics** | 2772 (99.68) | 9 (0.32) | 27618 (99.94) | 16 (0.06) | <0.001 |
| **MG 4: "heart" drugs** | 2773 (99.71) | 8 (0.29) | 27584 (99.82) | 50 (0.18) | 0.2474 |
| **MG 5: Antihypertensives incl. Diuretics and Renin-angiotensin-aldosterone system inhibitors** | 2770 (99.6) | 11 (0.4) | 27626 (99.97) | 8 (0.03) | <0.001 |
| **MG 6: Beta Blockers** | 2779 (99.93) | 2 (0.07) | 27627 (99.97) | 7 (0.03) | 0.196 |
| **MG 7: Statins, Fibrates incl. Proprotein convertase subtilisin/kexin type 9 inhibitors and Inclisiran** | 2780 (99.96) | 1 (0.04) | 27630 (99.99) | 4 (0.01) | 0.3809 |
| **MG 8: Immunosuppressants and Immunomodulators** | 2769 (99.57) | 12 (0.43) | 27620 (99.95) | 14 (0.05) | <0.001 |
| **MG 9: Systemic Steroids** | 2734 (98.31) | 47 (1.69) | 27495 (99.5) | 139 (0.5) | <0.001 |
| **MG 10: Chemotherapy** | 2774 (99.75) | 7 (0.25) | 27630 (99.99) | 4 (0.01) | <0.001 |
| **MG 11: Iron supplements, Erythropoietic stimulating agents, Vitamin B12, folic acid** | 2758 (99.17) | 23 (0.83) | 27533 (99.63) | 101 (0.37) | <0.001 |
| **MG 12: Antacids incl. Antihistamines** | 2744 (98.67) | 37 (1.33) | 27563 (99.74) | 71 (0.26) | <0.001 |
| **MG 13: Vitamin D and other Vitamin supplements** | 2700 (97.09) | 81 (2.91) | 27188 (98.39) | 446 (1.61) | <0.001 |
| **MG 14: Caplacizumab** | 2781 (100) | 0 (0.0) | 27634 (100) | 0 (0.0) | 1 |
| **MG 15: Systemic Hemostatics** | 2781 (100) | 0 (0.0) | 27634 (100) | 0 (0.0) | 1 |
| **MG 16: Hereditary angioedema Therapeutics** | 2781 (100) | 0 (0.0) | 27634 (100) | 0 (0.0) | 1 |
| **MG 17: Peripheral Vasodilators** | 2780 (99.96) | 1 (0.04) | 27633 (100) | 1 (0) | 0.1745 |
| **MG 18: Hormonal contraceptives and similar hormone preparations** | 2754 (99.03) | 27 (0.97) | 27388 (99.11) | 246 (0.89) | 0.6725 |
| **MG 19: Immunoglobulins** | 2778 (99.89) | 3 (0.11) | 27634 (100) | 0 (0.0) | <0.001 |
| **MG 20: Interferons and CSF** | 2748 (98.81) | 33 (1.19) | 27388 (99.11) | 246 (0.89) | 0.1177 |
| **MG 21: NSAR and other anti-inflammatory drugs** | 2599 (93.46) | 182 (6.54) | 26452 (95.72) | 1182 (4.28) | <0.001 |
| **MG 22: Gout medications** | 2779 (99.93) | 2 (0.07) | 27633 (100) | 1 (0.0) | 0.0235 |
| **MG 23: Antiepileptics** | 2711 (97.48) | 70 (2.52) | 27583 (99.82) | 51 (0.18) | <0.001 |
| **MG 24: Antipsychotics** | 2677 (96.26) | 104 (3.74) | 27387 (99.11) | 247 (0.89) | <0.001 |
| **MG 25: Rhinological and throat antiseptics** | 2568 (92.34) | 213 (7.66) | 26269 (95.06) | 1365 (4.94) | <0.001 |
| **MG 26: inhaled anti-obstructive drugs** | 2589 (93.1) | 192 (6.9) | 26892 (97.31) | 742 (2.69) | <0.001 |
| **MG 27: inhaled steroids** | 2588 (93.06) | 193 (6.94) | 26544 (96.06) | 1090 (3.94) | <0.001 |
| **MG 28: other COPD drugs** | 2719 (97.77) | 62 (2.23) | 27195 (98.41) | 439 (1.59) | 0.0151 |
| **MG 29: Cold and Cough preparations** | 2657 (95.54) | 124 (4.46) | 26883 (97.28) | 751 (2.72) | <0.001 |
| **MG 30: Systemic Antihistamines** | 2652 (95.36) | 129 (4.64) | 26703 (96.63) | 931 (3.37) | <0.001 |

### Outcome: Time to hospital discharge

Tables **S5 and S6** show the results of the simple and multivariable Cox-Regression models to evaluate associations between medication groups and hospital discharge, separately for COVID-hospitalized children (**S5**) and Influenza-hospitalized children (**S6**). Note that all confounder with a p<0.1 in the simple models were further investigated in the multivariable model.

**Table S5: Results of simple and multivariable Cox models for hospital discharge after COVID-19 hospitalisation**

| | Simple | | Multivariable | |
|---|---|---|---|---|
| **Confounder** | **HR (CI)** | **p-value** | **HR (CI)** | **p- value** |
| **Age Group: 6-10 (ref: 0-5)** | 0.85 (0.80-0.91) | <0.001 | 0.84 (0.78-0.91) | <0.001 |
| **Age Group: 11-15 (ref: 0-5)** | 0.96 (0.89-1.04) | 0.3016 | 0.96 (0.91-1.01) | 0.0963 |
| **Age Group: 16-18 (ref: 0-5)** | 0.91 (0.76-1.09) | 0.3028 | 0.90 (0.75-1.09) | 0.2782 |
| **Sex: W (ref: M)** | 1.12 (0.99-1.26) | 0.0759 | 1.12 (0.97-1.29) | 0.1323 |
| **Number Medication groups: 1 (ref: 0)** | 1.00 (0.94-1.07) | 0.9775 | 1.01 (0.87-1.17) | 0.9035 |
| **Number Medication groups: >=2 (ref: 0)** | 0.76 (0.63-0.91) | 0.0026 | 0.95 (0.74-1.23) | 0.7160 |
| **Wave** | 1.10 (1.02-1.19) | 0.0111 | 1.12 (1.05-1.21) | 0.0011 |
| **Wave** | 1.08 (0.95-1.23) | 0.2255 | 1.11 (0.97-1.28) | 0.1230 |
| **Anticoagulants: Yes (ref: No)** | 0.67 (0.35-1.28) | 0.2257 | | |
| **Antibiotics, Antivirals, Antiprotozoals or Anthelmintics: Yes (ref: No)** | 1.18 (0.97-1.42) | 0.0910 | 1.04 (0.84-1.30) | 0.6975 |
| **Insulins and other Antidiabetics: Yes (ref: No)** | 0.41 (0.11-1.61) | 0.2027 | | |
| **"heart" drugs: Yes (ref: No)** | 0.37 (0.13-1.00) | 0.0511 | 0.79 (0.25-2.46) | 0.6819 |
| **Antihypertensives incl. Diuretics and Renin-angiotensin-aldosterone system inhibitors: Yes (ref: No)** | 0.29 (0.10-0.84) | 0.0225 | 0.58 (0.18-1.91) | 0.3711 |
| **Immunosuppressants and Immunomodulators: Yes (ref: No)** | 0.86 (0.55-1.34) | 0.4973 | | |
| **Systemic Steroids: Yes (ref: No)** | 0.88 (0.69-1.12) | 0.2850 | | |
| **Chemotherapy: Yes (ref: No)** | 0.69 (0.44-1.06) | 0.0880 | 0.85 (0.46-1.58) | 0.6044 |
| **Iron supplements, Erythropoietic stimulating agents, Vitamin B12, folic acid: Yes (ref: No)** | 0.40 (0.23-0.69) | <0.001 | 0.43 (0.25-0.73) | 0.0019 |
| **Antacids incl. Antihistamines: Yes (ref: No)** | 0.61 (0.46-0.80) | <0.001 | 0.89 (0.70-1.13) | 0.3273 |
| **Vitamin D and other Vitamin supplements: Yes (ref: No)** | 0.67 (0.59-0.75) | <0.001 | 0.82 (0.74-0.92) | <0.001 |
| **Hormonal contraceptives and similar hormone preparations: Yes (ref: No)** | 1.07 (0.65-1.77) | 0.7796 | | |
| **Interferons and CSF: Yes (ref: No)** | 0.62 (0.45-0.85) | 0.0030 | 0.69 (0.56-0.85) | <0.001 |
| **NSAR and other anti-inflammatory drugs: Yes (ref: No)** | 1.00 (0.74-1.35) | 0.9924 | | |
| **Antiepileptics: Yes (ref: No)** | 0.47 (0.37-0.60) | <0.001 | 0.58 (0.41-0.80) | <0.001 |
| **Antipsychotics: Yes (ref: No)** | 0.71 (0.49-1.04) | 0.0782 | 0.85 (0.60-1.21) | 0.3644 |
| **Rhinological and throat antiseptics: Yes (ref: No)** | 1.18 (1.08-1.30) | <0.001 | 1.15 (1.01-1.31) | 0.0289 |
| **inhaled anti-obstructive drugs: Yes (ref: No)** | 0.92 (0.73-1.16) | 0.4709 | | |
| **inhaled steroids: Yes (ref: No)** | 1.34 (0.99-1.80) | 0.0551 | 1.31 (1.00-1.72) | 0.0489 |
| **other COPD drugs: Yes (ref: No)** | 0.75 (0.40-1.40) | 0.3688 | | |
| **Cold and Cough preparations: Yes (ref: No)** | 1.02 (0.74-1.42) | 0.8942 | | |
| **Systemic Antihistamines: Yes (ref: No)** | 1.16 (0.82-1.65) | 0.3905 | | |

**Table S6: Results of simple and multivariable Cox models for hospital discharge after Influenza hospitalisation**

| | Simple | | Multivariable | |
|---|---|---|---|---|
| **Confounder** | **HR (CI)** | **p-value** | **HR (CI)** | **p- value** |
| **Age Group: 6-10 (ref: 0-5)** | 1.16 (1.09-1.24) | <0.001 | 1.23 (1.14-1.32) | <0.001 |
| **Age Group: 11-15 (ref: 0-5)** | 1.21 (1.01-1.46) | 0.0409 | 1.31 (1.06-1.61) | 0.0120 |
| **Age Group: 16-18 (ref: 0-5)** | 1.46 (1.22-1.75) | <0.001 | 1.54 (1.28-1.86) | <0.001 |
| **Sex: W (ref: M)** | 0.97 (0.89-1.05) | 0.4395 | 0.97 (0.89-1.06) | 0.4474 |
| **Number Medication groups: 1 (ref: 0)** | 0.89 (0.80-0.99) | 0.0272 | 0.91 (0.81-1.01) | 0.0817 |
| **Number Medication groups: >=2 (ref: 0)** | 0.63 (0.54-0.73) | <0.001 | 0.80 (0.69-0.93) | 0.0029 |
| **Wave** | 1.27 (1.05-1.54) | 0.0158 | 1.26 (1.01-1.58) | 0.0428 |
| **Wave** | 1.28 (1.11-1.48) | <0.001 | 1.29 (1.10-1.51) | 0.0016 |
| **Wave** | 1.34 (1.11-1.61) | 0.0027 | 1.32 (1.08-1.62) | 0.0069 |
| **Wave** | 1.46 (1.23-1.73) | <0.001 | 1.45 (1.21-1.74) | <0.001 |
| **Wave** | NA | NA | NA (NA-NA) | NA |
| **Wave** | 0.83 (0.57-1.21) | 0.3305 | 0.83 (0.54-1.28) | 0.4058 |
| **Anticoagulants: Yes (ref: No)** | 0.93 (0.51-1.70) | 0.8205 | | |
| **Antibiotics, Antivirals, Antiprotozoals or Anthelmintics: Yes (ref: No)** | 1.02 (0.88-1.18) | 0.8297 | | |
| **Insulins and other Antidiabetics: Yes (ref: No)** | 1.26 (0.76-2.10) | 0.3700 | | |
| **"heart" drugs: Yes (ref: No)** | 2.07 (1.06-4.04) | 0.0334 | 1.77 (0.90-3.49) | 0.1005 |
| **Antihypertensives incl. Diuretics and Renin-angiotensin-aldosterone system inhibitors: Yes (ref: No)** | 0.45 (0.21-0.94) | 0.0331 | 0.62 (0.36-1.07) | 0.0880 |
| **Immunosuppressants and Immunomodulators: Yes (ref: No)** | 0.55 (0.26-1.16) | 0.1156 | | |
| **Systemic Steroids: Yes (ref: No)** | 0.88 (0.63-1.23) | 0.4662 | | |
| **Chemotherapy: Yes (ref: No)** | 0.67 (0.60-0.75) | <0.001 | 0.61 (0.42-0.88) | 0.0075 |
| **Iron supplements, Erythropoietic stimulating agents, Vitamin B12, folic acid: Yes (ref: No)** | 0.72 (0.34-1.53) | 0.3986 | | |
| **Antacids incl. Antihistamines: Yes (ref: No)** | 0.45 (0.31-0.66) | <0.001 | 0.51 (0.38-0.68) | <0.001 |
| **Vitamin D and other Vitamin supplements: Yes (ref: No)** | 0.75 (0.67-0.84) | <0.001 | 0.76 (0.66-0.87) | <0.001 |
| **Hormonal contraceptives and similar hormone preparations: Yes (ref: No)** | 0.97 (0.80-1.18) | 0.7767 | | |
| **Interferons and CSF: Yes (ref: No)** | 1.16 (0.92-1.48) | 0.2097 | | |
| **NSAR and other anti-inflammatory drugs: Yes (ref: No)** | 0.99 (0.89-1.10) | 0.9071 | | |
| **Antiepileptics: Yes (ref: No)** | 0.51 (0.40-0.64) | <0.001 | 0.55 (0.45-0.68) | <0.001 |
| **Antipsychotics: Yes (ref: No)** | 0.73 (0.58-0.92) | 0.0076 | 0.91 (0.77-1.06) | 0.2273 |
| **Rhinological and throat antiseptics: Yes (ref: No)** | 0.89 (0.81-0.97) | 0.0084 | 0.91 (0.79-1.04) | 0.1703 |
| **inhaled anti-obstructive drugs: Yes (ref: No)** | 0.84 (0.71-1.00) | 0.0476 | 0.89 (0.77-1.02) | 0.0957 |
| **inhaled steroids: Yes (ref: No)** | 0.99 (0.82-1.19) | 0.8866 | | |
| **other COPD drugs: Yes (ref: No)** | 1.12 (0.96-1.32) | 0.1559 | | |
| **Cold and Cough preparations: Yes (ref: No)** | 1.13 (0.92-1.38) | 0.2609 | | |
| **Systemic Antihistamines: Yes (ref: No)** | 1.03 (0.79-1.33) | 0.8412 | | |

**Figure S1: Kaplan-Meier curves and corresponding 95% confidence intervals for probability of hospital-discharge by age group within COVID-hospitalised (A) and Influenza-hospitalized (B) children.**

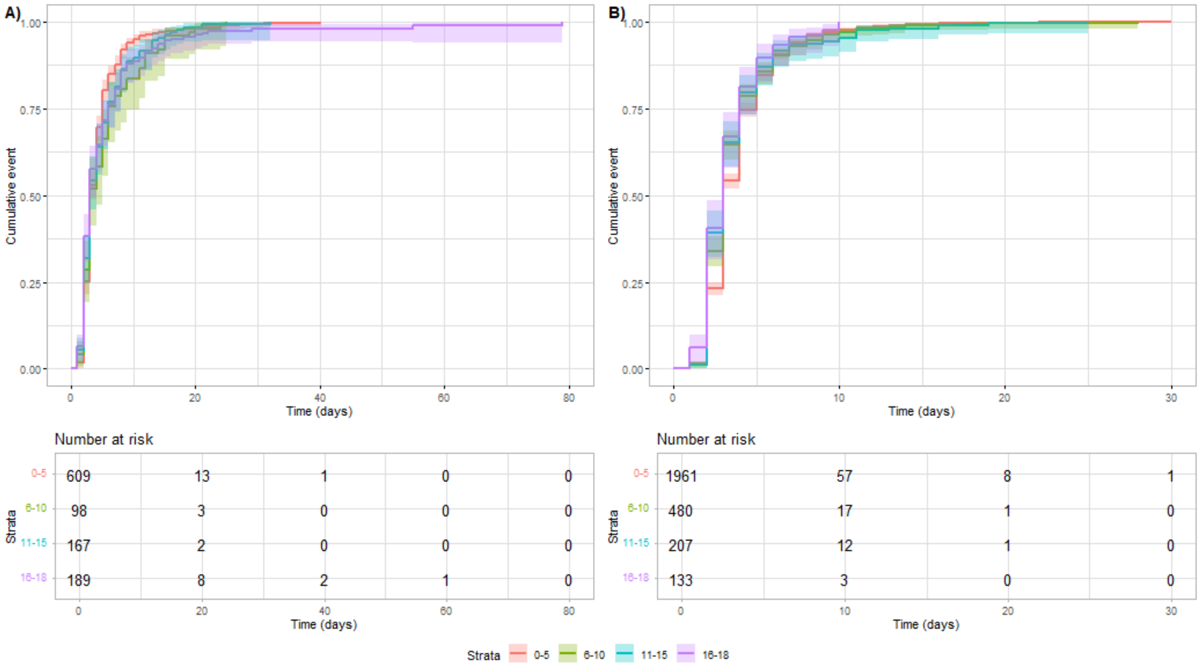


**Figure S2: Kaplan-Meier curves and corresponding 95% confidence intervals for probability of hospital-discharge for the number of medication groups within COVID- hospitalised (A) and Influenza-hospitalized (B) children.**

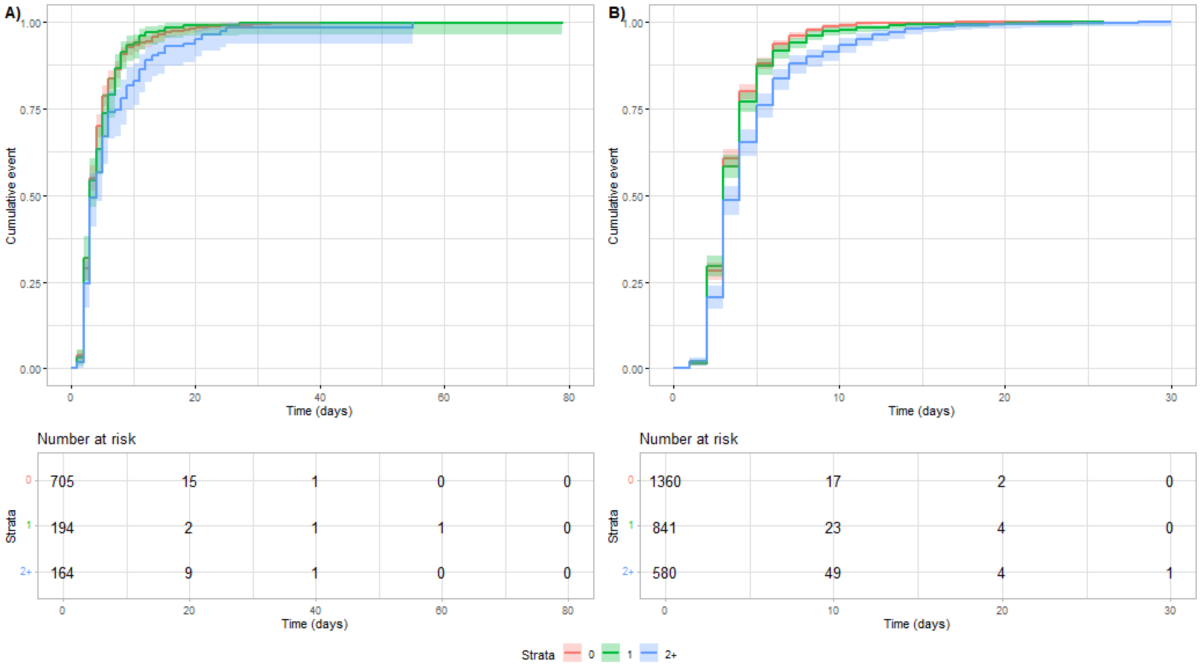

Table **S7** shows the results of the simple and multivariable Cox-Regression models to evaluate associations between propensity score matched COVID-hospitalized children and Influenza-hospitalized children. Note that all confounder with a p<0.1 in the simple models were further investigated in the multivariable model.

**Table S7: Results of the simple and multivariable Cox regression models to compare COVID-19 and Influenza patients for hospital discharge.**

| | Simple | | Multivariable | |
|---|---|---|---|---|
| | **HR (CI)** | **p-value** | **HR (CI)** | **p- value** |
| **Group: Flu (ref: Covid)** | 1.23 (0.95-1.57) | 0.111 | 1.22 (0.97-1.55) | 0.0928 |
| **Age Group: 6-10 (ref: 0-5)** | 1.05 (0.92-1.21) | 0.4523 | 1.05 (0.91-1.22) | 0.4993 |
| **Age Group: 11-15 (ref: 0-5)** | 1.09 (0.95-1.24) | 0.2256 | 1.09 (0.94-1.27) | 0.2335 |
| **Age Group: 16-18 (ref: 0-5)** | 1.22 (1.04-1.45) | 0.0172 | 1.22 (1.00-1.49) | 0.0551 |
| **Sex: W (ref: M)** | 1.00 (0.92-1.08) | 0.9293 | 1.00 (0.92-1.10) | 0.9235 |
| **Number Medication groups: 1 (ref: 0)** | 0.86 (0.76-0.97) | 0.0129 | 0.89 (0.78-1.01) | 0.0712 |
| **Number Medication groups: >=2 (ref: 0)** | 0.73 (0.60-0.90) | 0.0032 | 0.83 (0.62-1.12) | 0.2267 |
| **Anticoagulants: Yes (ref: No)** | 1.61 (0.82-3.14) | 0.1668 | | |
| **Antibiotics, Antivirals, Antiprotozoals or Anthelmintics: Yes (ref: No)** | 1.06 (0.95-1.17) | 0.3104 | | |
| **Insulins and other Antidiabetics: Yes (ref: No)** | 2.13 (0.78-5.77) | 0.1384 | | |
| **"heart" drugs: Yes (ref: No)** | 0.90 (0.55-1.46) | 0.6624 | | |
| **Antihypertensives incl. Diuretics and Renin-angiotensin-aldosterone system inhibitors: Yes (ref: No)** | 0.48 (0.28-0.84) | 0.0102 | 0.84 (0.42-1.68) | 0.6174 |
| **Immunosuppressants and Immunomodulators: Yes (ref: No)** | 0.81 (0.43-1.54) | 0.5221 | | |
| **Systemic Steroids: Yes (ref: No)** | 0.87 (0.69-1.10) | 0.255 | | |
| **Chemotherapy: Yes (ref: No)** | NA | NA | | |
| **Iron supplements, Erythropoietic stimulating agents, Vitamin B12, folic acid: Yes (ref: No)** | 1.06 (0.66-1.70) | 0.8139 | | |
| **Antacids incl. Antihistamines: Yes (ref: No)** | 0.53 (0.42-0.67) | 0 | 0.76 (0.58-1.01) | 0.0544 |
| **Vitamin D and other Vitamin supplements: Yes (ref: No)** | 0.69 (0.57-0.83) | 0.0001 | 0.70 (0.58-0.86) | 0.0005 |
| **Hormonal contraceptives and similar hormone preparations: Yes (ref: No)** | 1.14 (0.73-1.79) | 0.5604 | | |
| **Interferons and CSF: Yes (ref: No)** | 0.82 (0.53-1.25) | 0.3509 | | |
| **NSAR and other anti-inflammatory drugs: Yes (ref: No)** | 0.95 (0.71-1.28) | 0.7531 | | |
| **Antiepileptics: Yes (ref: No)** | 0.48 (0.32-0.73) | 0.0006 | 0.51 (0.27-0.97) | 0.0413 |
| **Antipsychotics: Yes (ref: No)** | 0.91 (0.71-1.18) | 0.4781 | | |
| **Rhinological and throat antiseptics: Yes (ref: No)** | 1.03 (0.97-1.09) | 0.3588 | | |
| **inhaled anti-obstructive drugs: Yes (ref: No)** | 1.00 (0.63-1.59) | 0.9995 | | |
| **inhaled steroids: Yes (ref: No)** | 1.33 (0.95-1.84) | 0.092 | 1.25 (0.92-1.70) | 0.1567 |
| **other COPD drugs: Yes (ref: No)** | 0.65 (0.48-0.89) | 0.0076 | 0.63 (0.47-0.84) | 0.0017 |
| **Cold and Cough preparations: Yes (ref: No)** | 0.90 (0.77-1.05) | 0.1699 | | |
| **Systemic Antihistamines: Yes (ref: No)** | 1.37 (1.07-1.75) | 0.014 | 1.23 (0.96-1.58) | 0.107 |

### Outcome: Readmission due to any reason after hospital discharge

Tables **S8** shows the numbers and percentages of patients with and without readmission, separately for COVID-hospitalized patients, Influenza-hospitalized patients with their respectively age-, sex- and region- matched controls.

Tables **S9** shows the numbers and percentages of patients with and without readmission, for COVID-hospitalized patients and Influenza-hospitalized patients before and after propensity score matching.

**Table S8: Readmission for COVID-19 and Influenza hospitalized children as compared to controls before propensity score matching**

| | | | COVID | | | | Influenza | | | |
|---|---|---|---|---|---|---|---|---|---|---|
| | | | COVID (all) | | Controls (all) | | Influenza (all) | | Controls (all) | |
| | | | No | Yes | No | Yes | No | Yes | No | Yes |
| **Sex** | **Age** | **Time** | **n (%)** | **n (%)** | **n (%)** | **n (%)** | **n (%)** | **n (%)** | **n (%)** | **n (%)** |
| Male | 0-5 | 30-day | 325 (96.15) | 13 (3.85) | 3322 (98.49) | 51 (1.51) | 1021 (96.14) | 41 (3.86) | 10458 (99.17) | 87 (0.83) |
| | | 180-day | 311 (92.01) | 27 (7.99) | 3225 (95.61) | 148 (4.39) | 916 (86.25) | 146 (13.75) | 10175 (96.49) | 370 (3.51) |
| | | 1-year | 298 (88.17) | 40 (11.83) | 3179 (94.25) | 194 (5.75) | 839 (79) | 223 (21) | 9894 (93.83) | 651 (6.17) |
| | | 1.5-year | 298 (88.17) | 40 (11.83) | 3168 (93.92) | 205 (6.08) | 839 (79) | 223 (21) | 9656 (91.57) | 889 (8.43) |
| | 6-10 | 30-day | 40 (95.24) | 2 (4.76) | 429 (99.77) | 1 (0.23) | 257 (95.19) | 13 (4.81) | 2690 (99.59) | 11 (0.41) |
| | | 180-day | 33 (78.57) | 9 (21.43) | 424 (98.6) | 6 (1.4) | 233 (86.3) | 37 (13.7) | 2666 (98.7) | 35 (1.3) |
| | | 1-year | 31 (73.81) | 11 (26.19) | 420 (97.67) | 10 (2.33) | 213 (78.89) | 57 (21.11) | 2632 (97.45) | 69 (2.55) |
| | | 1.5-year | 31 (73.81) | 11 (26.19) | 418 (97.21) | 12 (2.79) | 213 (78.89) | 57 (21.11) | 2591 (95.93) | 110 (4.07) |
| | 11-15 | 30-day | 75 (94.94) | 4 (5.06) | 804 (99.5) | 4 (0.5) | 112 (96.55) | 4 (3.45) | 1159 (99.91) | 1 (0.09) |
| | | 180-day | 66 (83.54) | 13 (16.46) | 796 (98.51) | 12 (1.49) | 100 (86.21) | 16 (13.79) | 1145 (98.71) | 15 (1.29) |
| | | 1-year | 66 (83.54) | 13 (16.46) | 793 (98.14) | 15 (1.86) | 91 (78.45) | 25 (21.55) | 1128 (97.24) | 32 (2.76) |
| | | 1.5-year | 66 (83.54) | 13 (16.46) | 793 (98.14) | 15 (1.86) | 91 (78.45) | 25 (21.55) | 1112 (95.86) | 48 (4.14) |
| | 16-18 | 30-day | 61 (91.04) | 6 (8.96) | 691 (99.14) | 6 (0.86) | 58 (92.06) | 5 (7.94) | 627 (99.84) | 1 (0.16) |
| | | 180-day | 58 (86.57) | 9 (13.43) | 680 (97.56) | 17 (2.44) | 51 (80.95) | 12 (19.05) | 618 (98.41) | 10 (1.59) |
| | | 1-year | 55 (82.09) | 12 (17.91) | 676 (96.99) | 21 (3.01) | 48 (76.19) | 15 (23.81) | 605 (96.34) | 23 (3.66) |
| | | 1.5-year | 55 (82.09) | 12 (17.91) | 675 (96.84) | 22 (3.16) | 48 (76.19) | 15 (23.81) | 597 (95.06) | 31 (4.94) |
| Female | 0-5 | 30-day | 256 (96.24) | 10 (3.76) | 2695 (99.26) | 20 (0.74) | 862 (96.1) | 35 (3.9) | 8826 (99.17) | 74 (0.83) |
| | | 180-day | 243 (91.35) | 23 (8.65) | 2637 (97.13) | 78 (2.87) | 797 (88.85) | 100 (11.15) | 8652 (97.21) | 248 (2.79) |
| | | 1-year | 236 (88.72) | 30 (11.28) | 2609 (96.1) | 106 (3.9) | 743 (82.83) | 154 (17.17) | 8468 (95.15) | 432 (4.85) |
| | | 1.5-year | 236 (88.72) | 30 (11.28) | 2606 (95.99) | 109 (4.01) | 742 (82.72) | 155 (17.28) | 8356 (93.89) | 544 (6.11) |
| | 6-10 | 30-day | 53 (94.64) | 3 (5.36) | 558 (99.64) | 2 (0.36) | 199 (96.6) | 7 (3.4) | 2087 (99.81) | 4 (0.19) |
| | | 180-day | 49 (87.5) | 7 (12.5) | 556 (99.29) | 4 (0.71) | 184 (89.32) | 22 (10.68) | 2067 (98.85) | 24 (1.15) |
| | | 1-year | 49 (87.5) | 7 (12.5) | 556 (99.29) | 4 (0.71) | 173 (83.98) | 33 (16.02) | 2046 (97.85) | 45 (2.15) |
| | | 1.5-year | 49 (87.5) | 7 (12.5) | 556 (99.29) | 4 (0.71) | 173 (83.98) | 33 (16.02) | 2026 (96.89) | 65 (3.11) |
| | 11-15 | 30-day | 80 (93.02) | 6 (6.98) | 854 (99.53) | 4 (0.47) | 84 (92.31) | 7 (7.69) | 907 (99.78) | 2 (0.22) |
| | | 180-day | 71 (82.56) | 15 (17.44) | 843 (98.25) | 15 (1.75) | 74 (81.32) | 17 (18.68) | 897 (98.68) | 12 (1.32) |
| | | 1-year | 69 (80.23) | 17 (19.77) | 833 (97.09) | 25 (2.91) | 63 (69.23) | 28 (30.77) | 886 (97.47) | 23 (2.53) |
| | | 1.5-year | 69 (80.23) | 17 (19.77) | 832 (96.97) | 26 (3.03) | 63 (69.23) | 28 (30.77) | 869 (95.6) | 40 (4.4) |
| | 16-18 | 30-day | 111 (93.28) | 8 (6.72) | 1179 (99.49) | 6 (0.51) | 68 (97.14) | 2 (2.86) | 697 (99.71) | 2 (0.29) |
| | | 180-day | 103 (86.55) | 16 (13.45) | 1166 (98.4) | 19 (1.6) | 60 (85.71) | 10 (14.29) | 692 (99) | 7 (1) |
| | | 1-year | 100 (84.03) | 19 (15.97) | 1157 (97.64) | 28 (2.36) | 51 (72.86) | 19 (27.14) | 681 (97.42) | 18 (2.58) |
| | | 1.5-year | 100 (84.03) | 19 (15.97) | 1153 (97.3) | 32 (2.7) | 51 (72.86) | 19 (27.14) | 671 (95.99) | 28 (4.01) |

**Table S9: Readmission for COVID-19 and Influenza hospitalized children as compared to controls after propensity score matching**

| | | | All patients | | | | Patients after PSM | | | |
|---|---|---|---|---|---|---|---|---|---|---|
| | | | COVID (all) | | Influenza (all) | | COVID (PSM) | | Influenza (PSM) | |
| | | | No | Yes | No | Yes | No | Yes | No | Yes |
| Sex | Age | Time | n (%) | n (%) | n (%) | n (%) | n (%) | n (%) | n (%) | n (%) |
| Male | 0-5 | 30-day | 325 (96.15) | 13 (3.85) | 1021 (96.14) | 41 (3.86) | 318 (96.36) | 12 (3.64) | 315 (94.88) | 17 (5.12) |
| | | 180-day | 311 (92.01) | 27 (7.99) | 916 (86.25) | 146 (13.75) | 305 (92.42) | 25 (7.58) | 291 (87.65) | 41 (12.35) |
| | | 1-year | 298 (88.17) | 40 (11.83) | 839 (79) | 223 (21) | 292 (88.48) | 38 (11.52) | 273 (82.23) | 59 (17.77) |
| | | 1.5-year | 298 (88.17) | 40 (11.83) | 839 (79) | 223 (21) | 292 (88.48) | 38 (11.52) | 273 (82.23) | 59 (17.77) |
| | 6-10 | 30-day | 40 (95.24) | 2 (4.76) | 257 (95.19) | 13 (4.81) | 36 (94.74) | 2 (5.26) | 39 (90.7) | 4 (9.3) |
| | | 180-day | 33 (78.57) | 9 (21.43) | 233 (86.3) | 37 (13.7) | 30 (78.95) | 8 (21.05) | 37 (86.05) | 6 (13.95) |
| | | 1-year | 31 (73.81) | 11 (26.19) | 213 (78.89) | 57 (21.11) | 28 (73.68) | 10 (26.32) | 33 (76.74) | 10 (23.26) |
| | | 1.5-year | 31 (73.81) | 11 (26.19) | 213 (78.89) | 57 (21.11) | 28 (73.68) | 10 (26.32) | 33 (76.74) | 10 (23.26) |
| | 11-15 | 30-day | 75 (94.94) | 4 (5.06) | 112 (96.55) | 4 (3.45) | 56 (96.55) | 2 (3.45) | 52 (98.11) | 1 (1.89) |
| | | 180-day | 66 (83.54) | 13 (16.46) | 100 (86.21) | 16 (13.79) | 52 (89.66) | 6 (10.34) | 48 (90.57) | 5 (9.43) |
| | | 1-year | 66 (83.54) | 13 (16.46) | 91 (78.45) | 25 (21.55) | 52 (89.66) | 6 (10.34) | 46 (86.79) | 7 (13.21) |
| | | 1.5-year | 66 (83.54) | 13 (16.46) | 91 (78.45) | 25 (21.55) | 52 (89.66) | 6 (10.34) | 46 (86.79) | 7 (13.21) |
| | 16-18 | 30-day | 61 (91.04) | 6 (8.96) | 58 (92.06) | 5 (7.94) | 38 (90.48) | 4 (9.52) | 34 (91.89) | 3 (8.11) |
| | | 180-day | 58 (86.57) | 9 (13.43) | 51 (80.95) | 12 (19.05) | 37 (88.1) | 5 (11.9) | 30 (81.08) | 7 (18.92) |
| | | 1-year | 55 (82.09) | 12 (17.91) | 48 (76.19) | 15 (23.81) | 35 (83.33) | 7 (16.67) | 29 (78.38) | 8 (21.62) |
| | | 1.5-year | 55 (82.09) | 12 (17.91) | 48 (76.19) | 15 (23.81) | 35 (83.33) | 7 (16.67) | 29 (78.38) | 8 (21.62) |
| Female | 0-5 | 30-day | 256 (96.24) | 10 (3.76) | 862 (96.1) | 35 (3.9) | 248 (96.5) | 9 (3.5) | 259 (97.74) | 6 (2.26) |
| | | 180-day | 243 (91.35) | 23 (8.65) | 797 (88.85) | 100 (11.15) | 236 (91.83) | 21 (8.17) | 242 (91.32) | 23 (8.68) |
| | | 1-year | 236 (88.72) | 30 (11.28) | 743 (82.83) | 154 (17.17) | 229 (89.11) | 28 (10.89) | 233 (87.92) | 32 (12.08) |
| | | 1.5-year | 236 (88.72) | 30 (11.28) | 742 (82.72) | 155 (17.28) | 229 (89.11) | 28 (10.89) | 233 (87.92) | 32 (12.08) |
| | 6-10 | 30-day | 53 (94.64) | 3 (5.36) | 199 (96.6) | 7 (3.4) | 49 (94.23) | 3 (5.77) | 54 (100) | 0 (0.0) |
| | | 180-day | 49 (87.5) | 7 (12.5) | 184 (89.32) | 22 (10.68) | 47 (90.38) | 5 (9.62) | 52 (96.3) | 2 (3.7) |
| | | 1-year | 49 (87.5) | 7 (12.5) | 173 (83.98) | 33 (16.02) | 47 (90.38) | 5 (9.62) | 49 (90.74) | 5 (9.26) |
| | | 1.5-year | 49 (87.5) | 7 (12.5) | 173 (83.98) | 33 (16.02) | 47 (90.38) | 5 (9.62) | 49 (90.74) | 5 (9.26) |
| | 11-15 | 30-day | 80 (93.02) | 6 (6.98) | 84 (92.31) | 7 (7.69) | 56 (91.8) | 5 (8.2) | 53 (94.64) | 3 (5.36) |
| | | 180-day | 71 (82.56) | 15 (17.44) | 74 (81.32) | 17 (18.68) | 49 (80.33) | 12 (19.67) | 46 (82.14) | 10 (17.86) |
| | | 1-year | 69 (80.23) | 17 (19.77) | 63 (69.23) | 28 (30.77) | 47 (77.05) | 14 (22.95) | 40 (71.43) | 16 (28.57) |
| | | 1.5-year | 69 (80.23) | 17 (19.77) | 63 (69.23) | 28 (30.77) | 47 (77.05) | 14 (22.95) | 40 (71.43) | 16 (28.57) |
| | 16-18 | 30-day | 111 (93.28) | 8 (6.72) | 68 (97.14) | 2 (2.86) | 49 (89.09) | 6 (10.91) | 51 (98.08) | 1 (1.92) |
| | | 180-day | 103 (86.55) | 16 (13.45) | 60 (85.71) | 10 (14.29) | 46 (83.64) | 9 (16.36) | 44 (84.62) | 8 (15.38) |
| | | 1-year | 100 (84.03) | 19 (15.97) | 51 (72.86) | 19 (27.14) | 44 (80) | 11 (20) | 37 (71.15) | 15 (28.85) |
| | | 1.5-year | 100 (84.03) | 19 (15.97) | 51 (72.86) | 19 (27.14) | 44 (80) | 11 (20) | 37 (71.15) | 15 (28.85) |

Tables **S10 and S11** show the results of the simple and multivariable Cox-Regression models to evaluate associations between medication groups and readmission due to any reason, separately for COVID-hospitalized children (**S12**) and Influenza-hospitalized children (**S13**). Note that all confounder with a p<0.1 in the simple models were further investigated in the multivariable model.

**Table S10: Results of simple and multivarialbe Cox models for readmission after COVID-19 hospitalisation**

| | Simple | | Multivariable | |
|---|---|---|---|---|
| **Confounder** | **HR (CI)** | **p-value** | **HR (CI)** | **p-value** |
| **Age Group: 6-10 (ref: 0-5)** | 1.13 (0.61-2.07) | 0.7057 | 1.02 (0.61-1.71) | 0.9420 |
| **Age Group: 11-15 (ref: 0-5)** | 1.11 (0.70-1.74) | 0.6630 | 1.16 (0.76-1.77) | 0.4999 |
| **Age Group: 16-18 (ref: 0-5)** | 0.94 (0.61-1.45) | 0.7817 | 1.05 (0.67-1.63) | 0.8466 |
| **Sex: W (ref: M)** | 0.94 (0.67-1.30) | 0.6920 | 0.94 (0.68-1.30) | 0.6980 |
| **Number Medication groups: 1 (ref: 0)** | 1.27 (1.01-1.60) | 0.0435 | 1.16 (0.89-1.50) | 0.2703 |
| **Number Medication groups: ≥ 2 (ref: 0)** | 2.93 (2.04-4.21) | <0.001 | 1.96 (1.39-2.78) | <0.001 |
| **Wave** | 1.02 (0.75-1.40) | 0.8828 | 1.04 (0.74-1.44) | 0.8353 |
| **Wave** | 0.44 (0.26-0.74) | 0.0023 | 0.43 (0.24-0.77) | 0.0043 |
| **Length of hospital stay** | 1.03 (1.02-1.04) | <0.001 | 1.03 (1.02-1.03) | <0.001 |
| **Anticoagulants: Yes (ref: No)** | 0.64 (0.29-1.40) | 0.2609 | | |
| **Antibiotics, Antivirals, Antiprotozoals or Anthelmintics: Yes (ref: No)** | 1.28 (0.78-2.11) | 0.3342 | | |
| **Insulins and other Antidiabetics: Yes (ref: No)** | 1.44 (0.10-21.09) | 0.7922 | | |
| **"heart" drugs: Yes (ref: No)** | NA (NA-NA) | NA | | |
| **Antihypertensives incl. Diuretics and Renin-angiotensin-aldosterone system inhibitors: Yes (ref: No)** | NA (NA-NA) | NA | | |
| **Immunosuppressants and Immunomodulators: Yes (ref: No)** | 0.38 (0.06-2.32) | 0.2916 | | |
| **Systemic Steroids: Yes (ref: No)** | 1.57 (1.04-2.38) | 0.0308 | 1.42 (0.84-2.41) | 0.1936 |
| **Chemotherapy: Yes (ref: No)** | 2.45 (1.03-5.82) | 0.0423 | 2.56 (0.95-6.95) | 0.0642 |
| **Iron supplements, Erythropoietic stimulating agents, Vitamin B12, folic acid: Yes (ref: No)** | 1.50 (0.55-4.07) | 0.4249 | | |
| **Antacids incl. Antihistamines: Yes (ref: No)** | 2.01 (0.86-4.68) | 0.1070 | | |
| **Vitamin D and other Vitamin supplements: Yes (ref: No)** | 1.01 (0.64-1.60) | 0.9536 | | |
| **Hormonal contraceptives and similar hormone preparations: Yes (ref: No)** | 2.31 (1.35-3.94) | 0.0021 | 2.53 (1.48-4.33) | <0.001 |
| **Interferons and CSF: Yes (ref: No)** | 2.82 (1.41-5.63) | 0.0034 | 1.74 (0.51-5.94) | 0.3761 |
| **NSAR and other anti-inflammatory drugs: Yes (ref: No)** | 0.79 (0.56-1.11) | 0.1671 | | |
| **Antiepileptics: Yes (ref: No)** | 2.81 (1.52-5.20) | <0.001 | 2.24 (0.88-5.68) | 0.0900 |
| **Antipsychotics: Yes (ref: No)** | 1.82 (0.97-3.41) | 0.0642 | 1.47 (0.45-4.78) | 0.5223 |
| **Rhinological and throat antiseptics: Yes (ref: No)** | 0.93 (0.48-1.79) | 0.8206 | | |
| **inhaled anti-obstructive drugs: Yes (ref: No)** | 0.98 (0.54-1.77) | 0.9360 | | |
| **inhaled steroids: Yes (ref: No)** | 0.63 (0.35-1.12) | 0.1163 | | |
| **other COPD drugs: Yes (ref: No)** | 2.25 (1.17-4.35) | 0.0157 | 2.70 (1.46-5.01) | 0.0016 |
| **Cold and Cough preparations: Yes (ref: No)** | 0.74 (0.34-1.61) | 0.4428 | | |
| **Systemic Antihistamines: Yes (ref: No)** | 0.60 (0.27-1.33) | 0.2056 | | |

**Table S11: Results of simple and multivariable Cox models for readmission after Influenza hospitalisation**

| | Simple | | Multivariable | |
|---|---|---|---|---|
| **Confounder** | **HR (CI)** | **p-value** | **HR (CI)** | **p-value** |
| **Age Group: 6-10 (ref: 0-5)** | 1.01 (0.80-1.27) | 0.9549 | 0.93 (0.75-1.15) | 0.4869 |
| **Age Group: 11-15 (ref: 0-5)** | 1.40 (1.10-1.79) | 0.0071 | 1.18 (0.86-1.62) | 0.3052 |
| **Age Group: 16-18 (ref: 0-5)** | 1.31 (1.10-1.57) | 0.0026 | 1.23 (1.02-1.47) | 0.0271 |
| **Sex: W (ref: M)** | 0.87 (0.73-1.03) | 0.1155 | 0.88 (0.74-1.04) | 0.1197 |
| **Number Medication groups: 1 (ref: 0)** | 1.37 (1.09-1.72) | 0.0066 | 1.33 (1.06-1.67) | 0.0132 |
| **Number Medication groups: ≥ 2 (ref: 0)** | 2.15 (1.89-2.45) | <0.001 | 1.57 (1.23-2.02) | <0.001 |
| **Wave** | 0.99 (0.79-1.24) | 0.9316 | 0.98 (0.77-1.25) | 0.8931 |
| **Wave** | 0.65 (0.50-0.86) | 0.0023 | 0.65 (0.50-0.85) | 0.0016 |
| **Length of hospital stay** | 1.11 (1.09-1.13) | <0.001 | 1.10 (1.08-1.12) | <0.001 |
| **Anticoagulants: Yes (ref: No)** | 2.92 (1.40-6.07) | 0.0041 | 2.75 (1.04-7.27) | 0.0412 |
| **Antibiotics, Antivirals, Antiprotozoals or Anthelmintics: Yes (ref: No)** | 0.96 (0.71-1.29) | 0.7663 | | |
| **Insulins and other Antidiabetics: Yes (ref: No)** | 0.72 (0.22-2.35) | 0.5884 | | |
| **"heart" drugs: Yes (ref: No)** | 0.79 (0.25-2.51) | 0.6877 | | |
| **Antihypertensives incl. Diuretics and Renin-angiotensin-aldosterone system inhibitors: Yes (ref: No)** | 0.71 (0.33-1.55) | 0.3927 | | |
| **Immunosuppressants and Immunomodulators: Yes (ref: No)** | 1.02 (0.32-3.31) | 0.9681 | | |
| **Systemic Steroids: Yes (ref: No)** | 1.17 (0.86-1.60) | 0.3085 | | |
| **Chemotherapy: Yes (ref: No)** | 6.97 (4.72-10.31) | <0.001 | 6.33 (4.39-9.13) | <0.001 |
| **Iron supplements, Erythropoietic stimulating agents, Vitamin B12, folic acid: Yes (ref: No)** | 0.70 (0.35-1.37) | 0.2942 | | |
| **Antacids incl. Antihistamines: Yes (ref: No)** | 1.72 (1.09-2.71) | 0.0191 | 1.77 (1.28-2.44) | <0.001 |
| **Vitamin D and other Vitamin supplements: Yes (ref: No)** | 1.06 (0.74-1.53) | 0.7481 | | |
| **Hormonal contraceptives and similar hormone preparations: Yes (ref: No)** | 1.22 (0.51-2.95) | 0.6524 | | |
| **Interferons and CSF: Yes (ref: No)** | 1.31 (0.76-2.27) | 0.3311 | | |
| **NSAR and other anti-inflammatory drugs: Yes (ref: No)** | 0.96 (0.81-1.14) | 0.6398 | | |
| **Antiepileptics: Yes (ref: No)** | 2.64 (2.06-3.39) | <0.001 | 2.09 (1.73-2.53) | <0.001 |
| **Antipsychotics: Yes (ref: No)** | 1.91 (1.74-2.09) | <0.001 | 1.44 (1.29-1.61) | <0.001 |
| **Rhinological and throat antiseptics: Yes (ref: No)** | 1.06 (0.85-1.32) | 0.6161 | | |
| **inhaled anti-obstructive drugs: Yes (ref: No)** | 1.40 (1.20-1.63) | <0.001 | 1.51 (1.20-1.90) | <0.001 |
| **inhaled steroids: Yes (ref: No)** | 0.87 (0.64-1.19) | 0.3860 | | |
| **other COPD drugs: Yes (ref: No)** | 0.68 (0.36-1.30) | 0.2481 | | |
| **Cold and Cough preparations: Yes (ref: No)** | 0.80 (0.58-1.11) | 0.1791 | | |
| **Systemic Antihistamines: Yes (ref: No)** | 0.72 (0.51-1.01) | 0.0556 | 0.77 (0.51-1.16) | 0.2037 |

Table **S12** shows the results of the simple and multivariable Cox-Regression models to evaluate associations between propensity score matched COVID-hospitalized children and Influenza-hospitalized children. Note that all confounder with a p<0.1 in the simple models were further investigated in the multivariable model.

**Table S12: Results of the simple and multivariable Cox regression models to compare re-admission due to any reason after between groups.**

| | Simple | | Multivariable | |
|---|---|---|---|---|
| **Confounder** | **HR (CI)** | **p- value** | **HR (CI)** | **p- value** |
| **Group: Flu (ref: Covid)** | 1.24 (1.04-1.47) | 0.0156 | 1.23 (1.03-1.47) | 0.0214 |
| **Age Group: 6-10 (ref: 0-5)** | 0.99 (0.67-1.48) | 0.9695 | 0.97 (0.65-1.46) | 0.892 |
| **Age Group: 11-15 (ref: 0-5)** | 1.35 (0.88-2.07) | 0.1631 | 1.34 (0.87-2.09) | 0.1861 |
| **Age Group: 16-18 (ref: 0-5)** | 1.30 (0.97-1.75) | 0.0787 | 1.26 (0.91-1.77) | 0.1678 |
| **Sex: W (ref: M)** | 0.88 (0.64-1.23) | 0.4656 | 0.88 (0.64-1.21) | 0.4319 |
| **Number Medication groups: 1 (ref: 0)** | 1.53 (1.14-2.04) | 0.0042 | 1.48 (1.09-2.01) | 0.0131 |
| **Number Medication groups: >=2 (ref: 0)** | 2.51 (1.89-3.33) | 0 | 2.06 (1.55-2.73) | 0 |
| **Length of hospital stay (days)** | 1.03 (1.01-1.06) | 0.0078 | 1.03 (1.01-1.06) | 0.0106 |
| **Anticoagulants: Yes (ref: No)** | 1.76 (0.90-3.42) | 0.0986 | 1.03 (0.38-2.77) | 0.9596 |
| **Antibiotics, Antivirals, Antiprotozoals or Anthelmintics: Yes (ref: No)** | 0.87 (0.63-1.21) | 0.421 | | |
| **Insulins and other Antidiabetics: Yes (ref: No)** | 0.86 (0.08-8.95) | 0.9026 | | |
| **"heart" drugs: Yes (ref: No)** | NA | NA | NA | NA |
| **Antihypertensives incl. Diuretics and Renin-angiotensin-aldosterone system inhibitors: Yes (ref: No)** | 0.85 (0.38-1.89) | 0.6938 | | |
| **Immunosuppressants and Immunomodulators: Yes (ref: No)** | 2.16 (0.86-5.42) | 0.101 | | |
| **Systemic Steroids: Yes (ref: No)** | 1.25 (0.66-2.36) | 0.489 | | |
| **Chemotherapy: Yes (ref: No)** | NA | NA | | |
| **Iron supplements, Erythropoietic stimulating agents, Vitamin B12, folic acid: Yes (ref: No)** | 1.24 (0.42-3.73) | 0.6962 | | |
| **Antacids incl. Antihistamines: Yes (ref: No)** | 3.13 (1.25-7.83) | 0.0146 | 2.10 (0.65-6.80) | 0.2177 |
| **Vitamin D and other Vitamin supplements: Yes (ref: No)** | 1.28 (0.89-1.85) | 0.1866 | | |
| **Hormonal contraceptives and similar hormone preparations: Yes (ref: No)** | 1.36 (0.44-4.18) | 0.5942 | | |
| **Interferons and CSF: Yes (ref: No)** | 2.15 (1.20-3.87) | 0.0106 | 2.21 (1.13-4.34) | 0.0207 |
| **NSAR and other anti-inflammatory drugs: Yes (ref: No)** | 0.75 (0.47-1.22) | 0.2466 | | |
| **Antiepileptics: Yes (ref: No)** | 2.83 (1.81-4.44) | 0 | 2.13 (1.17-3.86) | 0.0128 |
| **Antipsychotics: Yes (ref: No)** | 2.02 (1.52-2.69) | 0 | 1.47 (0.79-2.73) | 0.2277 |
| **Rhinological and throat antiseptics: Yes (ref: No)** | 0.90 (0.68-1.20) | 0.4764 | | |
| **inhaled anti-obstructive drugs: Yes (ref: No)** | 1.34 (0.79-2.26) | 0.2797 | | |
| **inhaled steroids: Yes (ref: No)** | 0.80 (0.38-1.69) | 0.5547 | | |
| **other COPD drugs: Yes (ref: No)** | 0.82 (0.52-1.31) | 0.4131 | | |
| **Cold and Cough preparations: Yes (ref: No)** | 0.61 (0.32-1.17) | 0.1365 | | |
| **Systemic Antihistamines: Yes (ref: No)** | 0.90 (0.49-1.66) | 0.7438 | | |

**Figure S3: Kaplan-Meier curves and corresponding 95% confidence intervals for probability of readmission by age group within COVID-hospitalised (A) and Influenza-hospitalized (B) children.**

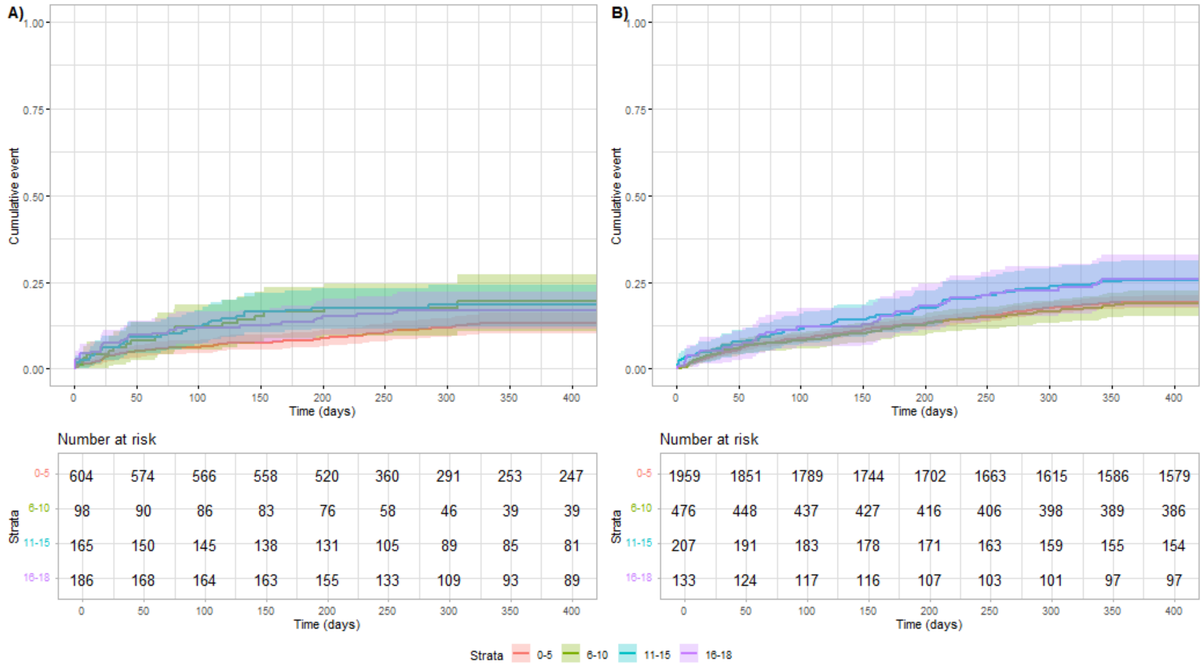


**Figure S4: Kaplan-Meier curves and corresponding 95% confidence intervals for probability of readmission for the number of medication groups within COVID- hospitalised (A) and Influenza-hospitalized (B) children.**

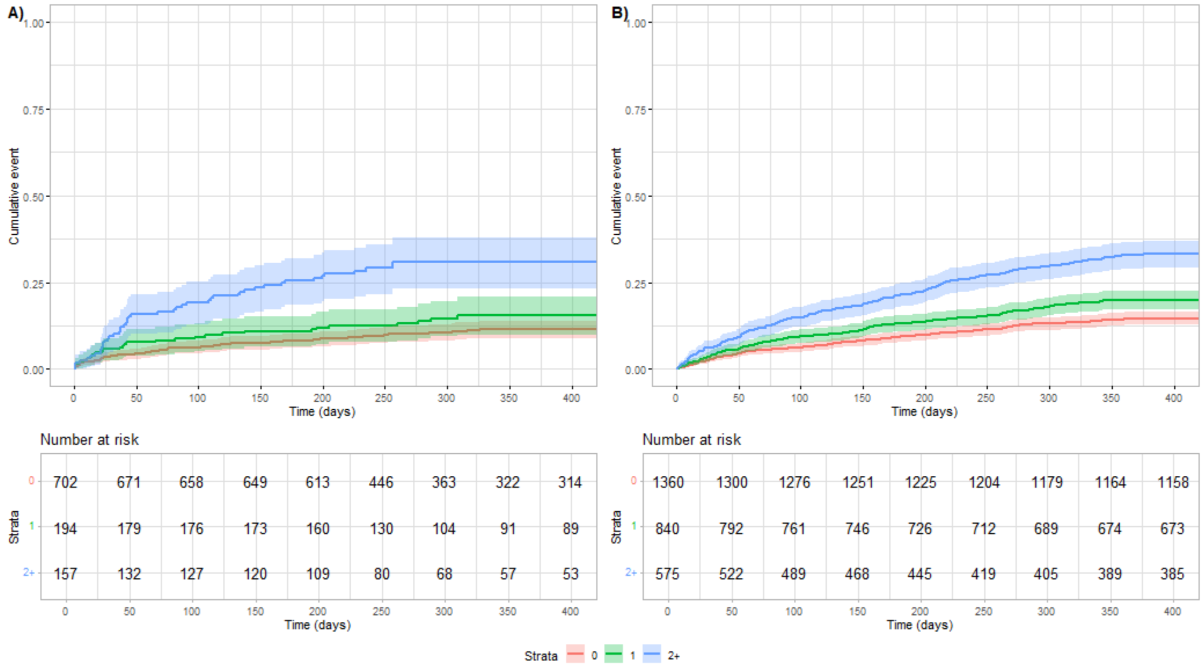

### Outcomes: short and long-term mortality

**Table S13** shows the hospital mortality and 30-day, 180-day, 1-year, and 1.5-year mortality rates for COVID- and Influenza-hospitalized patients and age-, sex- and region-matched controls separately for the four age groups and according to sex. Numbers are given for the overall patient and control cohorts.

**Table S14** shows hospital mortality and 30-day, 180-day, 1-year, and 1.5-year mortality rates for COVID- and Influenza-hospitalized patients and age-, sex- and region-matched controls separately for the four age groups and according to sex. Numbers are given for the overall patient and control cohorts, as well as for the propensity score-matched COVID and Influenza patients.

**Table S15** shows the hospital mortality and 30-day, 180-day, 1-year, and 1.5-year mortality rates for COVID-hospitalized patients and age-, sex- and region-matched controls separately for the four age groups, according to sex and the number of medication groups received (at least one medicament from the corresponding medication group at least once in the year before COVID-19 hospitalisation). Numbers are given for the overall patient and control cohorts.

**Table S16** shows the hospital mortality and 30-day, 180-day, 1-year, and 1.5-year mortality rates for Influenza-hospitalized patients and age-, sex- and region-matched controls separately for the four age groups, according to sex and the number of medication groups received (at least one medicament from the corresponding medication group at least once in the year before Influenza hospitalisation). Numbers are given for the overall patient and control cohorts.

### Table S13: Mortality for all COVID- and Influenza-patients and corresponding controls

| | | | COVID | | | | Influenza | | | |
|---|---|---|---|---|---|---|---|---|---|---|
| | | | COVID (all) | | Controls (all) | | Influenza (all) | | Controls (all) | |
| | | | Alive | Dead | Alive | Dead | Alive | Dead | Alive | Dead |
| **Sex** | **Age** | **Time** | **n (%)** | **n (%)** | **n (%)** | **n (%)** | **n (%)** | **n (%)** | **n (%)** | **n (%)** |
| **Male** | **0-5** | **Hospital** | 338 (99.71) | 1 (0.29) | 3373 (100) | 0 (0.0) | 1062 (99.91) | 1 (0.09) | 10545 (99.99) | 1 (0.01) |
| | | **30-day** | 338 (99.71) | 1 (0.29) | 3369 (99.88) | 4 (0.12) | 1062 (99.91) | 1 (0.09) | 10544 (99.98) | 2 (0.02) |
| | | **180-day** | 338 (99.71) | 1 (0.29) | 3369 (99.88) | 4 (0.12) | 1061 (99.81) | 2 (0.19) | 10542 (99.96) | 4 (0.04) |
| | | **1-year** | 172 (50.74) | 1 (0.29) | 1707 (50.61) | 4 (0.12) | 1058 (99.53) | 4 (0.38) | 10529 (99.84) | 7 (0.07) |
| | | **1.5-year** | 77 (22.71) | 1 (0.29) | 767 (22.74) | 4 (0.12) | 1058 (99.53) | 4 (0.38) | 10528 (99.83) | 8 (0.08) |
| | **6-10** | **Hospital** | 42 (100) | 0 (0.0) | 430 (100) | 0 (0.0) | 270 (100) | 0 (0.0) | 2701 (100) | 0 (0.0) |
| | | **30-day** | 42 (100) | 0 (0.0) | 430 (100) | 0 (0.0) | 270 (100) | 0 (0.0) | 2701 (100) | 0 (0.0) |
| | | **180-day** | 42 (100) | 0 (0.0) | 430 (100) | 0 (0.0) | 270 (100) | 0 (0.0) | 2701 (100) | 0 (0.0) |
| | | **1-year** | 25 (59.52) | 1 (2.38) | 270 (62.79) | 0 (0.0) | 270 (100) | 0 (0.0) | 2701 (100) | 0 (0.0) |
| | | **1.5-year** | 13 (30.95) | 1 (2.38) | 150 (34.88) | 0 (0.0) | 268 (99.26) | 2 (0.74) | 2701 (100) | 0 (0.0) |
| | **11-15** | **Hospital** | 79 (97.53) | 2 (2.47) | 808 (100) | 0 (0.0) | 116 (100) | 0 (0.0) | 1160 (100) | 0 (0.0) |
| | | **30-day** | 79 (97.53) | 2 (2.47) | 808 (100) | 0 (0.0) | 116 (100) | 0 (0.0) | 1160 (100) | 0 (0.0) |
| | | **180-day** | 79 (97.53) | 2 (2.47) | 808 (100) | 0 (0.0) | 116 (100) | 0 (0.0) | 1160 (100) | 0 (0.0) |
| | | **1-year** | 57 (70.37) | 2 (2.47) | 588 (72.77) | 0 (0.0) | 116 (100) | 0 (0.0) | 1160 (100) | 0 (0.0) |
| | | **1.5-year** | 27 (33.33) | 2 (2.47) | 270 (33.42) | 0 (0.0) | 115 (99.14) | 1 (0.86) | 1160 (100) | 0 (0.0) |
| | **16-18** | **Hospital** | 67 (95.71) | 3 (4.29) | 697 (100) | 0 (0.0) | 63 (100) | 0 (0.0) | 628 (100) | 0 (0.0) |
| | | **30-day** | 68 (97.14) | 2 (2.86) | 697 (100) | 0 (0.0) | 63 (100) | 0 (0.0) | 628 (100) | 0 (0.0) |
| | | **180-day** | 66 (94.29) | 4 (5.71) | 696 (99.86) | 1 (0.14) | 63 (100) | 0 (0.0) | 628 (100) | 0 (0.0) |
| | | **1-year** | 42 (60) | 4 (5.71) | 427 (61.26) | 1 (0.14) | 62 (98.41) | 0 (0.0) | 618 (98.41) | 0 (0.0) |
| | | **1.5-year** | 22 (31.43) | 4 (5.71) | 217 (31.13) | 1 (0.14) | 62 (98.41) | 0 (0.0) | 618 (98.41) | 0 (0.0) |
| **Female** | **0-5** | **Hospital** | 266 (98.52) | 4 (1.48) | 2715 (100) | 0 (0.0) | 897 (99.89) | 1 (0.11) | 8900 (100) | 0 (0.0) |
| | | **30-day** | 267 (98.89) | 3 (1.11) | 2714 (99.96) | 1 (0.04) | 897 (99.89) | 1 (0.11) | 8899 (99.99) | 1 (0.01) |
| | | **180-day** | 266 (98.52) | 4 (1.48) | 2712 (99.89) | 3 (0.11) | 897 (99.89) | 1 (0.11) | 8898 (99.98) | 2 (0.02) |
| | | **1-year** | 130 (48.15) | 4 (1.48) | 1331 (49.02) | 3 (0.11) | 896 (99.78) | 1 (0.11) | 8884 (99.82) | 6 (0.07) |
| | | **1.5-year** | 52 (19.26) | 4 (1.48) | 538 (19.82) | 3 (0.11) | 895 (99.67) | 2 (0.22) | 8883 (99.81) | 7 (0.08) |
| | **6-10** | **Hospital** | 56 (100) | 0 (0.0) | 560 (100) | 0 (0.0) | 206 (98.1) | 4 (1.9) | 2091 (100) | 0 (0.0) |
| | | **30-day** | 56 (100) | 0 (0.0) | 560 (100) | 0 (0.0) | 206 (98.1) | 4 (1.9) | 2091 (100) | 0 (0.0) |
| | | **180-day** | 56 (100) | 0 (0.0) | 560 (100) | 0 (0.0) | 206 (98.1) | 4 (1.9) | 2091 (100) | 0 (0.0) |
| | | **1-year** | 28 (50) | 0 (0.0) | 280 (50) | 0 (0.0) | 206 (98.1) | 4 (1.9) | 2091 (100) | 0 (0.0) |
| | | **1.5-year** | 14 (25) | 0 (0.0) | 140 (25) | 0 (0.0) | 206 (98.1) | 4 (1.9) | 2091 (100) | 0 (0.0) |
| | **11-15** | **Hospital** | 86 (100) | 0 (0.0) | 858 (100) | 0 (0.0) | 91 (100) | 0 (0.0) | 909 (100) | 0 (0.0) |
| | | **30-day** | 86 (100) | 0 (0.0) | 858 (100) | 0 (0.0) | 91 (100) | 0 (0.0) | 909 (100) | 0 (0.0) |
| | | **180-day** | 86 (100) | 0 (0.0) | 858 (100) | 0 (0.0) | 91 (100) | 0 (0.0) | 909 (100) | 0 (0.0) |
| | | **1-year** | 49 (56.98) | 0 (0.0) | 489 (56.99) | 0 (0.0) | 91 (100) | 0 (0.0) | 909 (100) | 0 (0.0) |
| | | **1.5-year** | 23 (26.74) | 0 (0.0) | 229 (26.69) | 0 (0.0) | 91 (100) | 0 (0.0) | 909 (100) | 0 (0.0) |
| | **16-18** | **Hospital** | 119 (100) | 0 (0.0) | 1185 (100) | 0 (0.0) | 70 (100) | 0 (0.0) | 699 (100) | 0 (0.0) |
| | | **30-day** | 119 (100) | 0 (0.0) | 1185 (100) | 0 (0.0) | 70 (100) | 0 (0.0) | 699 (100) | 0 (0.0) |
| | | **180-day** | 119 (100) | 0 (0.0) | 1185 (100) | 0 (0.0) | 70 (100) | 0 (0.0) | 699 (100) | 0 (0.0) |
| | | **1-year** | 75 (63.03) | 0 (0.0) | 748 (63.12) | 0 (0.0) | 69 (98.57) | 0 (0.0) | 689 (98.57) | 0 (0.0) |
| | | **1.5-year** | 42 (35.29) | 0 (0.0) | 420 (35.44) | 0 (0.0) | 69 (98.57) | 0 (0.0) | 689 (98.57) | 0 (0.0) |

**Table S14: Mortality for all COVID- and Influenza-patients before and after propensity score matching (PSM)**

| | | | All patients | | | | Patients after PSM | | | |
|---|---|---|---|---|---|---|---|---|---|---|
| | | | COVID (all) | | Influenza (all) | | COVID (PSM) | | Influenza (PSM) | |
| | | | Alive | Dead | Alive | Dead | Alive | Dead | Alive | Dead |
| Sex | Age | Time | n (%) | n (%) | n (%) | n (%) | n (%) | n (%) | n (%) | n (%) |
| Male | 0-5 | Hospital | 338 (99.71) | 1 (0.29) | 1062 (99.91) | 1 (0.09) | 330 (99.7) | 1 (0.3) | 332 (99.7) | 1 (0.3) |
| | | 30-day | 338 (99.71) | 1 (0.29) | 1062 (99.91) | 1 (0.09) | 330 (99.7) | 1 (0.3) | 332 (99.7) | 1 (0.3) |
| | | 180-day | 338 (99.71) | 1 (0.29) | 1061 (99.81) | 2 (0.19) | 330 (99.7) | 1 (0.3) | 332 (99.7) | 1 (0.3) |
| | | 1-year | 172 (50.74) | 1 (0.29) | 1058 (99.53) | 4 (0.38) | 168 (50.76) | 1 (0.3) | 331 (99.4) | 1 (0.3) |
| | | 1.5-year | 77 (22.71) | 1 (0.29) | 1058 (99.53) | 4 (0.38) | 76 (22.96) | 1 (0.3) | 331 (99.4) | 1 (0.3) |
| | 6-10 | Hospital | 42 (100) | 0 (0.0) | 270 (100) | 0 (0.0) | 38 (100) | 0 (0.0) | 43 (100) | 0 (0.0) |
| | | 30-day | 42 (100) | 0 (0.0) | 270 (100) | 0 (0.0) | 38 (100) | 0 (0.0) | 43 (100) | 0 (0.0) |
| | | 180-day | 42 (100) | 0 (0.0) | 270 (100) | 0 (0.0) | 38 (100) | 0 (0.0) | 43 (100) | 0 (0.0) |
| | | 1-year | 25 (59.52) | 1 (2.38) | 270 (100) | 0 (0.0) | 25 (65.79) | 0 (0.0) | 43 (100) | 0 (0.0) |
| | | 1.5-year | 13 (30.95) | 1 (2.38) | 268 (99.26) | 2 (0.74) | 13 (34.21) | 0 (0.0) | 43 (100) | 0 (0.0) |
| | 11-15 | Hospital | 79 (97.53) | 2 (2.47) | 116 (100) | 0 (0.0) | 58 (100) | 0 (0.0) | 53 (100) | 0 (0.0) |
| | | 30-day | 79 (97.53) | 2 (2.47) | 116 (100) | 0 (0.0) | 58 (100) | 0 (0.0) | 53 (100) | 0 (0.0) |
| | | 180-day | 79 (97.53) | 2 (2.47) | 116 (100) | 0 (0.0) | 58 (100) | 0 (0.0) | 53 (100) | 0 (0.0) |
| | | 1-year | 57 (70.37) | 2 (2.47) | 116 (100) | 0 (0.0) | 42 (72.41) | 0 (0.0) | 53 (100) | 0 (0.0) |
| | | 1.5-year | 27 (33.33) | 2 (2.47) | 115 (99.14) | 1 (0.86) | 20 (34.48) | 0 (0.0) | 53 (100) | 0 (0.0) |
| | 16-18 | Hospital | 67 (95.71) | 3 (4.29) | 63 (100) | 0 (0.0) | 42 (97.67) | 1 (2.33) | 37 (100) | 0 (0.0) |
| | | 30-day | 68 (97.14) | 2 (2.86) | 63 (100) | 0 (0.0) | 42 (97.67) | 1 (2.33) | 37 (100) | 0 (0.0) |
| | | 180-day | 66 (94.29) | 4 (5.71) | 63 (100) | 0 (0.0) | 42 (97.67) | 1 (2.33) | 37 (100) | 0 (0.0) |
| | | 1-year | 42 (60) | 4 (5.71) | 62 (98.41) | 0 (0.0) | 22 (51.16) | 1 (2.33) | 36 (97.3) | 0 (0.0) |
| | | 1.5-year | 22 (31.43) | 4 (5.71) | 62 (98.41) | 0 (0.0) | 13 (30.23) | 1 (2.33) | 36 (97.3) | 0 (0.0) |
| Female | 0-5 | Hospital | 266 (98.52) | 4 (1.48) | 897 (99.89) | 1 (0.11) | 257 (99.61) | 1 (0.39) | 265 (100) | 0 (0.0) |
| | | 30-day | 267 (98.89) | 3 (1.11) | 897 (99.89) | 1 (0.11) | 257 (99.61) | 1 (0.39) | 265 (100) | 0 (0.0) |
| | | 180-day | 266 (98.52) | 4 (1.48) | 897 (99.89) | 1 (0.11) | 257 (99.61) | 1 (0.39) | 265 (100) | 0 (0.0) |
| | | 1-year | 130 (48.15) | 4 (1.48) | 896 (99.78) | 1 (0.11) | 127 (49.22) | 1 (0.39) | 264 (99.62) | 0 (0.0) |
| | | 1.5-year | 52 (19.26) | 4 (1.48) | 895 (99.67) | 2 (0.22) | 51 (19.77) | 1 (0.39) | 264 (99.62) | 0 (0.0) |
| | 6-10 | Hospital | 56 (100) | 0 (0.0) | 206 (98.1) | 4 (1.9) | 52 (100) | 0 (0.0) | 54 (94.74) | 3 (5.26) |
| | | 30-day | 56 (100) | 0 (0.0) | 206 (98.1) | 4 (1.9) | 52 (100) | 0 (0.0) | 54 (94.74) | 3 (5.26) |
| | | 180-day | 56 (100) | 0 (0.0) | 206 (98.1) | 4 (1.9) | 52 (100) | 0 (0.0) | 54 (94.74) | 3 (5.26) |
| | | 1-year | 28 (50) | 0 (0.0) | 206 (98.1) | 4 (1.9) | 26 (50) | 0 (0.0) | 54 (94.74) | 3 (5.26) |
| | | 1.5-year | 14 (25) | 0 (0.0) | 206 (98.1) | 4 (1.9) | 13 (25) | 0 (0.0) | 54 (94.74) | 3 (5.26) |
| | 11-15 | Hospital | 86 (100) | 0 (0.0) | 91 (100) | 0 (0.0) | 61 (100) | 0 (0.0) | 56 (100) | 0 (0.0) |
| | | 30-day | 86 (100) | 0 (0.0) | 91 (100) | 0 (0.0) | 61 (100) | 0 (0.0) | 56 (100) | 0 (0.0) |
| | | 180-day | 86 (100) | 0 (0.0) | 91 (100) | 0 (0.0) | 61 (100) | 0 (0.0) | 56 (100) | 0 (0.0) |
| | | 1-year | 49 (56.98) | 0 (0.0) | 91 (100) | 0 (0.0) | 34 (55.74) | 0 (0.0) | 56 (100) | 0 (0.0) |
| | | 1.5-year | 23 (26.74) | 0 (0.0) | 91 (100) | 0 (0.0) | 17 (27.87) | 0 (0.0) | 56 (100) | 0 (0.0) |
| | 16-18 | Hospital | 119 (100) | 0 (0.0) | 70 (100) | 0 (0.0) | 55 (100) | 0 (0.0) | 52 (100) | 0 (0.0) |
| | | 30-day | 119 (100) | 0 (0.0) | 70 (100) | 0 (0.0) | 55 (100) | 0 (0.0) | 52 (100) | 0 (0.0) |
| | | 180-day | 119 (100) | 0 (0.0) | 70 (100) | 0 (0.0) | 55 (100) | 0 (0.0) | 52 (100) | 0 (0.0) |
| | | 1-year | 75 (63.03) | 0 (0.0) | 69 (98.57) | 0 (0.0) | 33 (60) | 0 (0.0) | 51 (98.08) | 0 (0.0) |
| | | 1.5-year | 42 (35.29) | 0 (0.0) | 69 (98.57) | 0 (0.0) | 17 (30.91) | 0 (0.0) | 51 (98.08) | 0 (0.0) |

**Table S15: Mortality for all COVID-patients and controls by number of medication groups received.**

| | | | COVID Patients | | | | Controls (all) | | | |
|---|---|---|---|---|---|---|---|---|---|---|
| | | | 0 | 1 | 2-5 | > 5 | 0 | 1 | 2-5 | > 5 |
| Sex | Age | | n (%) | n (%) | n (%) | n (%) | n (%) | n (%) | n (%) | n (%) |
| Male | 0-5 | Hospital | 1 (0.36) | 0 (0) | 0 (0) | 0 (0) | 0 (0) | 0 (0) | 0 (0) | 0 (0) |
| | | 30-day | 1 (0.36) | 0 (0) | 0 (0) | 0 (0) | 4 (0.14) | 0 (0) | 0 (0) | 0 (0) |
| | | 180-day | 1 (0.36) | 0 (0) | 0 (0) | 0 (0) | 4 (0.14) | 0 (0) | 0 (0) | 0 (0) |
| | | 1-year | 1 (0.36) | 0 (0) | 0 (0) | 0 (0) | 4 (0.14) | 0 (0) | 0 (0) | 0 (0) |
| | | 1.5-year | 1 (0.36) | 0 (0) | 0 (0) | 0 (0) | 4 (0.14) | 0 (0) | 0 (0) | 0 (0) |
| | 6-10 | Hospital | 0 (0) | 0 (0) | 0 (0) | 0 (0) | 0 (0) | 0 (0) | 0 (0) | 0 (0.0) |
| | | 30-day | 0 (0) | 0 (0) | 0 (0) | 0 (0) | 0 (0) | 0 (0) | 0 (0) | 0 (0.0) |
| | | 180-day | 0 (0) | 0 (0) | 0 (0) | 0 (0) | 0 (0) | 0 (0) | 0 (0) | 0 (0.0) |
| | | 1-year | 0 (0) | 0 (0) | 0 (0) | 1 (100) | 0 (0) | 0 (0) | 0 (0) | 0 (0.0) |
| | | 1.5-year | 0 (0) | 0 (0) | 0 (0) | 1 (100) | 0 (0) | 0 (0) | 0 (0) | 0 (0.0) |
| | 11-15 | Hospital | 0 (0) | 0 (0) | 1 (5) | 1 (50) | 0 (0) | 0 (0) | 0 (0) | 0 (0) |
| | | 30-day | 0 (0) | 0 (0) | 1 (5) | 1 (50) | 0 (0) | 0 (0) | 0 (0) | 0 (0) |
| | | 180-day | 0 (0) | 0 (0) | 1 (5) | 1 (50) | 0 (0) | 0 (0) | 0 (0) | 0 (0) |
| | | 1-year | 0 (0) | 0 (0) | 1 (5) | 1 (50) | 0 (0) | 0 (0) | 0 (0) | 0 (0) |
| | | 1.5-year | 0 (0) | 0 (0) | 1 (5) | 1 (50) | 0 (0) | 0 (0) | 0 (0) | 0 (0) |
| | 16-18 | Hospital | 2 (5.13) | 0 (0) | 1 (5.88) | 0 (0) | 0 (0) | 0 (0) | 0 (0) | 0 (0) |
| | | 30-day | 1 (2.56) | 0 (0) | 1 (5.88) | 0 (0) | 0 (0) | 0 (0) | 0 (0) | 0 (0) |
| | | 180-day | 2 (5.13) | 0 (0) | 1 (5.88) | 1 (100) | 0 (0) | 1 (0.81) | 0 (0) | 0 (0) |
| | | 1-year | 2 (5.13) | 0 (0) | 1 (5.88) | 1 (100) | 0 (0) | 1 (0.81) | 0 (0) | 0 (0) |
| | | 1.5-year | 2 (5.13) | 0 (0) | 1 (5.88) | 1 (100) | 0 (0) | 1 (0.81) | 0 (0) | 0 (0) |
| Female | 0-5 | Hospital | 0 (0) | 0 (0) | 2 (8) | 2 (100) | 0 (0) | 0 (0) | 0 (0) | 0 (0) |
| | | 30-day | 0 (0) | 0 (0) | 1 (4) | 2 (100) | 1 (0.04) | 0 (0) | 0 (0) | 0 (0) |
| | | 180-day | 0 (0) | 0 (0) | 2 (8) | 2 (100) | 3 (0.13) | 0 (0) | 0 (0) | 0 (0) |
| | | 1-year | 0 (0) | 0 (0) | 2 (8) | 2 (100) | 3 (0.13) | 0 (0) | 0 (0) | 0 (0) |
| | | 1.5-year | 0 (0) | 0 (0) | 2 (8) | 2 (100) | 3 (0.13) | 0 (0) | 0 (0) | 0 (0) |
| | 6-10 | Hospital | 0 (0) | 0 (0) | 0 (0) | 0 (0.0) | 0 (0) | 0 (0) | 0 (0) | 0 (0) |
| | | 30-day | 0 (0) | 0 (0) | 0 (0) | 0 (0.0) | 0 (0) | 0 (0) | 0 (0) | 0 (0) |
| | | 180-day | 0 (0) | 0 (0) | 0 (0) | 0 (0.0) | 0 (0) | 0 (0) | 0 (0) | 0 (0) |
| | | 1-year | 0 (0) | 0 (0) | 0 (0) | 0 (0.0) | 0 (0) | 0 (0) | 0 (0) | 0 (0) |
| | | 1.5-year | 0 (0) | 0 (0) | 0 (0) | 0 (0.0) | 0 (0) | 0 (0) | 0 (0) | 0 (0) |
| | 11-15 | Hospital | 0 (0) | 0 (0) | 0 (0) | 0 (0) | 0 (0) | 0 (0) | 0 (0) | 0 (0) |
| | | 30-day | 0 (0) | 0 (0) | 0 (0) | 0 (0) | 0 (0) | 0 (0) | 0 (0) | 0 (0) |
| | | 180-day | 0 (0) | 0 (0) | 0 (0) | 0 (0) | 0 (0) | 0 (0) | 0 (0) | 0 (0) |
| | | 1-year | 0 (0) | 0 (0) | 0 (0) | 0 (0) | 0 (0) | 0 (0) | 0 (0) | 0 (0) |
| | | 1.5-year | 0 (0) | 0 (0) | 0 (0) | 0 (0) | 0 (0) | 0 (0) | 0 (0) | 0 (0) |
| | 16-18 | Hospital | 0 (0) | 0 (0) | 0 (0) | 0 (0) | 0 (0) | 0 (0) | 0 (0) | 0 (0) |
| | | 30-day | 0 (0) | 0 (0) | 0 (0) | 0 (0) | 0 (0) | 0 (0) | 0 (0) | 0 (0) |
| | | 180-day | 0 (0) | 0 (0) | 0 (0) | 0 (0) | 0 (0) | 0 (0) | 0 (0) | 0 (0) |
| | | 1-year | 0 (0) | 0 (0) | 0 (0) | 0 (0) | 0 (0) | 0 (0) | 0 (0) | 0 (0) |
| | | 1.5-year | 0 (0) | 0 (0) | 0 (0) | 0 (0) | 0 (0) | 0 (0) | 0 (0) | 0 (0) |

**Table S16: Mortality for all Influenza-patients and controls by number of medication groups received.**

| | | | Influenza | | | | Control | | | |
|---|---|---|---|---|---|---|---|---|---|---|
| | | | 0 | 1 | 2-5 | > 5 | 0 | 1 | 2-5 | > 5 |
| Sex | Age | | n (%) | n (%) | n (%) | n (%) | n (%) | n (%) | n (%) | n (%) |
| Male | 0-5 | Hospital | 0 (0) | 0 (0) | 1 (0.48) | 0 (0) | 1 (0.01) | 0 (0) | 0 (0) | 0 (0) |
| | | 30-day | 0 (0) | 0 (0) | 1 (0.48) | 0 (0) | 2 (0.03) | 0 (0) | 0 (0) | 0 (0) |
| | | 180-day | 0 (0) | 1 (0.34) | 1 (0.48) | 0 (0) | 4 (0.05) | 0 (0) | 0 (0) | 0 (0) |
| | | 1-year | 1 (0.18) | 2 (0.68) | 1 (0.48) | 0 (0) | 7 (0.1) | 0 (0) | 0 (0) | 0 (0) |
| | | 1.5-year | 1 (0.18) | 2 (0.68) | 1 (0.48) | 0 (0) | 7 (0.1) | 1 (0.05) | 0 (0) | 0 (0) |
| | 6-10 | Hospital | 0 (0) | 0 (0) | 0 (0) | 0 (0) | 0 (0) | 0 (0) | 0 (0) | 0 (0) |
| | | 30-day | 0 (0) | 0 (0) | 0 (0) | 0 (0) | 0 (0) | 0 (0) | 0 (0) | 0 (0) |
| | | 180-day | 0 (0) | 0 (0) | 0 (0) | 0 (0) | 0 (0) | 0 (0) | 0 (0) | 0 (0) |
| | | 1-year | 0 (0) | 0 (0) | 0 (0) | 0 (0) | 0 (0) | 0 (0) | 0 (0) | 0 (0) |
| | | 1.5-year | 0 (0) | 0 (0) | 1 (1.82) | 1 (16.67) | 0 (0) | 0 (0) | 0 (0) | 0 (0) |
| | 11-15 | Hospital | 0 (0) | 0 (0) | 0 (0) | 0 (0.0) | 0 (0) | 0 (0) | 0 (0) | 0 (0) |
| | | 30-day | 0 (0) | 0 (0) | 0 (0) | 0 (0.0) | 0 (0) | 0 (0) | 0 (0) | 0 (0) |
| | | 180-day | 0 (0) | 0 (0) | 0 (0) | 0 (0.0) | 0 (0) | 0 (0) | 0 (0) | 0 (0) |
| | | 1-year | 0 (0) | 0 (0) | 0 (0) | 0 (0.0) | 0 (0) | 0 (0) | 0 (0) | 0 (0) |
| | | 1.5-year | 0 (0) | 0 (0) | 1 (3.03) | 0 (0.0) | 0 (0) | 0 (0) | 0 (0) | 0 (0) |
| | 16-18 | Hospital | 0 (0) | 0 (0) | 0 (0) | 0 (0.0) | 0 (0) | 0 (0) | 0 (0) | 0 (0) |
| | | 30-day | 0 (0) | 0 (0) | 0 (0) | 0 (0.0) | 0 (0) | 0 (0) | 0 (0) | 0 (0) |
| | | 180-day | 0 (0) | 0 (0) | 0 (0) | 0 (0.0) | 0 (0) | 0 (0) | 0 (0) | 0 (0) |
| | | 1-year | 0 (0) | 0 (0) | 0 (0) | 0 (0.0) | 0 (0) | 0 (0) | 0 (0) | 0 (0) |
| | | 1.5-year | 0 (0) | 0 (0) | 0 (0) | 0 (0.0) | 0 (0) | 0 (0) | 0 (0) | 0 (0) |
| Female | 0-5 | Hospital | 0 (0) | 0 (0) | 0 (0) | 1 (33.33) | 0 (0) | 0 (0) | 0 (0) | 0 (0) |
| | | 30-day | 0 (0) | 0 (0) | 0 (0) | 1 (33.33) | 1 (0.02) | 0 (0) | 0 (0) | 0 (0) |
| | | 180-day | 0 (0) | 0 (0) | 0 (0) | 1 (33.33) | 2 (0.03) | 0 (0) | 0 (0) | 0 (0) |
| | | 1-year | 0 (0) | 0 (0) | 0 (0) | 1 (33.33) | 6 (0.1) | 0 (0) | 0 (0) | 0 (0) |
| | | 1.5-year | 0 (0) | 1 (0.36) | 0 (0) | 1 (33.33) | 7 (0.11) | 0 (0) | 0 (0) | 0 (0) |
| | 6-10 | Hospital | 0 (0) | 1 (1.32) | 2 (4.55) | 1 (33.33) | 0 (0) | 0 (0) | 0 (0) | 0 (0) |
| | | 30-day | 0 (0) | 1 (1.32) | 2 (4.55) | 1 (33.33) | 0 (0) | 0 (0) | 0 (0) | 0 (0) |
| | | 180-day | 0 (0) | 1 (1.32) | 2 (4.55) | 1 (33.33) | 0 (0) | 0 (0) | 0 (0) | 0 (0) |
| | | 1-year | 0 (0) | 1 (1.32) | 2 (4.55) | 1 (33.33) | 0 (0) | 0 (0) | 0 (0) | 0 (0) |
| | | 1.5-year | 0 (0) | 1 (1.32) | 2 (4.55) | 1 (33.33) | 0 (0) | 0 (0) | 0 (0) | 0 (0) |
| | 11-15 | Hospital | 0 (0) | 0 (0) | 0 (0) | 0 (0) | 0 (0) | 0 (0) | 0 (0) | 0 (0) |
| | | 30-day | 0 (0) | 0 (0) | 0 (0) | 0 (0) | 0 (0) | 0 (0) | 0 (0) | 0 (0) |
| | | 180-day | 0 (0) | 0 (0) | 0 (0) | 0 (0) | 0 (0) | 0 (0) | 0 (0) | 0 (0) |
| | | 1-year | 0 (0) | 0 (0) | 0 (0) | 0 (0) | 0 (0) | 0 (0) | 0 (0) | 0 (0) |
| | | 1.5-year | 0 (0) | 0 (0) | 0 (0) | 0 (0) | 0 (0) | 0 (0) | 0 (0) | 0 (0) |
| | 16-18 | Hospital | 0 (0) | 0 (0) | 0 (0) | 0 (0) | 0 (0) | 0 (0) | 0 (0) | 0 (0) |
| | | 30-day | 0 (0) | 0 (0) | 0 (0) | 0 (0) | 0 (0) | 0 (0) | 0 (0) | 0 (0) |
| | | 180-day | 0 (0) | 0 (0) | 0 (0) | 0 (0) | 0 (0) | 0 (0) | 0 (0) | 0 (0) |
| | | 1-year | 0 (0) | 0 (0) | 0 (0) | 0 (0) | 0 (0) | 0 (0) | 0 (0) | 0 (0) |
| | | 1.5-year | 0 (0) | 0 (0) | 0 (0) | 0 (0) | 0 (0) | 0 (0) | 0 (0) | 0 (0) |
